# Genome-wide meta-analysis of panic attacks and disorder identifies genetic risk factors and implicates multiple body systems in panic and other psychiatric disorders

**DOI:** 10.1101/2025.06.15.25329656

**Authors:** Brittany L. Mitchell, Megan Skelton, Rujia Wang, Abigail R. ter Kuile, Alan E. Murphy, Genevieve Morneau-Vaillancourt, Danyang Li, Elham Assary, Matthew Hotopf, Jihua Hu, Anna Monistrol-Mula, Jodi T. Thomas, Ang Li, Jian Zeng, Chérie Armour, Andrew M. McIntosh, James T. R. Walters, Donald M. Lyall, Daniel J. Smith, Lifelines Cohort Study, NIHR BioResource Consortium, Nathalie Kingston, John R. Bradley, GLAD Study Authors, Gursharan Kalsi, Sang-Hyuck Lee, Yuhao (Leo) Lin, Evangelos Vassos, Saakshi Kakar, Laura Meldrum, Iona Smith, Gemma Chavarria Ventura, Catherine M. Olsen, David C. Whiteman, Sarah E. Medland, Penelope A. Lind, Enda M. Byrne, Ian B. Hickie, Naomi R. Wray, Lukas Mach, Michela Noseda, Andreas J. Forstner, Harold Snieder, Catharina A. Hartman, Nicholas G. Martin, Jürgen Deckert, Nathan Skene, Jonathan R. I. Coleman, Thalia C. Eley, Gerome Breen

**Affiliations:** QIMR Berghofer Medical Research Institute, Brisbane, Queensland, Australia; School of Biomedical Sciences, Faculty of Medicine, The University of Queensland, Brisbane, Queensland, Australia; School of Biomedical Sciences, Queensland University of Technology, Brisbane, Australia; Social, Genetic, and Developmental Psychiatry Centre, Institute of Psychiatry, Psychology and Neuroscience, King’s College London, London, UK; National Institute for Health and Care Research Maudsley Biomedical Research Centre, South London and Maudsley NHS Trust, London, UK; Department of Clinical, Educational, and Health Psychology, University College London, London, UK; UK Dementia Research Institute at Imperial College London, London, UK; Department of Brain Sciences, Imperial College London, London, UK; Research Centre of the Montreal University Institute of Mental Health, University of Montreal, Canada; Institute of Psychiatry, Psychology & Neuroscience, King’s College London, London, UK; Trauma and Mental Health Research Centre (TMHRC), School of Psychology, Queen’s University Belfast, Belfast, UK; Division of Psychiatry, Centre for Clinical Brain Sciences, University of Edinburgh, Edinburgh, UK; Centre for Neuropsychiatric Genetics and Genomics and National Centre for Mental Health, Cardiff University, Cardiff, UK; School of Health & Wellbeing, University of Glasgow, Glasgow, UK; NIHR BioResource, Cambridge Biomedical Campus, Cambridge, UK; Department of Haematology, University of Cambridge, Cambridge, Biomedical Campus, Cambridge, UK; School of Psychology, University of Queensland, Brisbane, Australia; University of Queensland Child Health Research Centre, Brisbane, Australia; Institute for Molecular Bioscience, University of Queensland, Brisbane, QLD, Australia; Department of Psychiatry, University of Oxford, Oxford, UK; Department of Bioengineering, Imperial College London, London, UK; Royal Brompton Hospital, London, UK; British Heart Foundation Centre of Research Excellence, Imperial College London, London, UK; Institute of Human Genetics, University of Bonn, School of Medicine & University Hospital Bonn, Bonn, Germany; Institute of Neuroscience and Medicine (INM-1), Research Center Jülich, Jülich, Germany; Department of Epidemiology, University of Groningen, University Medical Center Groningen, Groningen, Netherlands; Department of Psychiatry, University of Groningen, University Medical Center Groningen, Groningen, Netherlands; Institute of Clinical Epidemiology and Biometry,Medical Faculty, University of Würzburg, Würzburg, Germany; Department of Psychiatry, Psychosomatics and Psychotherapy, University Hospital, Würzburg, Germany

## Abstract

Panic attacks are sudden episodes of intense fear with physical and psychological symptoms, affecting approximately 28% of the population. Panic disorder (3-7% prevalence) involves recurrent attacks with persistent anticipatory worry, functional impairment, and avoidance. We conducted genome-wide association meta-analyses of a broad panic phenotype (panic attacks or panic disorder; 90,840 cases, 201,638 controls), panic disorder (41,721 cases, 201,638 controls), and isolated panic attacks (49,119 cases, 192,885 controls), identifying the first genome-wide significant variants for each trait (25 broad, 15 disorder, 5 attack). Gene-set analysis of single-cell RNA sequencing data for the broad phenotype implicated eye, heart, and lung neurons, pointing to sensory processing and interoceptive awareness. These peripheral neuronal associations generalised across psychiatric disorders. These findings offer novel insights into the biological mechanisms underlying panic and the overlooked role of afferent neurons across psychiatric disorders.

## Introduction

Panic attacks are sudden episodes of intense fear accompanied by physical and psychological symptoms, affecting approximately 28% of the population on a lifetime basis^1,2^. Isolated or sporadic panic attacks are infrequently studied but have reported heritability estimates >30% ^3,4^. Panic disorder has better established heritability estimates (40-50%) ^5^. Panic attacks and panic disorder, while sharing core symptomatology^6^, may differ in their genetic architecture: the acute attack is a discrete autonomic discharge event, whereas the disorder encompasses sustained anticipatory anxiety, avoidance behaviour, and functional impairment that may involve distinct neural circuit and cognitive mechanisms, and peripheral organ pathophysiology. Panic research has long implicated both local neuronal control of visceral organs and their afferent communication with the brain in panic-related pathophysiology^7–11^, and central nervous system (CNS) processes shared with other psychiatric disorders^12–14^. Clinical observations that panic attacks are accompanied by pronounced cardiorespiratory, gastrointestinal, and vestibular symptoms have led to longstanding hypotheses that afferent signalling from peripheral organs contributes to panic pathophysiology^8^, yet this has never been tested at the genetic level using tissue- and cell-type-specific approaches.

Previous genome-wide association study (GWAS) meta-analyses of panic disorder, conducted in European (effective sample size (Neff)=3,363)^15^ and Japanese populations (Neff=2,024)^16^, were underpowered to detect genome-wide significant loci. In addition, previous anxiety GWAS research has often grouped panic disorder with other anxiety disorders^17,18^. Panic disorder shows substantial phenotypic and genetic overlap with other internalising conditions, yet its episodic, paroxysmal presentation and prominent somatic symptom burden distinguish it from generalised anxiety and major depression, raising the question of whether panic-specific genetic signals exist beyond the shared internalising liability.

Despite their high prevalence and established heritability, isolated panic attacks have never been the subject of a dedicated GWAS, and it remains unknown whether their genetic architecture is distinguishable from that of the full clinical disorder. It is thus critical to examine panic attacks and disorder independently, and at scale, because their episodic nature, distinct symptomatology, and emphasis on fear may reflect distinct genetic aetiology compared to other anxiety disorders that are characterized by persistent worry. Our study addresses this gap by conducting the first dedicated GWAS meta-analysis of panic attacks to our knowledge, and the largest GWAS meta-analysis of panic disorder to date — the first to identify genome-wide significant loci — integrating data from seven international cohorts, predominantly of European ancestry. We aimed to identify novel genetic loci associated with these phenotypes with sufficient power to elucidate biological mechanisms, with particular attention to the role of peripheral organ systems.

## Results

### GWAS meta-analysis identifies novel loci for panic attacks and disorder

The cohorts in our meta-analysis utilized diverse ascertainment strategies but had similar assessments, primarily determining diagnostic status via validated self-report questionnaires based on diagnostic criteria as defined by the Diagnostic and Statistical Manual of Mental Disorders (DSM)^19,20^. Cases met lifetime diagnostic criteria for panic attacks or panic disorder, while controls were individuals without a history of panic attacks, anxiety and depressive disorders. Specific inclusion and exclusion criteria slightly varied across cohorts depending on the information available (e.g., medical records or self-report only; see Supplementary Table 1 and Supplementary Materials). Each cohort performed genotyping using microarray platforms, with the exception of the All of Us cohort which provided Whole Genome Sequence data, and followed best practice and similar pipelines for imputation (Supplementary Materials). Genome-wide association analyses were conducted using linear mixed models, adjusting for covariates such as principal components of ancestry and genotyping batch. Cohort summary statistics were then fixed-effects meta-analysed with the Forstner *et al.*^15^ summary statistics using METAL’s STDERR approach^21^, resulting in a final sample size of 90,840 panic broad cases, 41,721 panic disorder cases and 201,638 controls. A third phenotype, representing individuals who have experienced panic attacks but do not meet criteria for panic disorder, consisted of 49,119 cases and 192,885 controls. LDSC-derived metrics (lambda1000 and LDSC intercept) indicated minimal residual inflation due to confounding or cryptic relatedness in the meta-analysed summary statistics for all three phenotypes (Supplementary Table 1).

We identified 25 independent genome-wide significant loci associated with panic broad, 15 with panic disorder (Figure 1, Supplementary Figure 2, and Supplementary Table 2), and 5 with panic attacks (Supplementary Figure 1, Supplementary Figure 2, and Supplementary Table 2). Our panic broad and panic disorder GWAS had significant genetic correlation compatible with unity (r_g_=1, S.E.=0.007, p<10^-300^; Supplementary Table 3). Genetic correlations between panic attacks and both panic broad (rg=0.954, S.E.=0.007, p<10^-300^) and panic disorder (rg=0.91, S.E.=0.025, p=4.69×10^-287^) were high but significantly differed from unity (p=1.06×10⁻¹⁰ and p=2.51×10⁻⁴, respectively). There was no significant SNP effect heterogeneity across cohorts for any of our identified loci (Supplementary Table 2). Genome-wide, heterogeneity between contributing cohorts was minimal. The median I² was 0 across all three meta-analyses, with only 8.2–9.6% of variants showing I² > 50% and 5.6–6.0% reaching nominal heterogeneity (Cochran’s Q p < 0.05) — consistent with the chance expectation under the null hypothesis.

**Figure 1.**
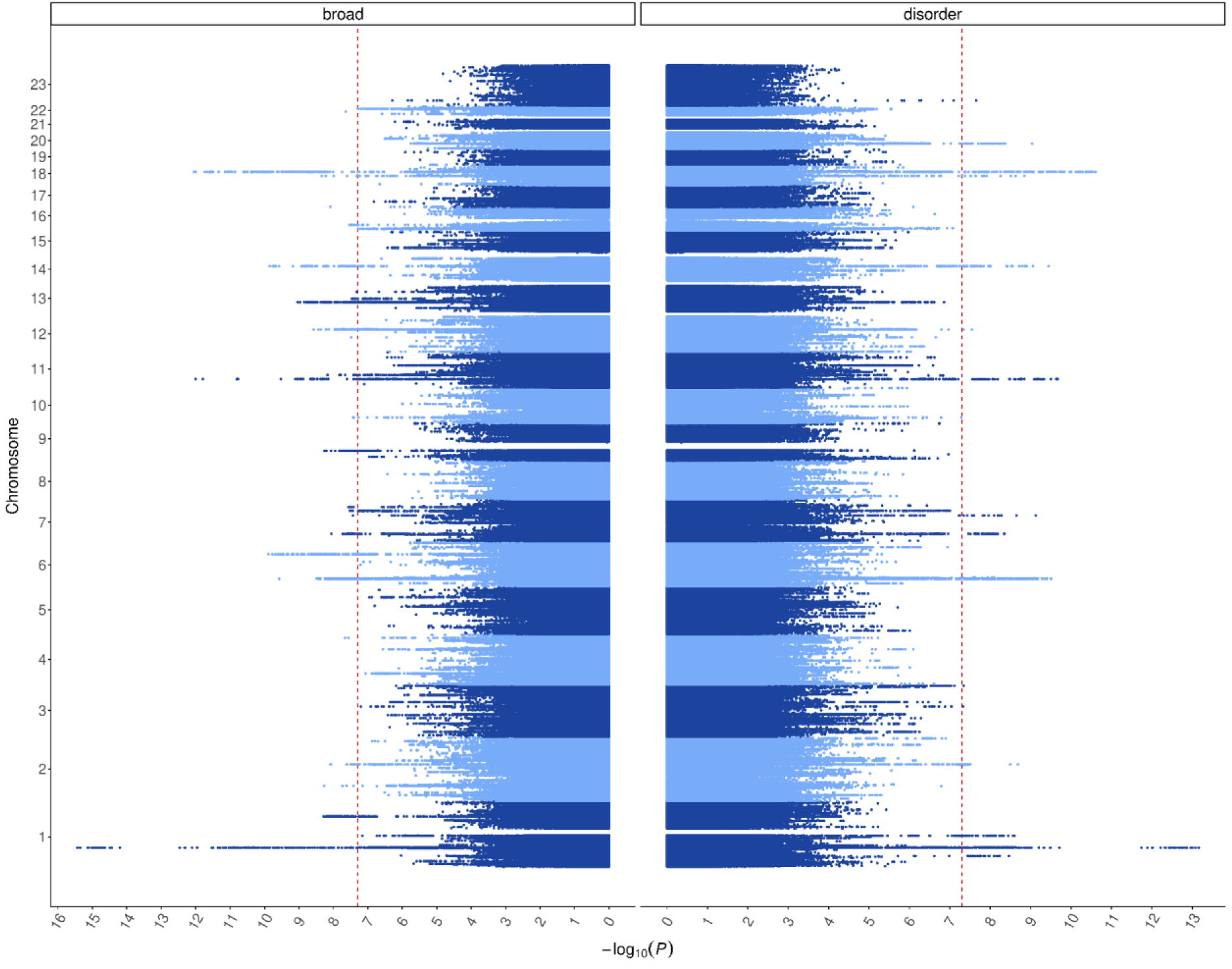
Mirror plot of the statistical significance of genome-wide variant associations for panic broad (left; case N=90,840) and panic disorder (right; case N=41,721) (with common controls without a history of panic disorder or attacks or any anxiety or depressive disorder; N=201,638). The red vertical lines indicate the genome-wide significance threshold (p < 5x10^-8^).

Summary statistics SNP-based heritability (h^2^_SNP_) estimates on the liability scale ranged from 9-12% for panic broad, 6-10% for panic disorder, and 4-7% for panic attacks across a range of probable population lifetime prevalence estimates^2^, calculated using SBayesRC^22^ (Supplementary Figure 3). Raw data h^2^_SNP_ estimates with BOLT-LMM^23^ in our largest cohort (the merged GLAD+ and UK Biobank sample) were, as expected^24^, higher; 16-27% for panic broad and 10-20% for panic disorder.

### Locus Annotation

We annotated independent significant variants using positional, chromatin-interaction, and xQTL (eQTL, sQTL, pQTL, sce-QTL) mapping using FUMA (v2.0.0; Supplementary Table 7)^25^. This is the first study to implicate these variants specifically in panic but comparisons to pre-existing GWAS^26,18,27^ revealed that 34 of the 39 variants had been associated (r^2^ > 0.1, 500kb window) with internalising phenotypes such as neuroticism, and broad definitions of anxiety^18,28^ or depression^29^. The remaining 5 novel loci included a panic disorder locus on chr 1 (lead variant rs1289881) which maps to *MAN1A2* through positional, eQTL, and sceQTL, and has been implicated in educational attainment. Another novel locus genome-wide significant for panic disorder was located on chr 6 (lead variant rs583129) in the MHC, nearest to *MYL8P* and mapping to numerous genes in this area of complex linkage disequilibrium (LD). This locus has been implicated in schizophrenia, insomnia, and arthritis. A panic broad association on chr 6 (lead variant rs11154014) mapped to *TBC1D32* and *GJA1* via position and xQTLs, and has been associated with circulatory phenotypes. A locus on chr 7 (lead variant rs10081338) was significant for panic broad and disorder, located within *HIBADH* and further maps to *TAX1BP1*, and *JAZF1*, and has evidence for associations with prostate cancer, kidney stones, and blood pressure measures. Finally, a locus on chr 22 (lead variant rs2267158) was genome-wide significant for panic broad and attacks. This locus has shown associations with measures of BMI and white matter microstructure. Only one locus was genome-wide significant across all analyses — the highest ranked variant for panic broad, rs1841499 within the *NEGR1* locus. Functionally-informed fine-mapping of loci (Supplementary Table 6) implicated possible associations of panic attacks with *PAX6* (the product of which is a master regulator of neurogenesis), with *KLC1* (product associated with neurodegeneration), and with *GAL3ST1* (product important for neuronal myelination; Supplementary Results).

For secondary analyses requiring sufficient loci (MAGMA gene-set analysis, partitioned heritability, colocalisation), we considered a minimum of 10 genome-wide significant loci as a practical threshold. The panic broad and disorder met this criterion but panic attack did not and was limited to genetic correlation analyses.

### Sensitivity analyses: effect of control screening stringency

Sensitivity analyses with controls screened only for panic (‘simple’ screening) yielded similar, though attenuated, results compared with the primary analyses that additionally excluded depression and anxiety disorders from controls (‘super’ screening; Supplementary Figures 2 and 4, Supplementary Table 4).

Super screening consistently yielded more genome-wide significant loci than simple screening despite smaller effective sample size (Neff): 25 versus 20 loci for the broad phenotype (Neff 205,747 vs 277,272), and 15 versus 11 for panic disorder (Neff 118,575 vs 140,229). This represents approximately 1.6–1.7-fold greater efficiency per unit of Neff under super screening, consistent with reduced phenotypic heterogeneity outweighing the power cost of smaller sample size (Supplementary Table 5). The panic attack phenotype was underpowered under both screening strategies (5 and 2 loci, respectively).

To assess whether loci absent in one GWAS but present in another reflected genuine phenotype specificity or insufficient power, we estimated statistical power for each locus– GWAS combination. Power was calculated from the non-centrality parameter of the discovering GWAS, scaled by the Neff ratio and adjusted for the difference in control screening stringency. Assuming r_g_ ≈ 0.7 between panic and MDD/anxiety and approximately 25% comorbid contamination in simple controls, effects are attenuated by approximately 17.5% in simple relative to super screening (Supplementary Table 5). The 1p31.1/*NEGR1* locus (rs1841499) was the only locus reaching genome-wide significance across all six GWAS. The MHC (6p22.1) and 7p15.2 loci each appeared in four of six. Several loci were specific to super screening, consistent with signals diluted by phenotypic heterogeneity in simple controls.

### Summary-data-based Mendelian Randomization

Summary-data-based Mendelian Randomization (SMR) analysis integrating eQTL, sQTL, pQTL, and mQTL data identified 10 genes showing pleiotropic associations between molecular phenotypes and panic phenotypes, consistent with a shared causal variant (Supplementary Table 8). *NEGR1* was the most consistently implicated gene, identified through pQTL analysis across panic broad and disorder, and through sQTL analysis in panic broad, suggesting that genetic effects on these panic phenotypes might be mediated through protein abundance of *NEGR1*. *SLC30A9* showed convergent evidence across different omics layers in panic broad and attacks with significant associations in both eQTL and sQTL analyses in brain tissue, indicating that genetic effects may operate through both expression and splicing of this gene. Additional identified associations are reported in Supplementary Table 8.

### Cell-type enrichment implicates central and peripheral neurons

We tested for enrichment of cell-type-specific gene expression using the broad panic GWAS, which provided the greatest statistical power of the three phenotypes. Gene expression specificity statistics^30^ were used to test for enrichment using all genes in linear regression MAGMA gene set analysis, with a second test (and effect size estimate) derived using the top 10% of genes by expression specificity, using stratified linkage disequilibrium score regression (S-LDSC)^31^ (Supplementary Figures 5-11). Cell-type specificity was derived from a human cell atlas of foetal gene expression, containing 377,456 cells from 77 distinct cell types in 15 organs^32^. Analysis was not restricted to the CNS due to prior known association of panic with the peripheral nervous system (PNS), specifically the autonomic nervous system ^33^. Where available, tissue-specific single-cell atlases were similarly tested to verify these results, including the mouse CNS^34^, human foetal and adult heart^35^, human adult lungs^36^ and human retina developmental atlases^37^. Moreover, to test if the identified cell type associations were unique to panic-related phenotypes, alternative psychiatric traits were tested, including chronic pain^38^, dimensional anxiety^39^, schizophrenia^40^, major depression^29^, and bipolar disorder^41^. Other results from MAGMA-level gene-mapping and pathway analyses can be found in Supplementary Tables 9 and 10.

The single-cell analyses for panic broad implicated a range of CNS cell types, alongside peripheral visceral neurons, at FDR < 0.05 (Supplementary Table 11). In the cerebrum, significant enrichment was observed for megakaryocytes, astrocytes, excitatory neurons, inhibitory neurons, and oligodendrocytes. In the cerebellum, granule neurons showed the strongest signal, followed by astrocytes, oligodendrocytes, unipolar brush cells, and inhibitory interneurons (Figure 2). Beyond the CNS, significant enrichment was found for visceral neurons of the heart, cardiac SATB2/LRRC7⁺ cells, visceral neurons of the lung, enteric nervous system neurons, and glia (FDR = 3.48×10⁻²) of the stomach. Multiple retinal cell types were implicated in the eye, including amacrine, PDE11A/FAM19A2⁺, horizontal, ganglion, and bipolar cells. In the heart and lung the remaining highly ranked populations — smooth muscle, lymphoid and myeloid cells — did not survive FDR correction, indicating that the significant peripheral signal was predominantly neuronal (Figure 2). Glial enrichment therefore spanned both CNS and peripheral tissues. We verified key results in independent, organ-specific single-cell transcriptomic atlases of the CNS, heart, lung and eye, although no independent data were available for retinal amacrine and ganglion cells (Figure 3, Supplementary Table 12, Supplementary Figures 5 and 6). These enrichments are consistent with longstanding hypotheses and prior studies in panic disorder^8,33,42–44^.

**Figure 2:**
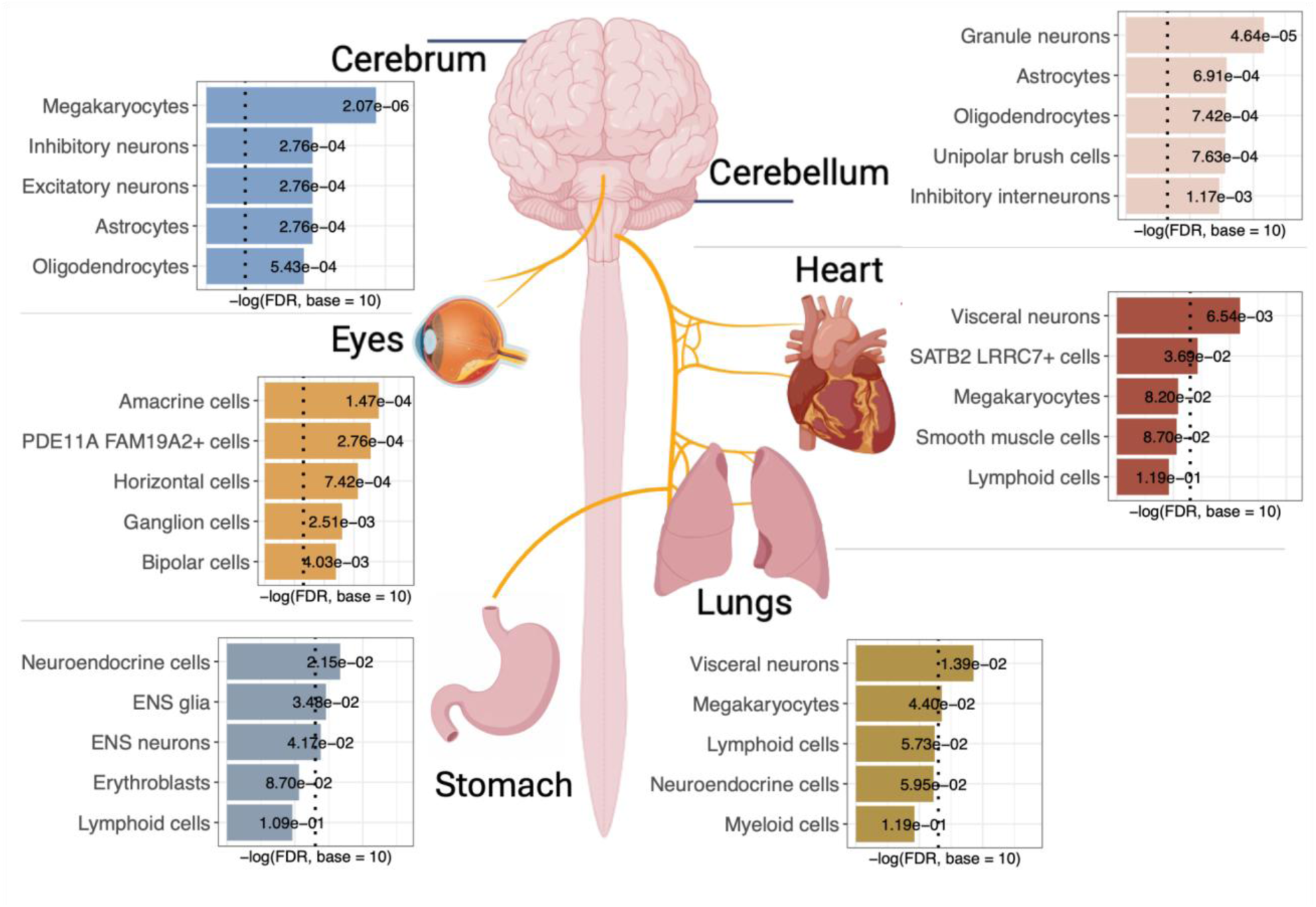
Cell type-specific central and peripheral nervous system (CNS/PNS) genetic enrichment for panic broad (panic disorder plus panic attacks) with super screened controls results, based on the whole human body, foetal developmental single-cell transcriptional dataset. The top five adjusted p-value enrichments from MAGMA gene set enrichment test (linear regression) are shown for the cerebrum, cerebellum, heart, lungs, eyes, and stomach with a red dashed vertical line indicating the false discovery rate (FDR) threshold of 0.05. The y-axis shows the -log10 FDR, where larger values indicate a greater level of significance, while the text gives the FDR values. Created with graphics from BioRender.com.

**Figure 3:**
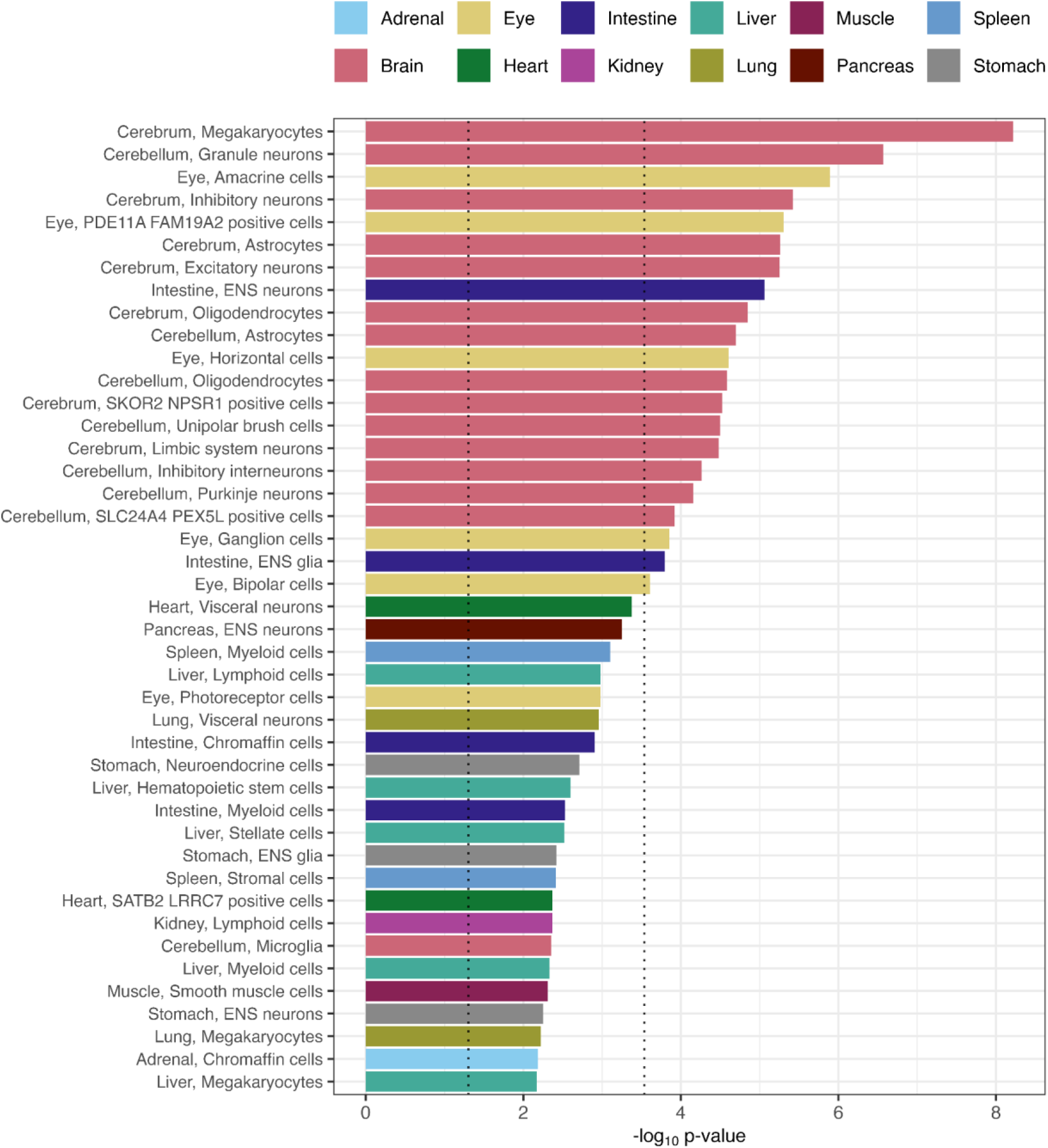
All cell type-specific, significant genetic enrichments (FDR < 0.05) for panic broad (panic disorder plus panic attacks) results based on the whole human body, foetal developmental single-cell transcriptional dataset with tissue-specific colouring. MAGMA gene set enrichment test p-value enrichments are shown for both linear regression, bars are coloured by organ of origin for the cell type. Black dashed vertical lines indicate p-value thresholds of 0.05 and Bonferroni-adjusted p-value threshold of 0.0001.

Differential enrichment analysis highlighted that while the peripheral neuronal signal was significant when compared to non-neuronal signatures (FDR=0.02), PNS neurons were not significant when compared to CNS neurons (FDR=0.95), which may indicate that the shared neuronal signal is driving enrichment, regardless of location (Table S13). Furthermore, we sought to understand whether these enrichments were unique to panic attacks or were shared with other psychiatric disorders and traits. Comparing the proportion of heritability explained using s-LDSC^45^ revealed that enrichment of these visceral and retinal neuron populations was broadly shared rather than panic-specific; comparable or stronger enrichment was observed in other disorders, most consistently bipolar disorder and schizophrenia (e.g. lung visceral neurons: bipolar disorder enrichment=2.0, FDR=7×10⁻¹¹; schizophrenia=1.8, FDR=7.7×10⁻¹⁰; panic broad=1.4, FDR=1.8×10⁻³) (Figure 4, Supplementary Table 14).

**Figure 4.**
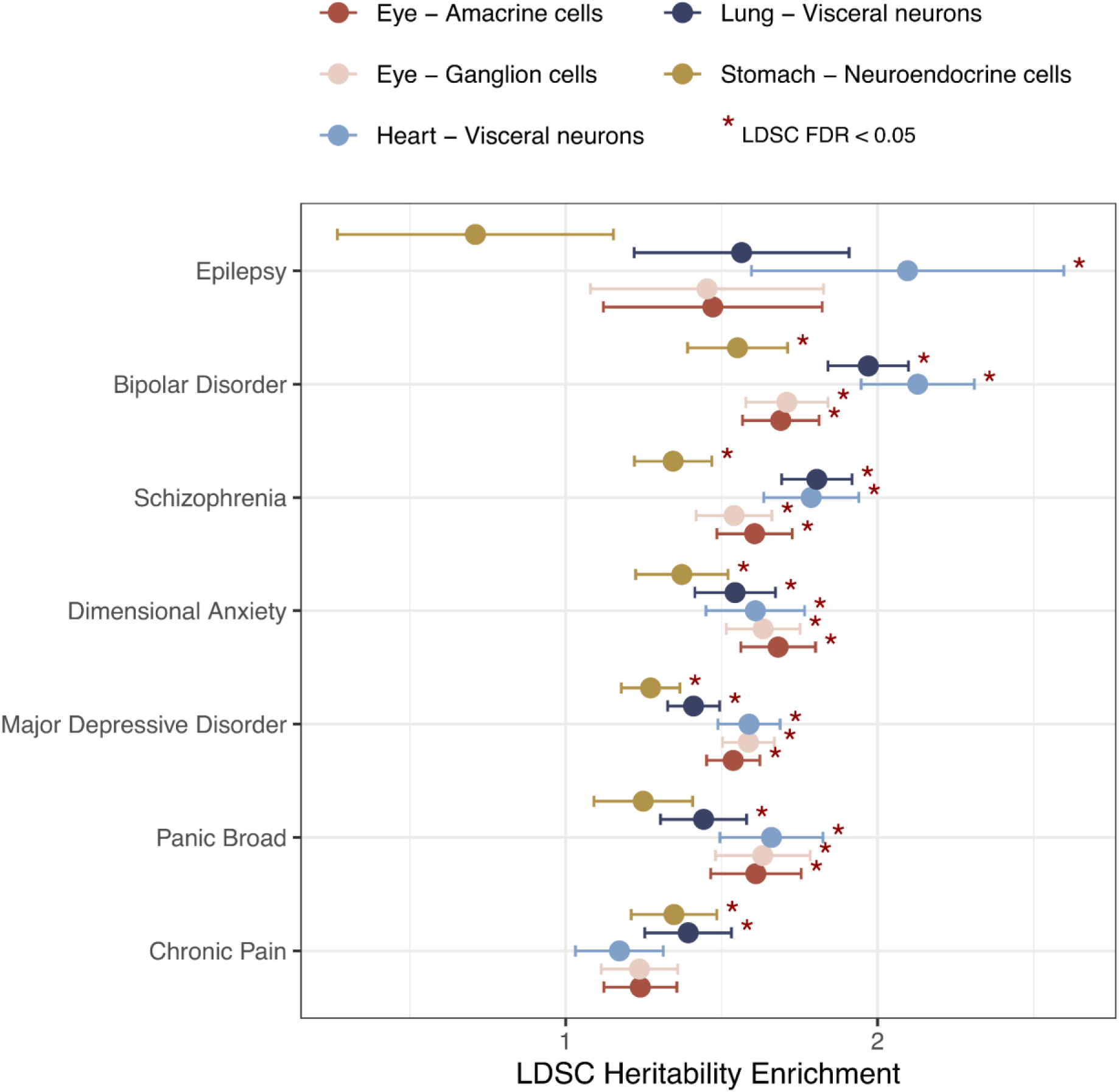
s-LDSC enrichment of psychiatric and neurological disorders across selected CNS and PNS neurons: GWAS of epilepsy^46^, bipolar disorde^41^ schizophrenia ^40^, dimensional anxiety^39^, major depression^29^, and chronic pain^38^. The red asterisk denotes FDR-adjusted significant associations, and the error bars give the enrichment standard error. Bars are ordered from top to bottom legend labels.

### Drug target enrichment

To identify potential repurposing of existing drugs and drug classes for panic broad and disorder (not panic attacks, due to power), we used the DrugTargetor method^47^ (Supplementary Tables 15-18). Among 163 drug classes, two were significantly associated with panic broad and disorder (Bonferroni-corrected p<0.05): antipsychotics and psycholeptics (including anxiolytics such as benzodiazepines, antipsychotics, hypnotics and sedatives). Most of these have been used or suggested for panic treatment^48^. No individual drug compounds were significant in either phenotype.

### Genetic correlations with psychiatric and physical traits

Genetic correlations between the GWAS for panic-related phenotypes and controls screened for panic alone (simple screened) and 104 other psychiatric and physical traits were estimated using LDSC^49^ (Figure 5; full results for simple screening are in Supplementary Tables 19-21). The less strict control screening strategy was adopted for this analysis to avoid upwards bias in the genetic correlations with other psychiatric traits^50^. Across the simple screened versions of panic broad, disorder and attacks, we observed significant positive associations with anxiety disorders (r_g_=0.80-0.92), major depressive disorder (r_g_=0.77-0.79), PTSD symptoms (r_g_=0.55-0.64), and neuroticism (r_g_=0.53-0.56). All three phenotypes also showed positive genetic correlations with coronary artery disease (r_g_=0.15-0.25); blood pressure also had positive correlations but these were much weaker (Figure 5; full results super screening Supplementary Tables 22-24). Using LDSC block-jackknife, we identified 13 unique external traits with significantly different genetic correlations between panic phenotypes that were either super or simple screened (Supplementary Table 25)^49,51^. Across the three pairwise comparisons for simple screened phenotypes, 9 traits differed between panic attacks and panic broad, 8 between panic attacks and panic disorder, and 7 between panic disorder and panic broad. These were predominantly socioeconomic status/cognition and substance-use related traits where panic disorder and panic broad showed significantly stronger genetic correlations than panic attacks. The largest differences were for opioid use disorder (attacks-r_g_=0.10; disorder-r_g_=0.37; |Δr_g_| from unrounded estimates=0.28; jackknife p=4.85×10^-6^) and educational attainment (attacks-r_g_=0.00; disorder-r_g_=-0.24; |Δr_g_|=0.25; p=7.78×10^-19^).

**Figure 5.**
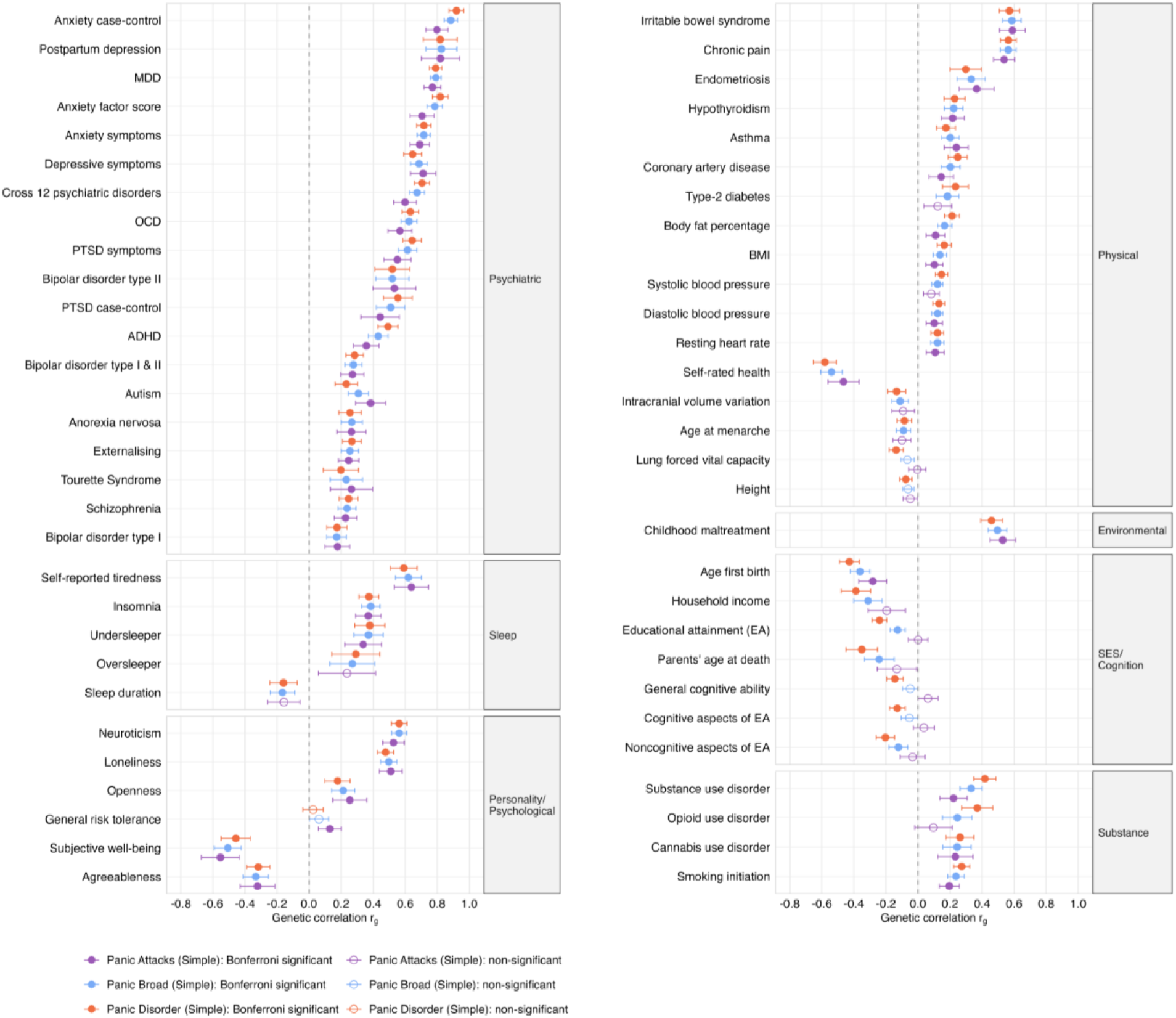
Genetic correlations between panic phenotypes with other traits and disorders, estimated using LDSC. Three panic phenotypes (panic broad, panic disorder, and panic attacks) with simple screened controls were tested for genetic correlations with 104 external traits. Filled shapes: r_g_ significantly different from 0 (Bonferroni-corrected P ≤ 4.81 × 10⁻⁴); hollow shapes: not significantly different from 0. Error bars represent 95% confidence intervals.MDD=major depressive disorder, OCD=obsessive-compulsive disorder, PTSD=post-traumatic stress disorder, OCD = obsessive-compulsive disorder, ADHD=attention deficit hyperactivity disorder, BMI=body mass index.

### Polygenic score prediction and transferability across ancestries

We assessed polygenic scores (PGS) derived from the panic meta-analyses in four independent target samples: two of European ancestry (QIMR Adult Twins, and TEDS via a leave-one-out meta-analysis) and two non-European samples from UK Biobank (African and South-Asian ancestry; Supplementary Table 26). Variance explained is reported on the liability scale, assuming population prevalences of 28%, 5% and 23% for panic broad, panic disorder and panic attacks respectively, with discrimination summarised by the area under the receiver-operating curve (AUC).

In both European-ancestry samples, all three phenotypes were significantly predicted in the expected direction. The panic disorder PGS performed best, explaining 3.18% of liability-scale variance (95% CI 1.72–4.64%; AUC 0.61; P=8.9×10⁻¹⁶) in QIMR Adult Twins and 5.03% (2.64–7.42%; AUC 0.61; P=6.2×10⁻¹⁰) in TEDS. The panic broad PGS explained 1.12% (0.45–1.79%; AUC 0.55; P=9.9×10⁻⁸) in QIMR and 7.05% (5.08–9.02%; AUC 0.61; P=5.3×10⁻¹³) in TEDS, and the panic attacks PGS 0.58% (0.05–1.11%; AUC 0.52; P=0.033) and 4.85% (2.86–6.84%; AUC 0.59; P=8.7×10⁻⁸), respectively.

Cross-ancestry transferability was limited and phenotype-dependent. In the South-Asian sample, the panic broad PGS explained 10.86% of liability-scale variance (95% CI 4.16–17.56%; AUC 0.62; P=1.2×10⁻⁶) and the panic attacks PGS 5.48% (0.22–10.74%; AUC 0.60; P=6.8×10⁻⁴), though the wide confidence intervals reflect the small sample (N≈288). In the African sample, only the panic attacks PGS reached nominal significance (1.49%, −1.37–4.35%; AUC 0.58; P=0.034). The panic disorder PGS did not transfer to either non-European sample (African 0.06%, P=0.70; South-Asian 0.04%, P=0.79).

## Discussion

We report the first genome-wide significant loci for panic attacks and the largest meta-analysis of panic disorder, identifying 25 loci for panic broad, 15 for panic disorder, and 5 for panic attacks, where earlier studies at effective sample sizes below 3,400 found none^15,16^. These loci implicate specific biology, and two features of that biology are prominent: panic shares its common-variant architecture with the other internalising and major psychiatric disorders, which we expected, and a significant part of the shared signal concentrates in visceral and peripheral neuronal populations that psychiatric genetics has rarely examined, which we did not. The genetic overlap itself is to be expected; cross-disorder pleiotropy across the major psychiatric diagnoses is well documented^52^, and our genetic correlations place panic firmly among the internalising disorders alongside anxiety, depression, and post-traumatic stress. Nor is the link between mental illness and the body a new idea. Kraepelin recorded autonomic disturbance in psychosis in 1899^53^, the excess of cardiac and pulmonary disease in schizophrenia and mood disorders has been characterised for decades ^54^, and reduced heart rate variability and central autonomic dysfunction are established in bipolar disorder and depression^55,56^. That literature has read the body largely as a downstream target of central autonomic output, or as a consequence of medication, smoking, and deprivation. What our data add is a hypothetical aetiological claim: common-genetic variant risk for panic is enriched in the neurons of the heart, lung, and gut themselves and may underlie a significant fraction of these body dysfunctions, and this enrichment recurs across bipolar disorder and schizophrenia rather than being specific to panic.

Common-variant heritability, estimated within our largest single cohort from raw genotype data, was 16-27% for panic broad and 10-20% for panic disorder. Raw-data methods applied within one cohort give larger, more reliable heritability estimates than summary-statistic estimators ^24^. The corresponding summary statistic SBayesRC estimates from the meta-analysis were 9 to 12% for panic broad, 6 to 10% for panic disorder, and 4 to 7% for panic attacks. These values place panic near major depression ^27^ and dimensional anxiety ^57^, and below schizophrenia ^40^ and bipolar disorder ^41^, which fits its position in the internalising spectrum.

Panic broad and panic disorder were genetically indistinguishable (r_g_ = 1.00, SE = 0.007), whereas panic attacks diverged from both (r_g_ = 0.95 and 0.91, each below unity). Isolated attacks carry genetic variation the disorder does not, matching the clinical separation between a discrete autonomic surge and a disorder built on recurrent attacks, anticipatory worry, and avoidance ^6^. All three phenotypes correlated strongly with anxiety disorders, major depression, post-traumatic stress, and neuroticism, and modestly with coronary artery disease. Panic disorder correlated more strongly than panic attacks with opioid use disorder and inversely with educational attainment. This suggests that the genetic architecture of panic disorder overlaps more with substance use and cognitive/socioeconomic traits than that of isolated panic attacks. We estimated these correlations from simple-screened controls to avoid inflating correlations with related psychiatric traits ^50^. That panic disorder, but not isolated panic attacks, shows appreciable genetic correlation with opioid use disorder and an inverse correlation with educational attainment indicates that the genetic liability to the disorder extends beyond that of the acute attack, encompassing factors linked to chronicity, comorbidity, and socioeconomic adversity. Together with the departure of the panic-attacks genetic correlation from unity, this argues against treating panic disorder as simply recurrent panic attacks at the genetic level, and supports analysing the two constructs separately.

One locus reached significance across all three phenotypes and both screening strategies: *NEGR1* at 1p31.1, the top panic broad signal and a recurrent association in depression GWAS^27^. Along with *SLC30A9, NEGR1* also emerged as the most consistent gene across expression, splicing, protein, and methylation data in summary-based Mendelian randomization ^58,59^, specially through protein abundance, which marks it for functional follow-up. Of 39 independent significant variants, 34 had prior associations with neuroticism, anxiety, or depression ^18,26–28^; five loci were new to psychiatric genetics, at *MAN1A2*, an MHC signal, *TBC1D32/GJA1*, *HIBADH/TAX1BP1/JAZF1*, and a locus on chromosome 22. Functionally informed fine-mapping ^60,61^ prioritised *PAX6*, *KLC1*, and *GAL3ST1* for panic attacks, genes governing neurogenesis, axonal transport, and myelination.

The single-cell results are the most notable biological finding beyond the loci. Enrichment for panic broad implicated central neurons across cerebrum and cerebellum alongside peripheral visceral neurons of the heart and lung, enteric neurons of the stomach, and several retinal populations. However, the retinal enrichment for panic broad of eye bipolar cells was not replicated in an independent single-cell dataset, though the limited validation of that dataset means this neither confirms nor excludes a retinal contribution. These enrichments fit longstanding physiological accounts of panic centred on cardiac and respiratory afferents ^8,33,42–44^. However, most prior psychiatric single-cell work restricted its reference panels to the central nervous system and set peripheral and visceral neurons aside by construction. We retained these populations because the panic phenotype, dominated by cardiac and respiratory symptoms, warranted their inclusion; doing so revealed associations that a CNS-only design cannot detect. This raises a possibility we state as hypothesis rather than result: if part of panic pathology sits in peripheral neurons, it may be more accessible to systemic drugs than central pathology, and so more druggable. The hypothesis follows directly from the phenotype and would not have surfaced from a brain-only analysis.

Two observations qualify this without weakening it. First, peripheral neurons showed no specific enrichment relative to central neurons (FDR = 0.95), so we cannot exclude that a neuronal expression programme shared between compartments drives much of the signal. Second, across psychiatric disorders, the same visceral and retinal enrichments appeared, often more strongly, in bipolar disorder ^41^ and schizophrenia ^40^ by s-LDSC ^45^ (lung visceral neurons: bipolar 2.0, schizophrenia 1.8, panic broad 1.4). Neither observation is surprising. The nervous system runs throughout the body and shares molecular machinery across its compartments, and most psychiatric disorders have somatic expression that a brain-only lens misses. That these neuronal populations are enriched for genetic associations across several disorders makes them more compelling to study, not less.

Drug-target enrichment ^47^ recovered two classes for panic broad and disorder, psycholeptics and antipsychotics, both already used in panic ^48^; however, no individual compound reached significance. Polygenic scores predicted panic out of sample, with the panic disorder score explaining 3.2% (QIMR Adult Twins) and 5.0% (TEDS) of liability-scale variance in independent European-ancestry cohorts. Transferability to non-European samples was limited and phenotype-dependent: panic broad and panic attacks scores were significant in the South-Asian sample and panic attacks in the African sample, while panic disorder did not transfer, consistent with attenuated portability of European-derived weights and the need for larger discovery efforts in diverse ancestries [62].^22^.

Several priorities follow from these findings. Functional characterisation of the peripheral neuronal enrichment in adult rather than foetal tissue is required to establish whether these populations contribute to panic pathophysiology in the mature nervous system. Discovery in ancestrally diverse cohorts, together with deeper phenotyping through diagnostic phenotyping, would enable improved locus discovery and the genetic separation of panic attacks from panic disorder. Furthermore, sex-stratified analyses are essential to disentangle whether the identified signals reflect a shared or sex-dimorphic genetic architecture. Finally, the hypothesis that peripheral pathology is more accessible to systemically administered drugs than central pathology is directly testable and, if supported, could inform therapeutic development.

Three limitations bound these conclusions. Phenotyping rested on validated self-report rather than clinical interview; discovery was in European ancestry; and the panic attacks GWAS, at five loci, was underpowered for most secondary analyses. The peripheral neuronal enrichments draw on foetal reference data ^32^ and need functional validation. Within those bounds, we place panic in the common-variant architecture of the internalising disorders, and show that the biology these loci flag reaches beyond the brain. Studying panic, defined by what the body does during an attack, led us to find associations of peripheral neuronal populations that matter across psychiatric disorders.

## Online methods

### Participants and phenotypes

This meta-analysis integrated data from seven international cohorts: Australian Genetics of Depression Study, All of Us, Australian Genetics of Bipolar Study, Genetic Links to Anxiety and Depression Study and NIHR BioResource, UK Biobank, Lifelines and Twins Early Development Study. All cohort participants used in these GWAS were of European ancestry. The cohorts primarily used the panic module of the Composite International Diagnostic Interview Short Form (CIDI-SF ^19^), a validated self-report questionnaire based on DSM-5 diagnostic criteria. We defined three phenotypes in this study: panic broad (panic disorder or recurrent panic attacks), panic disorder, and isolated panic attacks (individuals experiencing panic attacks but not panic disorder). Cases were defined as individuals who met lifetime diagnostic criteria for panic attacks or panic disorder or, if diagnostic data were unavailable, who self-reported a panic disorder clinical diagnosis based on a single-item question. Controls were individuals without a history of panic attacks, panic disorder, or other anxiety and mood disorders (super screened controls). To estimate bias as a result of our control definition ^50^, we also created versions of our phenotypes where controls were only screened for panic (simple screened controls), and used these specifically for genetic correlation analyses.

The two biobank cohorts (UK Biobank and All of Us) had additional medical record diagnostic data available that was incorporated in the phenotype definition for these cohorts. Within the UK Biobank, primary (GP READ) and secondary (ICD-10) medical records were additionally used but over 75% of the cases came from the diagnostic questionnaire (Supplementary Table 27). The All of Us Research Program additionally contributed panic disorder cases as ascertained through medical record (SNOMED) codes (concept ID 436074) and control definitions applied as for the other cohorts, including additional screening of depression and anxiety diagnoses using relevant SNOMED codes (Supplementary Materials). Further details, including cohort-specific inclusion and exclusion criteria, are in the Supplementary Material and Supplementary Table 1.

For the panic broad and panic disorder GWAS, these were combined with published panic disorder summary statistics from an existing panic disorder GWAS in Europeans by Forstner et al. (2021). The Forstner-inclusive analyses comprised 90,840 panic broad cases, 41,721 panic disorder cases and 201,638 controls of European ancestry. The panic attacks phenotype (49,119 cases, 192,885 controls) did not incorporate the Forstner data.

### Genotyping and quality control

Each cohort performed genotyping using microarray platforms (or whole genome sequencing in the case of All of Us) and applied stringent quality control procedures to ensure data integrity. Imputation was performed using reference panels (e.g., Haplotype Reference Consortium 1.1 ^62^, 1000 Genomes Project ^63^) to expand genotype coverage. Imputation reference panel, λGC, LDSC intercept and effective sample size for each cohort are given in Supplementary Table 1; per-cohort attenuation ratios are reported in Supplementary Table 1.

### Genome-wide association studies and meta-analysis

Genome-wide association analyses were conducted within each cohort using linear mixed regression models, adjusting for principal components of ancestry and other cohort relevant covariates (see Supplementary Materials). Prior to meta-analysis, quality control was conducted on each cohort’s summary statistics, including filtering variants based on allele frequency (MAF>1%), imputation quality (R2>0.6), and Hardy-Weinberg disequilibrium to exclude low-confidence variants (p < 1 × 10⁻⁶). Summary statistics were then meta-analyzed using a fixed-effects meta-analysis in METAL^21^. SNPs present in >50% of the effective sample size and more than half the cohorts were retained for downstream analyses. Between-study heterogeneity at each variant was assessed using METAL to ensure our associated loci were not driven from cohort heterogeneity. For each variant, METAL performs a second pass over the per-study summary statistics and computes Cochran’s Q statistic (reported as HetChiSq) on k − 1 degrees of freedom, where k is the number of contributing studies, together with its associated p-value (HetPVal) and the I² statistic (HetISq), which quantifies the percentage of variation across studies attributable to heterogeneity rather than chance. Per-variant heterogeneity statistics for genome-wide significant loci are reported in Supplementary Table 2. Genome-wide heterogeneity (median I², proportion of variants with I² > 50%, and proportion with Cochran’s Q p-value < 0.05) was additionally calculated to quantify the proportion of observed variance in effect sizes attributable to true heterogeneity rather than sampling error.

Multi-SNP-based conditional and joint association (GCTA-COJO; v1.94.1) was used to identify significant independent SNPs, within 10Mb windows and with genome-wide significance set at the typical threshold of p < 5x10^-8^.

### Functional Annotation and Pathway Analysis

FUMA (v2.0.0) was used to perform gene-based and gene-set analyses using MAGMA (v.1.08), xQTL and chromatin interaction analyses. These provide insight into potential biological mechanisms underlying the genetic associations.

#### MAGMA gene-based analyses

SNPs were mapped to 19886 protein-coding genes. Genome-wide significance was defined at P=0.05/19886=2.514x10^-6^.

#### MAGMA gene-set analyses

There were 17012 gene sets in the analysis; 6494 curated gene sets (c2.all) and 10529 GO terms (c5.bp, c5.cc and c5.mf) from MsigDB (v2023.1.Hs) were tested in FUMA (v2.0.0). Bonferroni correction was performed for all tested gene sets.

#### Gene property analysis for tissue specificity

To identify tissue specificity of the phenotype, MAGMA gene-property analyses test relationships between tissue-specific gene expression profiles and disease-gene associations. GTEx v8 (54 specific tissue types and 30 general tissue types) and BrainSpan (human brain samples taken at 29 different ages and 11 general developmental stages) data sets were used.

#### Quantitative Trait Loci analyses (xQTLs)

Lead variants and all significant SNPs in LD in each locus were functionally annotated using FUMA xQTLs, which maps variants to genes through evidence from multiple datasets. As per the defaults in FUMA v2.0.0, only significant associations are used, and where these were unavailable (single-cell eQTL datasets “bryois2022Brain” and “jerber2021Dopaminergic”), a default threshold of p < 1x10^-3^ was set.

##### Expression Quantitative Trait Loci (eQTL) analyses

Variants are associated with the total level of expression (mRNA) of a gene in a given tissue. Datasets used were: Brain (5 regions from MetaBrain and 13 from GTEx v10) and Nerve (GTEx v10 - Nerve Tibial) tissues.

##### Splicing Quantitative Trait Loci (sQTL) analyses

Variants are associated with changes in splicing resulting in different combinations and therefore different versions of a protein from the same gene, but the amount of expression (mRNA) is unchanged. The datasets used were Brain (13 from GTEx v10) and Nerve (GTEx v10 - Nerve Tibial) tissues.

##### Protein Quantitative Trait Loci (pQTL) analyses

Variants are associated with protein levels. Datasets used were Brain (“12_yang_2021” 1 set of multi-tissues) and cerebrospinal fluid (“12_yang_2021” 1 set of multi-tissues).

##### Single-cell expression Quantitative Trait Loci (sce-QTL) analyses

Variants are associated with gene expression within a particular cell type, rather than averaged expression across all cells in a tissue as per eQTL analysis. The datasets used were: Brain (bryois2022Brain 8 cell types, jerber2021Dopaminergic 15 cell types, singlebrain 36 cell types, brainscope 17 cell types).

#### Chromatin interaction mapping

3D Chromatin Interaction mapping was performed in FUMA using built-in chromatin interaction data from PsychENCODE (EP links one-way and promoter anchored loops) and Hi-C brain tissue data (adult cortex, fetal cortex, dorso-lateral prefrontal cortex, hippocampus, neural progenitor cell) from GSE87112. FDR threshold of significance was set as p<1x10^-6^.

### Fine-mapping

To identify putative causal SNPs, functionally-informed per-SNP prior causal probabilities were derived in PolyFun (1.0.0) using the baseline-LF 2.2 model, with linkage disequilibrium (LD) scores from the UK Biobank European reference panel ^60^. Fine-mapping was performed in SuSiE (0.11.92)^61^, limiting to variants with MAF≥0.01 and INFO≥0.6, assuming one causal SNP per locus, and using windows defined from independent LD blocks in European ancestry individuals from the 1000 Genomes project ^64^. A posterior inclusion probability (PIP) cutoff of ≥0.95 was used to identify putative causal SNPs. Credible causal sets were defined as the minimum set of ranked variants with cumulative PIP≥0.95. We did not apply the minimum pairwise r^2^ criterion for credible causal sets from SuSiE, as the PolyFun + SuSiE pipeline only uses LD estimates for the LDSC step, not for fine-mapping when a single causal variant is specified.

### Summary-data-based Mendelian Randomization

To further prioritize candidate genes underlying GWAS risk loci for downstream functional validation, we performed Summary-data-based Mendelian Randomization (SMR) and Heterogeneity in Dependent Instruments (HEIDI) analyses using the SMR Portal under default parameters (v1.4.1) ^58,59^. These analyses tested whether the observed associations between genetic variants and the panic phenotypes were mediated by molecular traits (gene expression, alternative splicing, DNA methylation, and protein expression) via pleiotropy. For the eQTL and sQTL analyses, we integrated brain and tibial nerve molecular datasets, including BrainMeta, 13 brain subregions from GTEx (v10), and GTEx tibial nerve data (v10), while mQTL analyses were conducted using brainMeta data. For pQTL analyses, we used blood-based data from the INTERVAL, FENLAND, and SCALLOP consortia, as brain-based data was not available in the SMR Portal. The significance threshold for the SMR analysis was set using a strict Bonferroni correction (P < 0.05 /number of genes tested within each molecular dataset). A significant SMR result can indicate either a pleiotropic model, or a linkage model; therefore, the HEIDI test was conducted to distinguish between these two scenarios. Only genes with a non-significant HEIDI test result (p > 0.05) were retained, supporting a pleiotropic rather than a linkage mechanism.

### Single-cell analyses

#### Reference datasets

The cell-by-gene matrices for all single-cell RNA-Seq studies were sourced from CZ CELLxGENE ^65^ or previously processed for MAGMA Celltyping analysis ^30^. These were the human foetal cell atlas ^32^, the mouse CNS atlas ^34^, the human lung cell atlas ^36^, the human adult and foetal epicardium atlas ^35^ and the human retina developmental cell atlas ^37^. All single-cell RNA-Seq datasets were processed using the EWCE R package (v1.13.1) ^66^ to first remove unexpressed, non-varying or non-differentially expressed genes across cell types. Secondly, the genes’ mean expression and specificity for each cell type were calculated. For each cell type, specificity was calculated by dividing the expression of each gene by the sum of expression for that gene across all cell types. Genes from the mouse CNS single-cell atlas were first mapped to human based on one-to-one orthologs using the Orthogene R package, where genes without a one-to-one mouse:human ortholog were dropped ^67^.

#### Enrichment analyses

Single-cell enrichment analyses were conducted using Generalized Gene-Set Analysis of GWAS Data (MAGMA) and stratified LD Score Regression (s-LDSC). The MAGMA single-cell enrichment analysis was conducted using the “*MAGMA.Celltyping*” R package (v2.0.14) with results shown for both the linear regression (Linear) and the top 10% of genes based on specificity (Top 10%) approaches ^30^. *MAGMA.Celltyping* uses MAGMA along with gene-by-cell type specificity matrices to measure the enrichment of GWAS signals in each cell type. MAGMA (v1.10.0) ^68^ was used to first map SNPs to genes, with a maximum window of 35k bp upstream and 10k bp downstream, followed by the cell type association analysis. All enrichment tests were corrected for multiple testing with the Benjamini-Hochberg approach and a false discovery rate (FDR) threshold of 0.05.

The s-LDSC single-cell enrichment analysis was conducted, as previously described ^30^, by creating a gene set of the top 10% of genes per cell type by specificity. These genes were used to create bed files with their coordinates extended by 10 kb upstream and 1.5 kb downstream of the gene body in a strand-specific manner. s-LDSC quantifies the contribution of polygenicity under the assumption that genetic effect values for true associations are positively correlated with LD scores since SNPs with higher LD scores tend to have higher chi-squared statistics, i.e. the strength of association of a SNP with a trait ^45^. Thus, s-LDSC can be used to quantify whether a set of genomic loci, containing a specific category of SNPs, contributes more or less to heritability than expected given the proportion of SNPs in the genomic set. This category of SNPs is chosen if their physical location is within the boundaries of the set of genomic loci, which in our analysis is 15k bp upstream and 1.5k bp downstream of the cell type-specific gene bodies. And the heritability of a SNP is quantified as how many SNPs it is in LD with, within the chosen genomic loci (i.e. the LD score). This heritability enrichment is relative to the number of contributing SNPs relative to all SNPs. For example, if we took all coding regions as the genomic loci of interest, and if coding SNPs make up 10% of all SNPs but explain 30% of heritability, the enrichment would be .3/.1=3, meaning they contribute 3 times more to heritability than expected based on their frequency.

In this manner, s-LDSC (v1.0.1) ^45^ was used to test for heritability enrichment. Specifically, annotation files for each cell type’s gene bed files were first created with Phase 3 of the 1000 genomes reference. Followed by the generation of LD scores with a window size of 1 centiMorgan (cM) i.e. approximately 1 million base pairs, filtering to HapMap3 SNPs to match the baseline model. Finally, the enrichment analysis was run for the GWAS summary statistics across the different cell types and those in the baseline model whilst excluding the major histocompatibility complex (MHC) (due to the known difficulties predicting LD in this region). The Benjamini-Hochberg approach was used for multiple test correction with a threshold of 0.05, and the heritability enrichment (Proportion heritability / Proportion SNPs, as described above) was reported.

#### Differential gene single-cell enrichment analysis

The differential gene enrichment analysis was conducted by first identifying differentially expressed genes for the differing cell type groups in the human foetal cell atlas ^32^, following the approach previously outlined ^69^. In short, reads for each gene were pseudobulked; aggregated by summing, for each foetal sample which helps reduce the dropout issue in single-cell data and avoids the false inflation of confidence from non-independent samples of pseudoreplication approaches ^70,71^. The differential analysis was conducted using EdgeR LRT approach ^72^, from the EdgeR R package (v 4.0.16). Differentially expressed genes were defined by a Benjamini-Hochberg multiple test corrected p-value threshold of 0.05 and subsetted to those upregulated in the cell type group of interest. The resulting genes were passed to MAGMA for gene set enrichment analysis (v1.10.0) ^68^ with a maximum window of 35k bp upstream and 10k bp downstream, in the panic broad Panic Attack GWAS summary statistics set.

#### GWAS summary statistics processing

Other psychiatric disorder GWAS were evaluated for cell type enrichment analysis as part of this study, including chronic pain ^38^, dimensional anxiety ^39^, schizophrenia ^40^, major depressive disorder – European ancestry, non-23andMe ^29^, and bipolar disorder – European ancestry, non-23andMe ^41^.

All GWAS summary statistics were uniformly processed using the MungeSumstats R package (v1.15.0), with the ‘format_sumstats’ function and default parameters with dbSNP v155 ^73^ and converting build to GRCh37 where necessary for cell typing analysis.

### SNP-based heritability and genetic correlations

Summary statistic SNP-based heritability estimates for panic attacks and panic disorder were calculated using SBayesRC^22^ in GCTB (v2.5.2). These estimates quantify the proportion of phenotypic variance explained by common genetic variants. To account for differences in population prevalence, heritability estimates were converted to the liability scale ^74^ across a range of population prevalence estimates, with probable ranges deemed as 5-25% for panic broad, 1–10% for panic disorder and 5–30% for panic attacks.

We used LDSC (v.1.0.10) to estimate genetic correlations between i) each of the panic phenotypes and ii) each panic phenotype with 104 other psychiatric and physical traits ^49^. Internal genetic correlations between panic phenotypes were tested for a significant difference from 0 (default LDSC z-test) and from 1 (chi-squared test applied to [(|rg|−1)/SE]²), with Bonferroni-corrected significance threshold P ≤ 5.56×10⁻³ (0.05/9 internal pairs tested). The 104 external traits were selected from our internal database of GWAS summary statistics and were considered sufficiently powered based on LDSC SNP-based heritability Z-scores exceeding 4. These comparator datasets comprised publicly available and consortium summary statistics, including the Million Veteran Program anxiety GWAS; the Million Veteran Program data were used solely as a comparison trait in genetic correlation analyses and did not contribute to the panic meta-analysis. Statistical significance for external trait genetic correlations with each panic phenotype was determined using a Bonferroni-corrected P-value threshold of 4.81x10^-4^ (0.05/104 external traits tested). We used the block-jackknife LDSC method to assess whether the genetic correlation with each external trait differed significantly (p < 4.81x10^-4^) between the panic phenotypes ^49,51^. LDSC analyses were conducted using HapMap3 SNPs, MAF ≥ 0.01, INFO ≥ 0.6, and with LD scores derived from the European 1000 Genomes Project reference panel.

### Polygenic scoring

Polygenic scores (PGS) for panic broad, panic disorder, and panic attacks were calculated to characterize variance explained across different ancestries ^22^. PGS were calculated in an independent target sample of European ancestry from the Adult Twin cohort at QIMR Berghofer, where only one twin per family was retained in the analysis (N=3,727 unrelated). Panic phenotyping in this cohort was determined using self-reported diagnosis/experiences of panic attacks ^75^ as previously described. To compare this to a more stringent, diagnostic phenotyping, PGS were also computed for individuals of European ancestry in TEDS - using leave one-sample-out GWASs.

To test the transferability of our meta-analysis to other ancestries, African and South-Asian ancestry PGSs were calculated in the UK Biobank using a similar diagnostic approach as used in the meta-analysis - with the exception of the use of GP codes which were not accessible on the UKB RAP at time of submission. We excluded related participants with a kinship coefficient > 0.0422 as provided by the UKB, with controls preferentially removed (see Supplementary Table 26 for sample sizes). Although there is a pre-existing GWAS of panic disorder in individuals of East-Asian ancestry ^75^, these summary statistics were not available.

PGS were calculated using SBayesRC in GCTB (v2.5.2) ^22^, a Bayesian regression framework that accounts for linkage disequilibrium (LD) among SNPs and integrates functional annotation data to refine effect size estimates. The analyses incorporated the pre-computed LD eigen-decomposition data derived from UK Biobank participants of European ancestry (7 million imputed SNPs) alongside per-SNP functional annotation files covering the same set of variants that are provided by the SBayesRC developers. In our European analysis, the Markov chain was run for 21,000 iterations with the first 1,000 discarded as burn-in, with all remaining parameters left at their default values. In the trans-ancestry analysis, the Markov chain was run for 5000 iterations with a burn in of 1,000. Posterior SNP effect sizes estimated by SBayesRC were then used to calculate PGS for each participant using the individual genotype data and the PLINK (v1.9) –score function. Within each target cohort, PGS were standardised to aid interpretation of effect sizes.

We conducted logistic regressions to assess the variance explained by PGS for each panic phenotype across ancestries (i.e., Nagelkerke R²), converting the results into liability R² from Lee 2011 ^76^ with an additional error term included ^74^, and assuming a prevalence of 28% for panic broad, 5% for panic disorder, and 23% for panic attacks ^2^.

In the European target sample, the first 4 genetic principal components and batch were included as covariates. In the UK Biobank African- and South-Asian-ancestry target samples, the first 10 genetic principal components as provided by the UKB, assessment centre and genotyping batch were included as covariates.

Receiver operating characteristic analyses were also conducted. To test whether the AUC >0.5, Z-scores were calculated with Equation (1), where x = AUC, and H0 = 0.5. One-tailed p-values were obtained from the standard normal distribution and adjusted for nine comparisons.

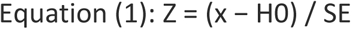

### Drug Targetor

Drug Targetor (Nov 2020 db release) tested the association of drug-related gene sets and drug class enrichment. DrugTargetor collected drug-gene interactions from two databases ChEMBL ^77,78^ and DGIdb ^79^. The analysis used the same settings as Strom et al. ^18^, including the hypothesis of drug action for the nervous system, and 1500 maximum unique drugs and 200 maximum classes of drugs. MAGMA version 1.10 was applied to prioritise associated genes using gene flanks of −35kb 5’ and +10kb 3’ ^68^. The area under the enrichment curve and associated p-value from one-sided Mann–Whitney–Wilcoxon tests were used to assess the enrichment of drug classes ^80^.

## Acknowledgments

**AGDS/QSkin:** We are indebted to the participants for giving their time to contribute to this study. We thank all the people who helped in the conception, implementation, beta testing, media campaign, and data cleaning. The Australian Genetics of Depression Study was funded by grant 108663 from the Australian National Health and Medical Research Council (NHMRC). This work was supported by NHMRC Investigator Grants to BLM (2017176); NRW (1173790); NGM (1172990); and IBH (2016346). The QSkin Study is supported by an NHMRC Clinical Trials and Cohort Grant [APP1185416]. D.C.W. is supported by a NHMRC Investigator grant (APP1155413). EMB received funding from the University of Queensland Health Research Accelerator Program

**GLAD+**: We thank NIHR BioResource volunteers for their participation, and gratefully acknowledge NIHR BioResource centres, NHS Trusts and staff for their contribution. We thank the National Institute for Health and Care Research, NHS Blood and Transplant, and Health Data Research UK as part of the Digital Innovation Hub Programme. The views expressed are those of the author(s) and not necessarily those of the NHS, the NIHR or the Department of Health and Social Care.

We gratefully acknowledge the participation of the NIHR BioResource Centre Maudsley, Biomedical Research Centre at South London and Maudsley NHS Foundation Trust and King’s College London volunteers and thank the staff for their help with volunteer recruitment. We thank the NIHR Biomedical Research Centre at South London and the Maudsley NHS Foundation Trust and King’s College London for funding. This study represents independent research supported by the NIHR Biomedical Research Centre BioResource at South London and Maudsley NHS Foundation Trust and King’s College London. We gratefully acknowledge capital equipment funding from the Maudsley Charity (Grant Ref. 980) and Guy’s and St Thomas’s Charity (Grant Ref. STR130505).

**Lifelines Cohort Study:** The Lifelines Biobank initiative has been made possible by funding from the Dutch Ministry of Health, Welfare and Sport, the Dutch Ministry of Economic Affairs, the University Medical Center Groningen (UMCG the Netherlands), University of Groningen and the Northern Provinces of the Netherlands. The generation and management of GWAS genotype data for the Lifelines Cohort Study is supported by the UMCG Genetics Lifelines Initiative (UGLI). UGLI is partly supported by a Spinoza Grant from NWO, awarded to Cisca Wijmenga. The authors wish to acknowledge the services of the Lifelines Cohort Study, the contributing research centers delivering data to Lifelines, and all the study participants.

**MVP:** This study used summary statistics from the Million Veteran Program to conduct genetic correlations with our results. The authors thank Million Veteran Program (MVP) staff, researchers, and volunteers, who have contributed to MVP, and especially participants who previously served their country in the military and now generously agreed to enroll in the study. (See https://www.research.va.gov/mvp/ for more details). The citation for MVP is Gaziano, J.M. et al. Million Veteran Program: A mega-biobank to study genetic influences on health and disease. J Clin Epidemiol 70, 214-23 (2016). This research is based on data from the Million Veteran Program, Office of Research and Development, Veterans Health Administration.

**QIMR GBP**: We thank the participants for giving their time and support for this project. We acknowledge and thank M. Steffens for her generous donations in loving memory of J. Banks. Data collection was funded and data analysis was supported by the Australian National Health and Medical Research Council (No. APP1138514) to S.E.M.. S.E.M. is supported by a National Health and Medical Research Council Investigator Grant (No. APP2025674).

**TEDS**: We gratefully acknowledge the ongoing contribution of the Twins Early Development Study (TEDS) participants and their families. TEDS is funded by a UK Medical Research Council (MRC) programme grant (MR/V012878/1) to TC Eley (previously MR/M021475/1 to R Plomin).

**UK Biobank:** This research has been conducted using the UK Biobank Resource under Application Number 82087. This work uses data provided by patients and collected by the NHS as part of their care and support.

**Additional**: The authors acknowledge the use of the King’s College London research computing facility, CREATE (https://doi.org/10.18742/rnvf-m076). N.S. received funding from a UKRI Future Leaders Fellowship [grant number MR/T04327X/1] and the UK Dementia Research Institute award number UK DRI-5008 through UK DRI Ltd.

## Consortia

**GLAD Study group author:** Gerome Breen, Thalia C. Eley, Matthew Hotopf, Chérie Armour, Ian R. Jones, Andrew M. McIntosh, James T. R. Walters, Daniel J. Smith, Jonathan R. I. Coleman, Allan H. Young, Joshua E. J. Buckman, Antony J. Cleare, Ewan Carr, Katrina A. S. Davis, Kimberly A. Goldsmith, Colette R. Hirsch, Georgina Krebs, Katharine A. Rimes, Evangelos Vassos, David Veale, Janet Wingrove, Roland Zahn, Molly R. Davies, Gursharan Kalsi, Shannon Bristow, Susannah C. B. Curzons, Henry C. Rogers, Katherine N. Thompson, Brett N. Adey, Ian Marsh, Dina Monssen, Monika McAtarsney-Kovacs, Charles J. Curtis, Jahnavi Arora, Saakshi Kakar, Laura Meldrum, Iona Smith, Launch placement students, Le Roy Dowey, Victor Gault, Donald M. Lyall, Ruth K. Price, Keith G. Thomas, Zain Ahmad, Helena L. Davies, Christopher Hübel, Sang Hyuck Lee, Abigail R. ter Kuile, Danyang Li, Yuhao (Leo) Lin, Jessica Mundy, Jared G. Maina, Alish Palmos, Alicia J. Peel, Kirstin Purves, Christopher Rayner, Megan Skelton, Rujia Wang, Johan Zvrskovec

**UMCG Genetics Lifelines Initiative (UGLI) group author: LifeLines Cohort Study** - Raul Aguirre-Gamboa, Patrick Deelen, Lude Franke, Jan A Kuivenhoven, Esteban A Lopera Maya, Ilja M Nolte, Serena Sanna, Harold Snieder, Morris A Swertz, Peter M. Visscher, Judith M Vonk, Cisca Wijmenga, Naomi Wray. See supplemental information for consortium member affiliations.

**NIHR BioResource consortium group author**: John Bradley, Nathalie Kingston, Hannah Stark, Carola Kanz, Alexei Moulton, Nigel Ovington, Jacinta Lee, Debbie Clapham-Riley, Katie Mills

## Data and code availability

For the purposes of open access, the authors have applied a Creative Commons Attribution (CC BY) licence to any Accepted Author Manuscript version arising from this submission.

Summary statistics will be made available upon acceptance from a website such as Figshare or the GWAS catalog. Individual data from each cohort are available to approved researchers, subject to registration and application processes conducted by the study teams. GLAD+ Study data is available via a data request application to the NIHR BioResource (https://bioresource.nihr.ac.uk/using-our-bioresource/academic-and-clinical-researchers/apply-for-bioresource-data/). The data are not publicly available due to restrictions outlined in the study protocol and specified to participants during the consent process. for details of the data freeze including the variables for the analyses described in this paper. Data application processes are detailed in the following links for Lifelines https://www.lifelines-biobank.com/researchers/working-with-us, TEDS https://www.teds.ac.uk/researchers/teds-data-access-policy/, and UK Biobank https://www.ukbiobank.ac.uk/enable-your-research/apply-for-access. Information on how to apply to access the MVP summary statistics used in this analysis can be found here https://www.ncbi.nlm.nih.gov/projects/gap/cgi-bin/study.cgi?study_id=phs001672.v11.p1. The AGDS and GBP data from QIMR Berghofer are not publicly available due to restrictions outlined in the study protocol and participant consent agreements. However, the summary statistics used in this study are available upon request by contacting. Access to raw data requires a formal Data Transfer Agreement.

Project code will be made available from a public repository on GitHub upon acceptance. The FUMA results will be published on the FUMA website^25^.

## References

