## Supplementary Materials for "Genome-wide meta-analysis of panic attacks and disorder identifies genetic risk factors and implicates multiple body systems in panic and other psychiatric disorders"

**for**

### Supplementary Methods

### 1. Cohort Information

#### AGDS: Australian Genetics of Depression Study

The Australian Genetics of Depression Study (AGDS) is a nationwide volunteer cohort of adults aged 18-90 who have been diagnosed with, and/or treated for, depression. Recruitment procedures and characteristics of AGDS are described in a published cohort profile^1^. Participants joined the study after responding to either a nationwide media campaign or a letter from the Australian Government’s Department of Human Services, mailed to 110,000 Australian residents who had received 4 or more antidepressant prescriptions over the prior 4.5 years of any of the 10 most common antidepressants in Australia.

Enrolled participants completed a comprehensive online survey about their physical and mental health. Participants had the option to donate a saliva sample for genetic (DNA) analyses; over 75% of AGDS returned a sample via a mail-out kit. Participants completed the CIDI-SF panic disorder module. Genotyping used the Illumina Global Screening Array V2. Samples were merged with the 1000 Genomes project samples and genetic principal components (PCs) were calculated using a set of single nucleotide polymorphisms (SNPs) not in linkage disequilibrium. Pre-imputation quality control was done using PLINK 1.9^2^, including removing SNPs with a minor allele frequency < 0.005, SNP call rate < 97.5%, and identification of participants with genetic similarity to a European reference group (>4 SD from Ancestry Principal Components [PCs] PC1/PC2 centroid) and Hardy-Weinberg equilibrium (p<1×10^-6^), before imputation using the Haplotype Reference Consortium 1.1 reference panel. The study received ethical approval by the QIMR Berghofer Medical Research Institute Human Research Ethics Committee in Brisbane, Australia. Written informed consent was obtained from participants.

In the AGDS cohort, cases were defined using the CIDI-SF: panic disorder cases met DSM-5 criteria while panic broad cases met criterion A. Due to the recruitment focus of AGDS, participants implicitly had internalising disorders. Therefore, controls were taken from the QSkin cohort, which is an Australian-based cohort focused on skin cancer, with screening questions that include mental health conditions^3^. QSkin controls self-reported having never experienced panic attacks, or receiving a diagnosis of panic disorder, any anxiety or depressive disorder. GWAS were performed using SAIGE (v 1.3.0). Null logistic mixed models were conducted on chromosomes 1 to 23, adjusted for sex and the first four principal components (PCs). For the association of genetic variants on the X chromosome, the genotypes in males were coded as 0/2. Genetic variants with a minor allele frequency less than 1% and an imputation quality score less than 0.6 were excluded from the GWAS results.

#### AoU: All of Us

The All of Us (AoU) Research Program is an ongoing prospective national cohort study in the United States, currently comprising more than 413,000 participants aged 18 years or older. AoU collects a wide range of data, including survey responses, electronic health records, physical measurements, wearable devices, and genomic information^4^. In this analysis, we used whole-genome sequencing (WGS) data generated by the AoU Research Program. WGS libraries were prepared using PCR-free protocols and sequenced on the Illumina NovaSeq 6000 platform. The resulting genomic data underwent standardized quality-control procedures and achieved a mean sequencing coverage of at least 30×. For all analyses in this study, we used the conservative predicted European ancestry assignment (ancestry_pred_other) from AoU, which allows individuals with uncertain ancestry classification to be assigned to the "Other" category.

***Phenotype in AoU***

Panic cases and controls were defined in AoU using Electronic health records (SNOMED codes), self-reported diagnosis and a questionnaire in the Behavioural Health and Personality Questionnaire that assessed panic criteria approximating the DSM-5 criteria. In this, panic attacks were not assessed using the 13 item criterion A, but instead using the description *“The next section asks about panic attacks. Panic attacks are also called anxiety attacks. These are sudden, strong feelings of fear that reach their peak within a few minutes. When a person is having an anxiety attack, they usually have physical symptoms, too. When these symptoms happen, they can be severe and frightening, so much so that people may think they are having a heart attack. Your answers to these next questions may help researchers understand how to better prevent and treat these attacks. “*

**Panic broad cases** answered “yes” to the following question *“With this definition in mind, did you ever in your life have a panic attack?”* (Concept ID: 1703900) or had an Electronic health record of panic disorder (concept ID 436074). A total of 29,060 participants were identified as panic broad cases.

**Panic disorders cases** had an EHR of panic disorder (concept ID 436074) or met the DSM criteria for panic disorder in the questionnaire in Behavioural Health and Personality Questionnaire as below:

Participants reported a lifetime history of panic attacks (concept ID 1703900), at least one unexpected (“out-of-the-blue”) panic attack (1703927), persistent concern about future attacks or behavioral changes following attacks (1703922), and recurrent panic attacks, defined as more than two lifetime attacks (1703899). Participants with missing responses or who selected “Skip” for any criterion were excluded from case classification.

There are 11,952 participants in the AoU who met the criteria of panic disorders and have been used in this study.

**Panic Attack cases** were individuals that experience panic attacks but did not meet panic disorder criteria as above (N= 17,108).

*Controls*

Two control definitions were used. Super screened controls were used in the primary analyses, whereas simple-screened controls were used in sensitivity analyses.

Super screened controls are based on the simple screened control, further screening out participants who have

1.	EHR diagnosis of depression (Concept ID 440383) or

2.	Self-reported depression, defined as endorsing “Self (1384656)” for the question “Including yourself, who in your family has had depression?” or

3.	Meeting criteria for DSM-5 depression based on the AoU mental health questionnaire. Participants were considered to meet the criteria if they endorsed depressed mood (1704045) and/or anhedonia (1703998) and reported at least five depressive symptoms across depressed mood (1704045), anhedonia (1703998), weight/appetite change (1704047 or 1703985), sleep disturbance (1704036), fatigue (1704012), excessive guilt or worthlessness (1703972), impaired concentration (1704025), and thoughts of death (1704136), with symptoms occurring during a period lasting at least two weeks. Weight change and appetite change were combined into a single symptom criterion. or

4.	EHR diagnosis of anxiety disorders (441542) or

5.	Participants were classified as possible GAD cases if they endorsed excessive anxiety or worry for at least six months, defined by either feeling worried/anxious (1704052) or worrying excessively (1703979); reported worry about multiple life domains (1704006); reported difficulty controlling worry (1703982); and endorsed at least two of five available associated symptoms: restlessness or feeling keyed up/on edge (1704050), difficulty concentrating or mind going blank (1704032), irritability (1704053), muscle aches or tension (1704043), and sleep disturbance (1704040). Responses of “Most of the time” or “All or almost all of the time” were coded as symptoms present.

We identified 95,939 super-screened controls based on the described definitions.

Simple screened control European-ancestry participants with sequenced genomic data but without

1.	panic attacks or

2.	panic disorder or

3.	self-reported anxiety or panic to survey question “Including yourself, who in your family has had anxiety reaction/panic disorder? Select all that apply --self (1384475)”, or

4.	self-reported social phobia based on survey “Including yourself, who in your family has had a social phobia? - Self (1384464)”

In total 158,435 participants have been identified as simple controls in this study.

***GWAS in AoU***

In All of Us, variants were quality controlled using PLINK 2.0 by excluding variants with minor allele frequency (MAF) <1%, minor allele count (MAC) <20, genotype missingness >5%, or Hardy–Weinberg equilibrium P-value <1×10⁻¹⁵. Genome-wide association analyses were restricted to genetically inferred European-ancestry participants and performed using REGENIE (v3.2.9) by using logistic regression, adjusted for the first 10 genetic principal components, biospecimen source, and recruitment site indicators. The X chromosome was analyzed assuming complete dosage compensation in males.

#### LL: Lifelines

Lifelines is a prospective population-based cohort study recruiting over 167,000 participants including multi-generation family members in the North of the Netherlands between 2006 and 2013^5^. The second follow-up assessment took place between 2014 and 2017, and the third follow-up assessment between 2019 and 2024. Lifelines employs a broad range of investigative procedures in assessing the biomedical, socio-demographic, behavioral, physical and psychological factors which contribute to the health and disease of the general population, with a special focus on multimorbidity and complex genetics. Among all participants, genome-wide genetic data of over 78,000 participants are available. The Lifelines Cohort study is conducted according to the principles of the Declaration of Helsinki and in accordance with the research code of University Medical Center Groningen, and is approved by its medical ethical committee. All participants signed an informed consent form.

Panic disorder and panic broad were measured using the MINI International Neuropsychiatric Interview (MINI) for adults^6^. The MINI was performed as an individual face-to-face interview by a trained research nurse at baseline when participants visited a Lifelines research facility. During the follow-up, the MINI was administered as a digital questionnaire with participants entering their answers under the supervision of a trained research nurse on location.

Panic disorder was measured using three repeated MINI interviews administered at baseline, second, and third assessments. There are 17 items related to panic disorder in the MINI, and each item elicits a yes/no response which can be mapped to DSM-5 panic disorder criteria. Additionally, the question "Could you indicate which of the following disorders you have (had): panic disorder?" has been asked from baseline and every 1.5 years in follow-up questionnaires, resulting in a total of six repeated measurements of self-reported panic disorder. Panic disorder was defined based on meeting case criteria on the MINI or self-reported 'yes' for panic disorder.

A panic attack was defined as experiencing four or more of the 13 symptoms listed in the MINI (DSM-5 Criterion A). The panic broad phenotype was defined as a "yes" for panic attack on the MINI, self-reported panic disorder.

Controls for both panic disorder and panic broad were categorized as follows: 1) Primary analysis controls: Individuals with no history of panic disorder, panic attacks, depression, or anxiety as per the MINI or self-report items. The MINI used in the Lifelines cohort was restricted to the following diagnoses: major depressive episode, dysthymia, panic disorder, agoraphobia, social phobia and generalized anxiety disorder. 2) Simple screened sensitivity analysis controls: Individuals with no history of panic disorder or panic attacks.

Genome-wide genotyping was available to over 78,000 participants. The first subset of 17,033 participants was genotyped using the Illumina CytoSNP-12v2 array. Pre-imputation quality control was performed in which samples and variants were excluded with a call rate < 95%, as well as variants with Hardy-Weinberg equilibrium (HWE) P<1×10^-4^, or minor allele frequency (MAF) <1%, and samples with a sex mismatch, deviating heterozygosity (>4 SD from the mean) or of non-European ancestry. A total of 15,400 samples and 265,000 SNPs were available for analysis. The second subset of 38,030 participants was genotyped using the Infinium Global Screening Array® (GSA) MultiEthnic Disease Version. Standard quality control was performed on both samples and markers, including removal of samples and variants with a low genotyping call rate (<99%), variants showing deviation from HWE (P<1×10^-6^) or excess of Mendelian errors in families (>1% of the parent-offspring pairs), and samples with a sex mismatch, and very high or low heterozygosity. After quality control, a total of 36,339 samples and 571,420 SNPs were available for analysis. The third subset of 29,166 participants were genotyped using the FinnGen Thermo Fisher Axiom® (AFFY) custom array. Standard quality control was performed on both samples and markers, including removal of samples and variants with a low genotyping call rate (<99%), variants showing deviation from HWE (P<1×10^-6^) or excess of Mendelian errors in families, and samples with a sex mismatch, and very high or low heterozygosity. After quality control, a total of 28,250 samples and 462,731 SNPs on autosomal and X chromosomes were available for analysis. These three genotyping datasets were imputed using the HRC panel v1.1 ^7^ at the Sanger imputation server, and variants with an imputation quality score > 0.4 for variants with a MAF > 0.01 were retained. Finally, 78,581 participants with genetic data were available.

GWASs for panic disorder and panic attack were conducted using REGENIE v3.4.1 across three datasets. For autosomes, the analysis included 10 principal components (PCs) as covariates. For the X chromosome, available only for GSA and AFFY datasets, the analysis included 10 PCs and sex as covariates, and Regenie automatically recodes male genotypes as diploid (0/2) to match the dosage scale of female genotypes.

**UMCG Genetics Lifelines Initiative (UGLI) group author: LifeLines Cohort Study**
Raul Aguirre-Gamboa (1), Patrick Deelen (1), Lude Franke (1), Jan A Kuivenhoven (2), Esteban A Lopera Maya (1), Ilja M Nolte (3), Serena Sanna (1), Harold Snieder (3), Morris A Swertz (1), Peter M. Visscher (3,4), Judith M Vonk (3), Cisca Wijmenga (1), Naomi Wray (4).
1. Department of Genetics, University of Groningen, University Medical Center Groningen, The Netherlands 2. Department of Pediatrics, University of Groningen, University Medical Center Groningen, The Netherlands 3. Department of Epidemiology, University of Groningen, University Medical Center Groningen, The Netherlands 4. Institute for Molecular Bioscience, The University of Queensland, Brisbane, Queensland, Australia.

#### GLAD UKB: GLAD+ and UK Biobank

The NIHR BioResource Genetic Links to Anxiety and Depression (GLAD) Study is an online study of adults living in the UK with lifetime experience of depression and/or anxiety ^8^. Participants have completed extensive online questionnaires on sociodemographics, mental and physical health, and lifestyle. During the COVID-19 pandemic, the GLAD Study research team recontacted GLAD participants and healthy volunteers from other NIHR BioResource (<https://bioresource.nihr.ac.uk/>) studies to conduct the Covid-19 Psychiatry and Neurological Genetics (COPING) study, including >20,000 participants with psychiatric disorders and >11,000 healthy volunteers, two-thirds of whom have been genotyped. “GLAD+” consists of GLAD as well as participants from the COPING study, who completed the same measures but were not recruited on the basis of mental health diagnostic status.

UK Biobank (UKB) is a UK population-based health study ^9^. Eligible participants were aged between 40 and 69 at recruitment and answered touchscreen, online and verbal questions, as well as agreeing to medical record linkage for general practitioner (GP) READ v2/v3 codes and hospital inpatient ICD-10 codes. A subset of participants completed one or both of two follow-up online mental health questionnaires (MHQ) ^10,11^, the most recent of which included the CIDI-SF panic disorder module.

GLAD+ panic cases were identified through baseline questionnaire responses. Participants who completed the CIDI-SF panic module were panic disorder cases if they met all case criteria, or panic broad cases if they met Criterion A (≥4 panic attack symptoms). Participants who were missing the CIDI-SF but self-reported having ever been diagnosed with panic disorder by a professional were also defined as panic disorder and panic broad cases. The panic attack cases were all panic broad cases who did not meet case criteria for panic disorder.

UKB participants were defined as panic disorder cases if they met all CIDI-SF panic case criteria, and/or had a GP READ code for panic disorder (Supplementary Table 27), although the latter only accounted for ~11% of the cases. UKB participants were defined as panic broad cases if they met at least one of: Criterion A of the CIDI-SF panic module, a GP READ code for panic disorder or attack, the ICD-10 “panic disorder/state/attack” from linked records at baseline assessment. Participants who were missing the CIDI-SF but self-reported having ever been diagnosed with panic disorder by a professional were also defined as panic disorder and panic broad cases. As for GLAD+, panic attack cases were panic broad cases who did not meet panic disorder case criteria.

A common control group across panic phenotypes was created from GLAD+ and UKB participants. From GLAD+, controls were all participants who were not recruited for a history of anxiety or depression (i.e. NIHR BioResource excluding GLAD), had completed and did not meet case criteria for all available anxiety and depression CIDI-SF modules (panic attacks/disorder, depression, generalised anxiety disorder, social phobia, specific phobia, agoraphobia), and did not self-report having ever been diagnosed with any anxiety or depression disorder (panic disorder, MDD, GAD, social anxiety, specific phobia, agoraphobia). UKB controls were participants who did not meet case criteria on the CIDI-SF panic (MHQ2), MDD (MHQ1 and 2) or GAD modules (MHQ1), and did not self-report receiving any anxiety or depression diagnosis at MHQ1 (social phobia, any other phobia, depression, anxiety, nerves or generalized anxiety disorder, agoraphobia) or MHQ2 (depression, generalized anxiety disorder, social phobia, agoraphobia, any other phobia, panic disorder), and had no GP READ or ICD-10 code for any anxiety or depression diagnosis (Table S22).

Ethical approval for the GLAD Study and NBR COPING study was obtained from the London-Fulham Research Ethics Committee (REC reference 18/LO/1218 and 20/SW/0078, respectively, COPING project no. 282754.). The dataset used in the present analysis was freeze 2023-06-07. UKB analyses were conducted under application ID 82087. All participants provided informed consent.

All data from GLAD+ study were genotyped by ThermoFisher on the UK Biobank Axiom Array v1 and v2 across numerous genotyping batches. Genetic data were restricted to individuals from European ancestries (749,044 SNPs before quality control). Ancestry was determined using GenoPred ^12^, by projecting GLAD+ individuals on genomic principal components from the 1000 Genomes reference data, and assigning individuals a genetic ancestry if they lay < 3SD from the mean of individuals from that ancestry superpopulation. Quality control exclusions were conducted for variants with MAF < 0.01, call rate < 0.95, or deviant from Hardy-Weinberg equilibrium (p < 1×10⁻⁸). Individuals were excluded if they: had withdrawn from the study following genotyping, were a duplicate of a higher-quality sample (not including known identical twins), were known to be mislabelled, their genotypic sex (males Fx > 0.8, females Fx < 0.5) did not match their sex assigned at birth, were outliers on genome-wide heterozygosity (|F-hat| > 0.2), or had an excess of relatives (average pi-hat > 3SD from the mean). Following quality control, 33,635 individuals and 484,182 variants were available for imputation. Imputation was carried out to TopMED Freeze 8^13^. Data was then further restricted to MAF ≥0.01 and R2 ≥ 0.3, leaving 15,009,228 variants for analysis.

UKB participants provided blood samples, genotyped on the UK Biobank Axiom Array (90% of the sample) or the UK BiLEVE array^14^. Several sample exclusions were implemented based on quality control procedures performed by the UKB and described elsewhere (i.e., missing rate, heterozygosity, sex discordance)^9^. In addition, we excluded participants with genotype missingness > 0.02 and related individuals identified as KING > 0.04, excluding one participant from each related pair, while preferentially selecting from each pair those participants that answered the MHQ. Imputed dosage data for 487,422 participants based on 670,739 markers was provided by the UKB. Genetic variants were imputed using IMPUTE4 software with the Haplotype Reference Consortium reference panel^15^, 1000 Genomes phase 3^16^, and UK10K Consortium reference panel^17^. SNP quality control involved the exclusion of imputed SNPs with MAF < 0.01 and INFO < 0.4. After quality control procedures we obtained a sample size of 413,259 participants of mixed ancestry and 9,830,370 imputed genotypes. Ancestry classification into super populations was implemented using 4 means-clustering on the first two ancestry-informed principal components, identifying 397,319 European ancestry participants.

To maximise power in this case-control analysis, we pooled the participants into a single sample, with GLAD+ providing most cases, and UKB the controls. We merged genotype data from GLAD+ and UKB using Plink 1.9. For the GWAS analysis, we performed a final round of quality control on the combined sample, restricting to variants with MAF > 0.01, missing rate < 0.02, Hardy-Weinberg equilibrium exact test p-value > 1×10⁻⁸, and individuals with missingness < 0.02.

We used KING^9^ to assess relatedness between GLAD+ and UKB participants. A total of 1,815 participant pairs shared identical genetic data; we assumed that pairs within the same dataset represent twins, while pairs across the two datasets represent duplicates (n = 1,633). Where a participant had provided data for both studies (‘duplicates’), we preferentially retained the higher-quality UKB blood-based genotype data, complete phenotype data, and evidence of meeting case criteria, and removed the other data for that participant. Principal components were projected onto the whole pooled sample from unrelated individuals using flashpca^18^. The resulting sample was 488,349 individuals of which 186,224 had panic data, and 8,274,311 imputed SNPs. The GWAS was performed in Regenie version 3.1.3^19^ with covariates of batch, array and the first 10 genetic principal components. For x-chromosome analysis, sex was included as a covariate, and Regenie automatically recodes male genotypes as diploid (0/2) to match the dosage scale of female genotypes.

#### QIMR Genetics of Bipolar Study

The Australian Genetics of Bipolar Disorder (GBP) cohort consists of 6,682 participants (mean age: 44.8 ± 13.6 years) who completed a self-reported questionnaire, with 4,706 individuals (70%) genotyped from saliva samples. Comorbid psychiatric disorders, primarily anxiety-related, were reported by 64.5% of participants.^20^ DNA samples were collected via Isohelix GeneFix GFX-02 2 mL Saliva Collection Devices and genotyping was conducted using the Illumina Global Screening Array V.2.0. Panic broad cases met criterion A of the CIDI-SF. Panic disorder cases met the full CIDI-SF for panic disorder. Controls were the same for broad, disorder and attacks; they did not report a comorbid depression or anxiety diagnosis and did not meet criteriafor any anxiety disorder or depression using the CIDI-SF . For the simple screened sensitivity analysis, controls were individuals that did not meet criteria for panic attacks or disorder (but may have other anxiety or depressive disorders). GWAS were conducted using SAIGE (v1.3.0) with logistic mixed models, adjusting for sex and the first four principal components (PCs). Quality control procedures excluded genetic variants with a MAF < 0.01 and INFO < 0.6. For the association of genetic variants on the X chromosome, the genotypes in males were coded as 0/2.

#### TEDS: Twins’ Early Development Study

The Twins’ Early Development Study (TEDS)^21,22^ is a population-based cohort of twins born in England and Wales between 1994 and 1996. All genotyped participants are of European ancestry, and include a maximum of one individual per monozygotic pair, and up to two per dizygotic pair.

Panic cases and controls were identified through questionnaires administered at the age 26 data collection wave. Participants were defined as panic disorder cases if they met all case criteria on the CIDI-SF panic module, or panic attack cases if they met Criterion A (≥4 panic attack symptoms). Participants who were missing the CIDI-SF but self-reported having ever been diagnosed with panic disorder by a professional were also defined as panic disorder and panic attack cases.

A common control group was created who had completed all anxiety and depression CIDI-SF modules (panic disorder, depression, generalised anxiety disorder, social phobia, specific phobia, agoraphobia) and did not meet case criteria for any, and did not self-report having ever been diagnosed with any anxiety or depression disorder (missing or responded no for panic disorder, depression, GAD, social phobia, specific phobia, agoraphobia).

TEDS has been granted ethical approval by the King’s College London ethics committee (References: PNM/09/10-104 and HR/DP-20/21-22060). Informed consent was obtained from all participants. This research was conducted under TEDS data request ID 642. Funding: UK Medical Research Council (MRC) programme grant (MR/V012878/1) to TC Eley (previously MR/M021475/1 awarded to R Plomin).

TEDS participants were genotyped in two waves. At wave 1, buccal cell samples were genotyped on AffymetrixGeneChip 6.0 arrays at the Wellcome Trust Sanger Institute. At wave 2, saliva samples were genotyped on Illumina HumanOmniExpressExome-8v1.2 arrays at the Social, Genetic and Developmental Psychiatry Centre, King’s College London. These data were imputed to HRC 1.1 and harmonised, with quality control steps applied (INFO > 0.75, genotype missingness < 2%, individual missingness <2%, MAF > 1%, HWE > 1x10^-6^)^23^. The GWAS was performed in Regenie version 3.1.3^19^, with the covariates batch and the first 10 genetic principal components.

**Pre-existing panic meta-GWAS summary statistics (*Forstner et al. 2021*)*:***

We used summary statistics from a previous panic meta-GWAS performed using six GWAS of patients and controls from Denmark, Estonia, Sweden, and Germany, consisting of 2,147 cases and 7,760 controls. Cases had a lifetime diagnosis of panic disorder according to DSM-III-R, DSM-IV, or ICD-10 criteria. As all cases met panic disorder criteria, with no additional panic attack phenotype, the same summary statistics were used for both the panic broad and panic disorder meta-analysis. A panic attacks only phenotype had not been created as part of this study and therefore was not included in our panic attacks meta-analysis. Of the three German studies, one screened controls for any mental health problem, one for any anxiety or affective disorder, and one used unscreened controls. The Swedish study screened controls for lifetime diagnosis of schizophrenia, schizoaffective disorder or bipolar disorder, and the Danish and Estonian controls were screened for any mental disorder. These control definitions were modestly different to our primary analysis, which screened for any anxiety or depression disorder. All participants provided written informed consent. Please see the publication for further details of phenotyping, genotyping, and association analysis.

### Supplementary Results

#### Regional Association Plots

Presented for the 39 loci significant across any of the six phenotypes (panic broad, panic disorder, panic attacks ‘super screened’ for any anxiety or depression disorder; panic broad, panic disorder, panic attacks ‘simple screened’ for panic broad).

***rs7540965***
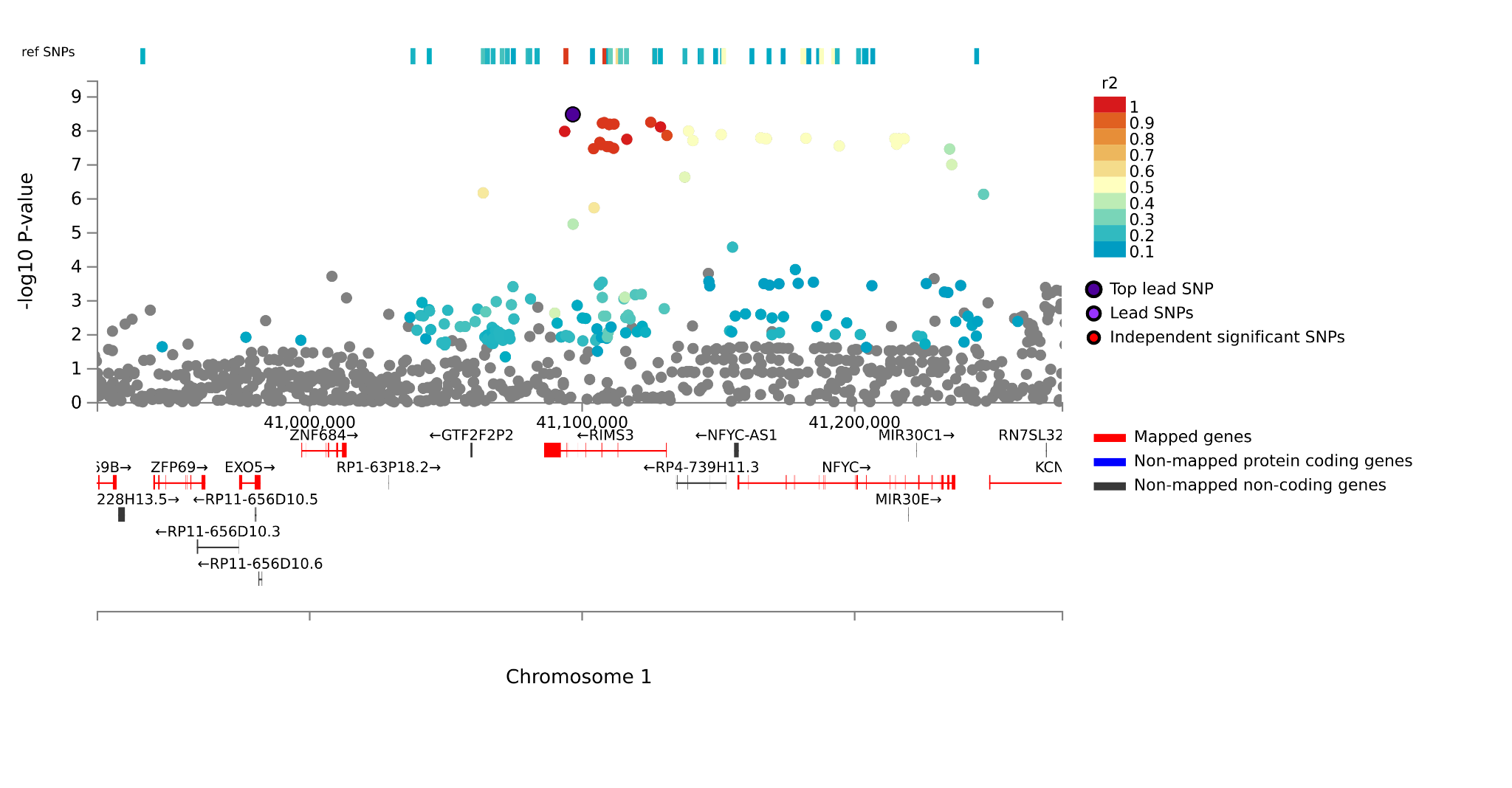

***rs1841499***
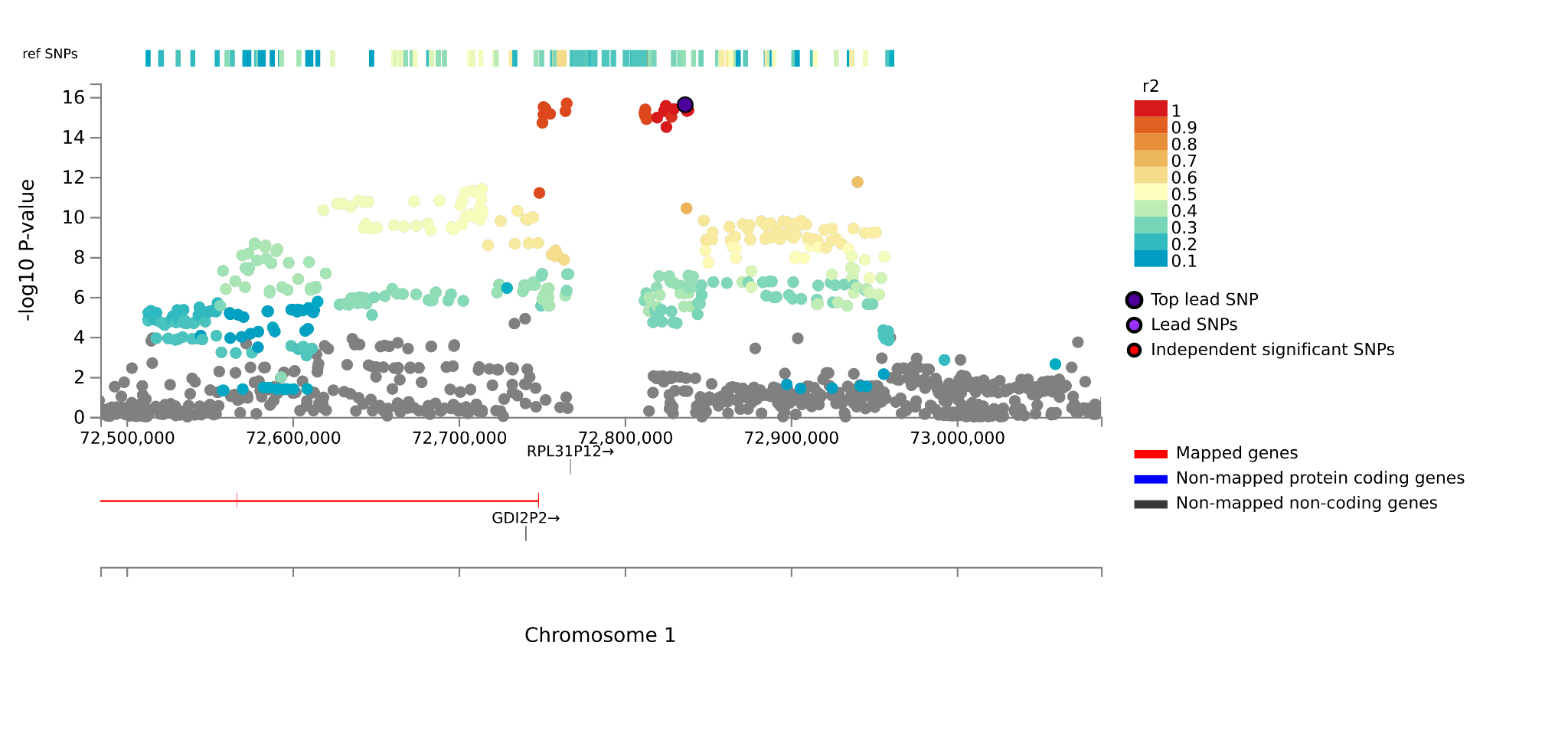

***rs3101336; rs3964487***
These are COJO independent SNPs but in FUMA with 500kb and LD (r2 > 0.1) parameters they are grouped into the same locus, as seen below.

***rs1289881***
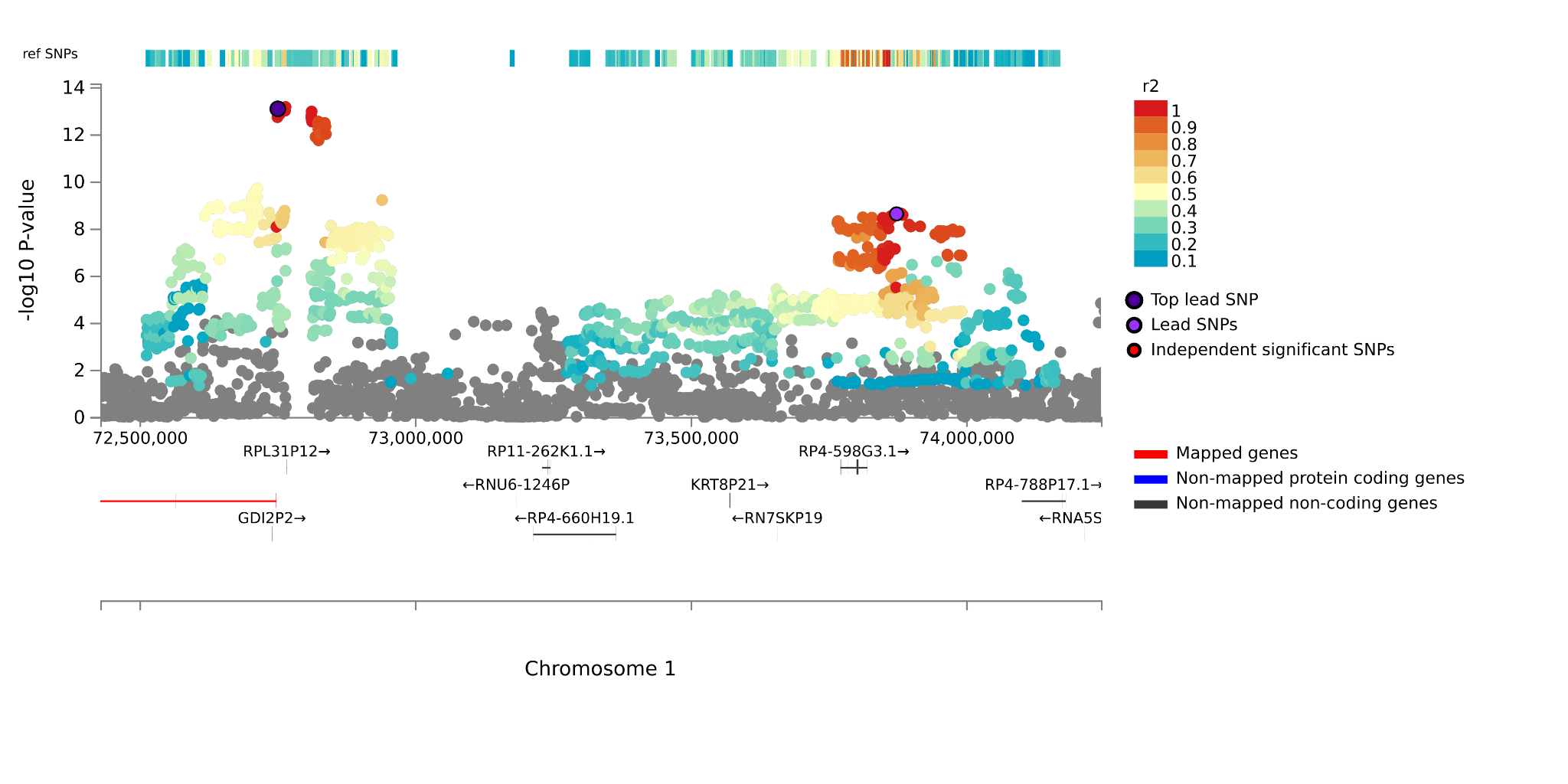

***rs10159228***
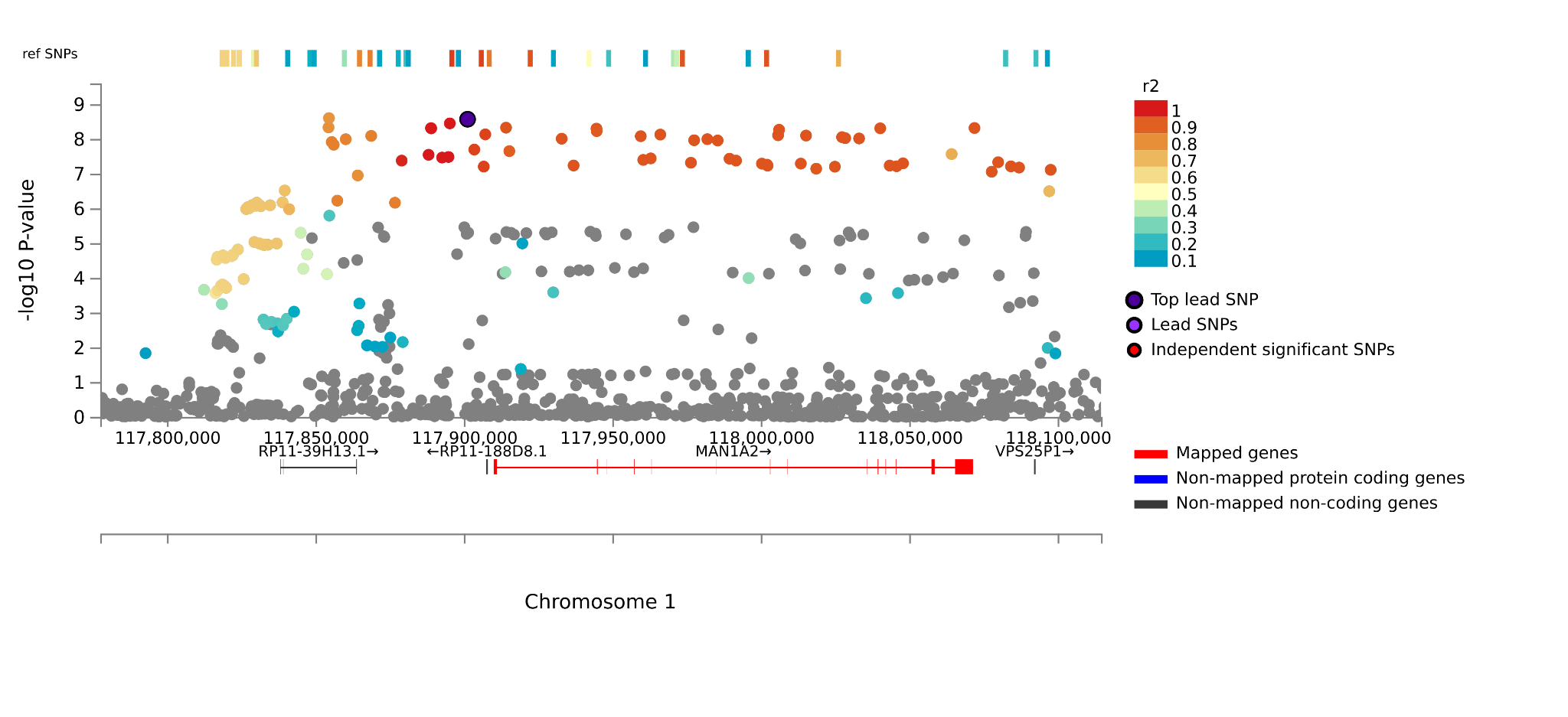

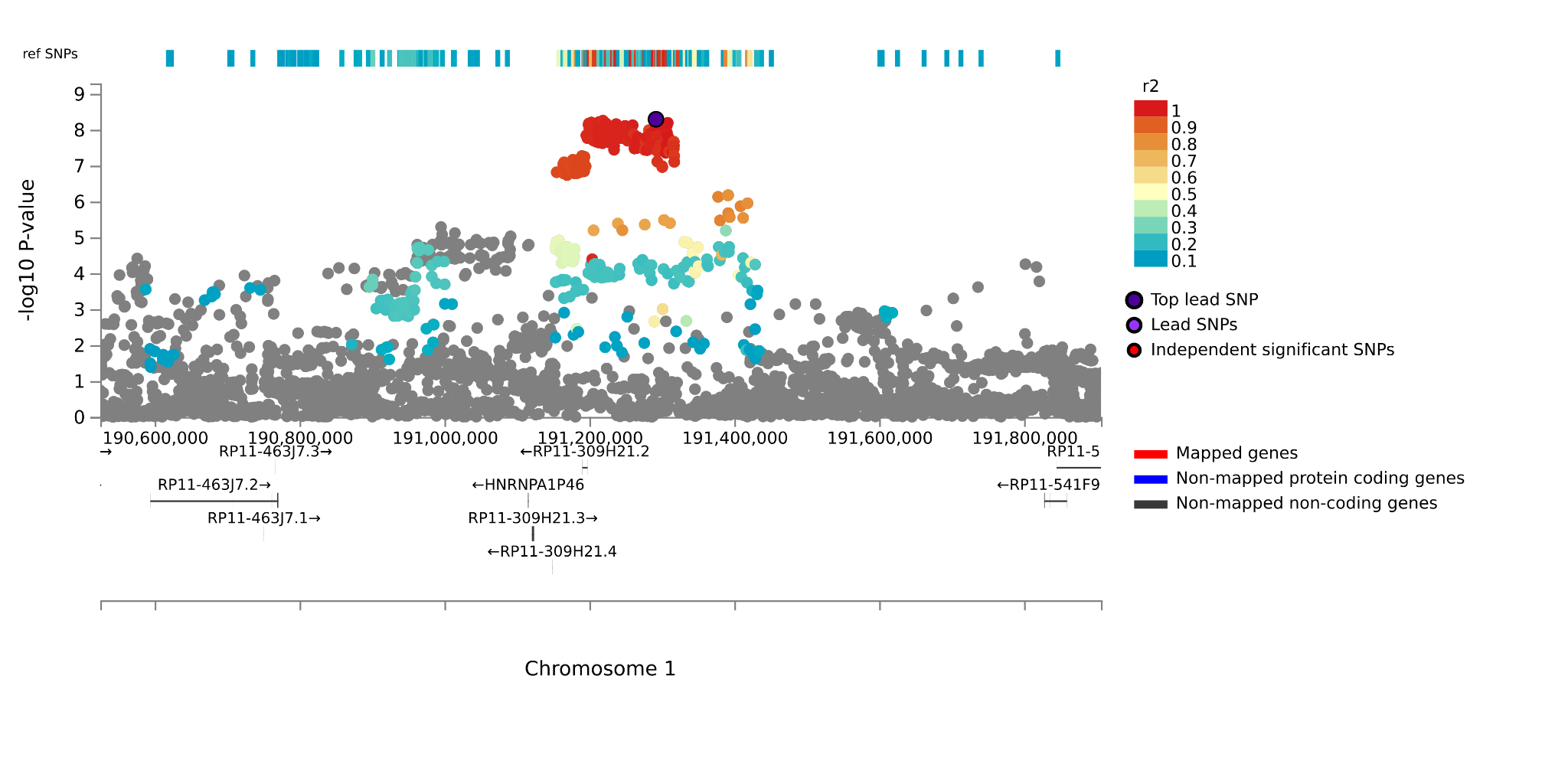

***rs13011472***
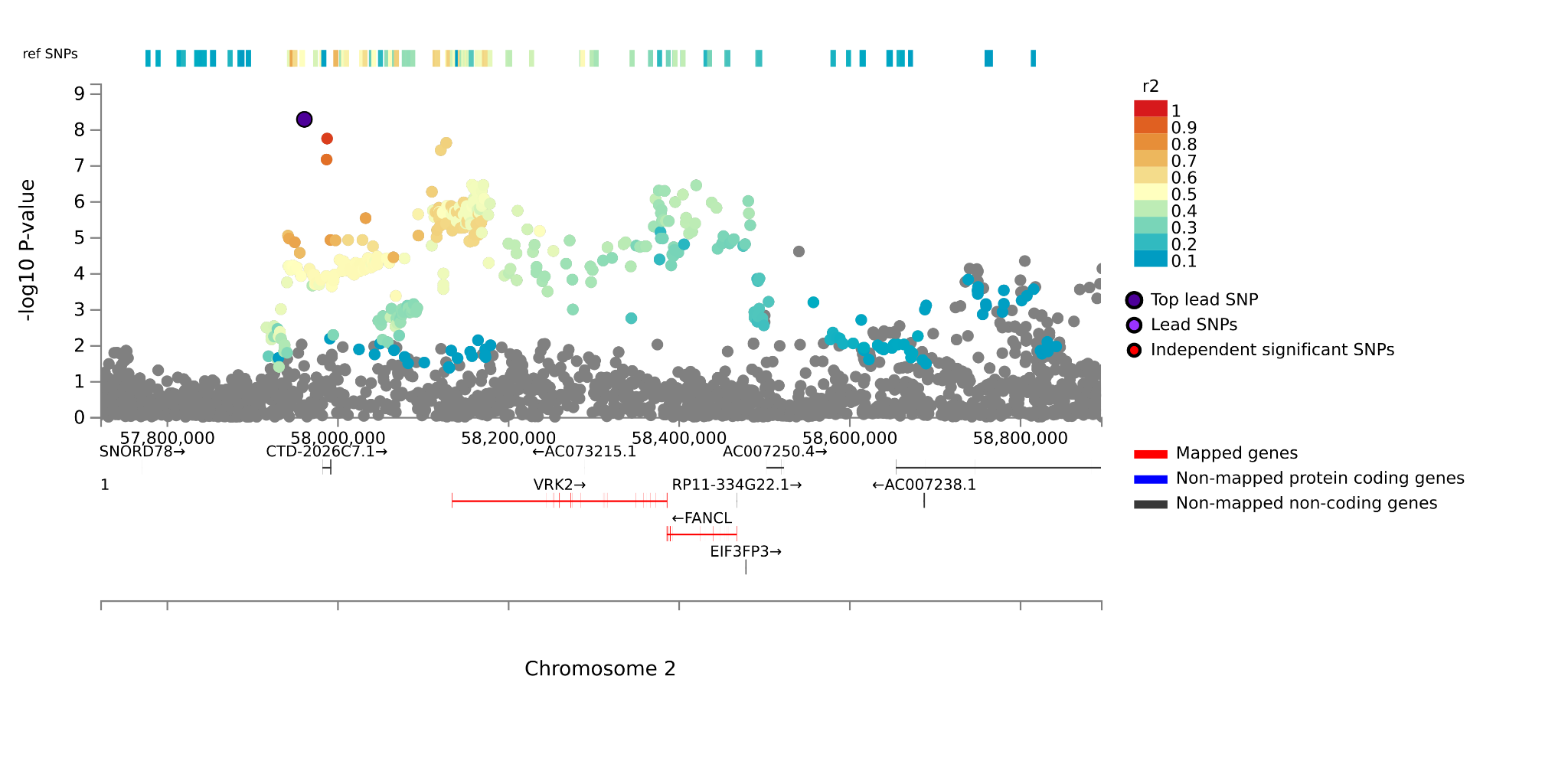

***rs10202231***

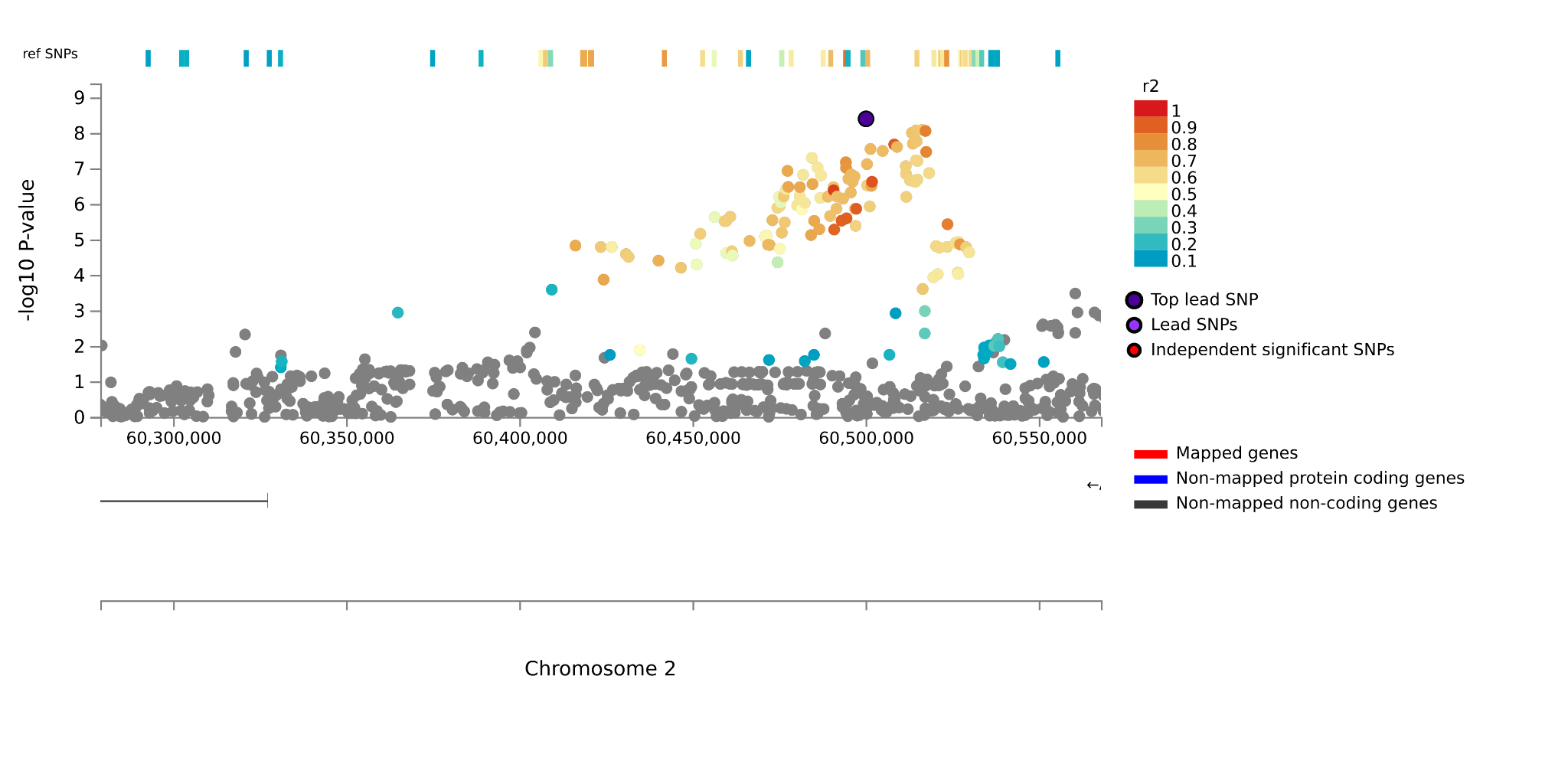

***rs352186***

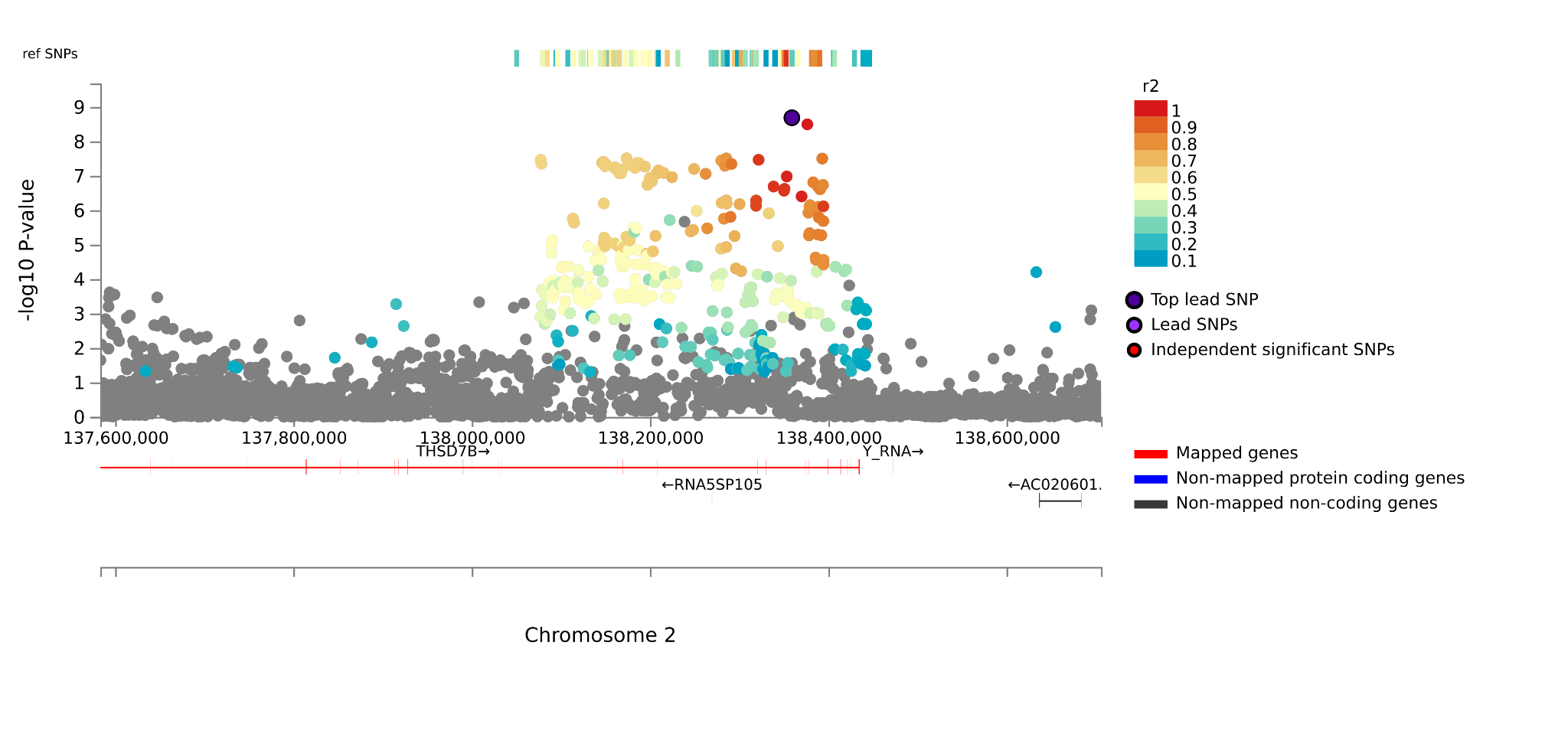

***rs10497101***

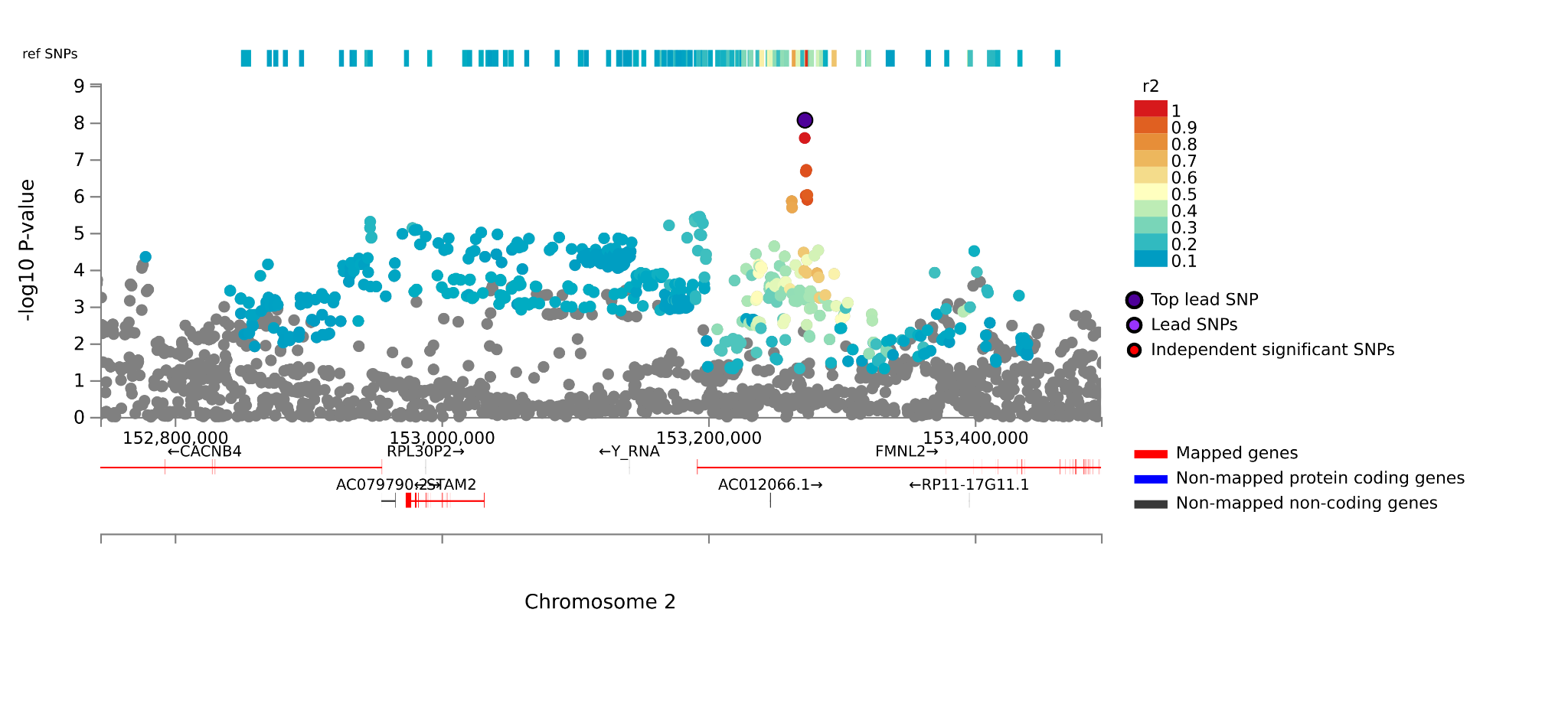

***rs16854051***
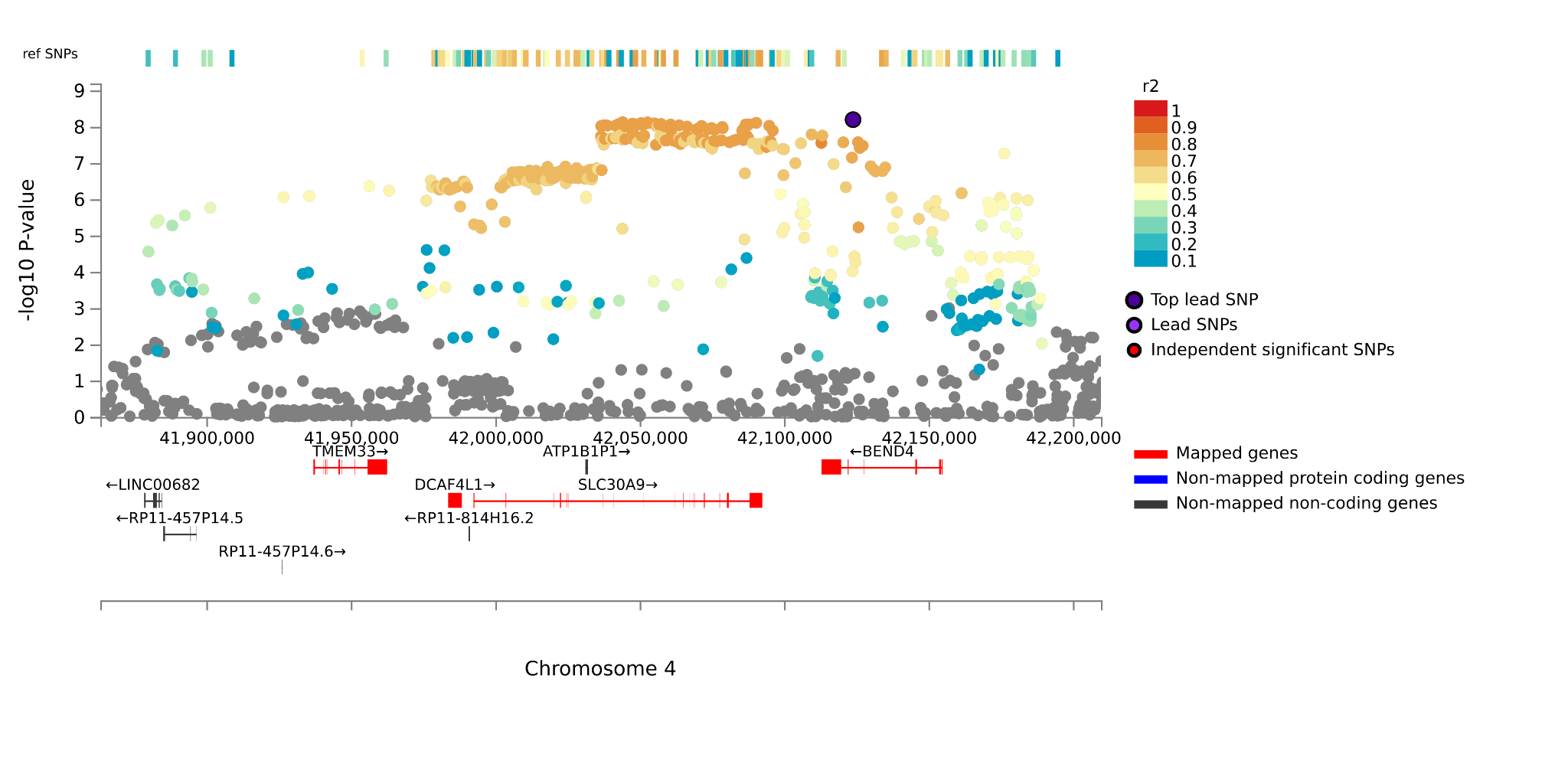

***rs41533650***

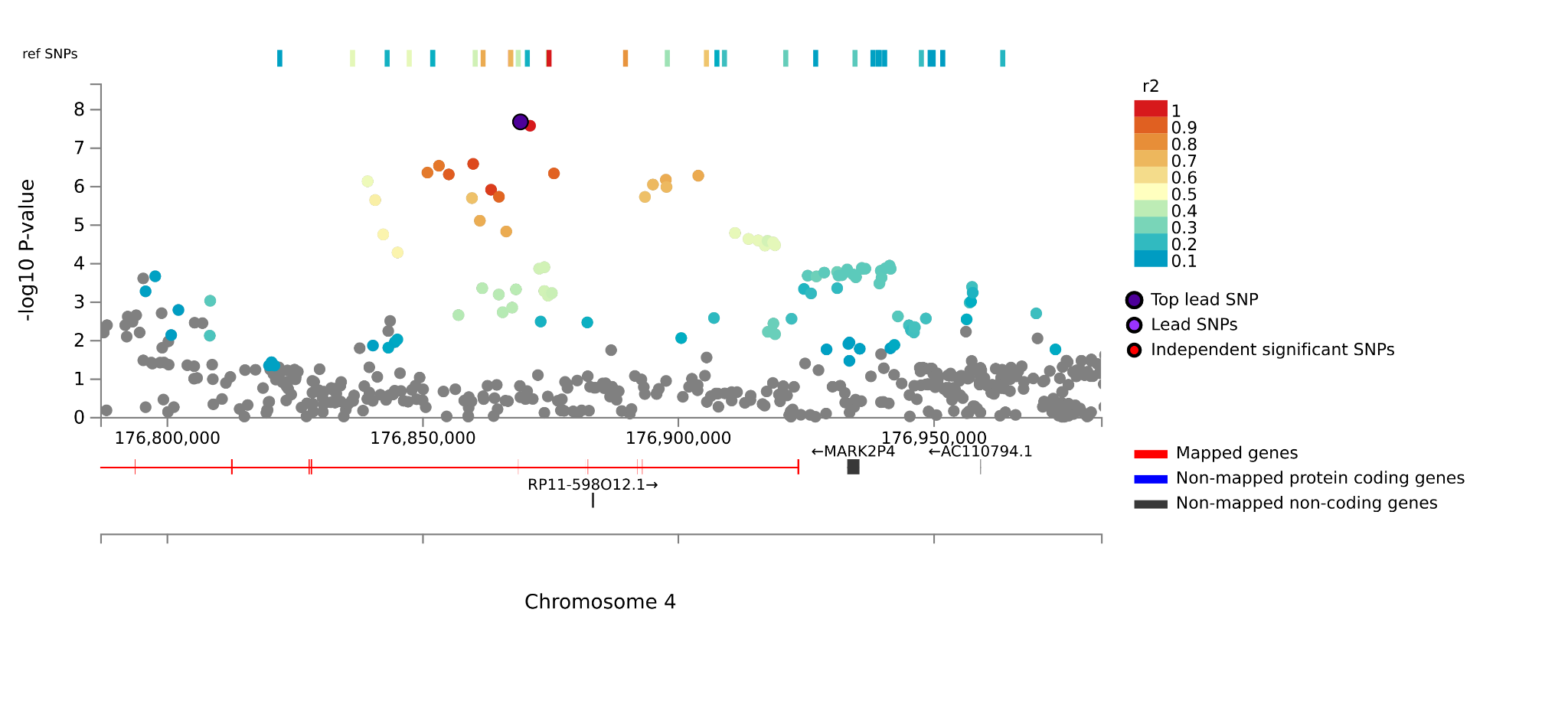

***rs17280797***
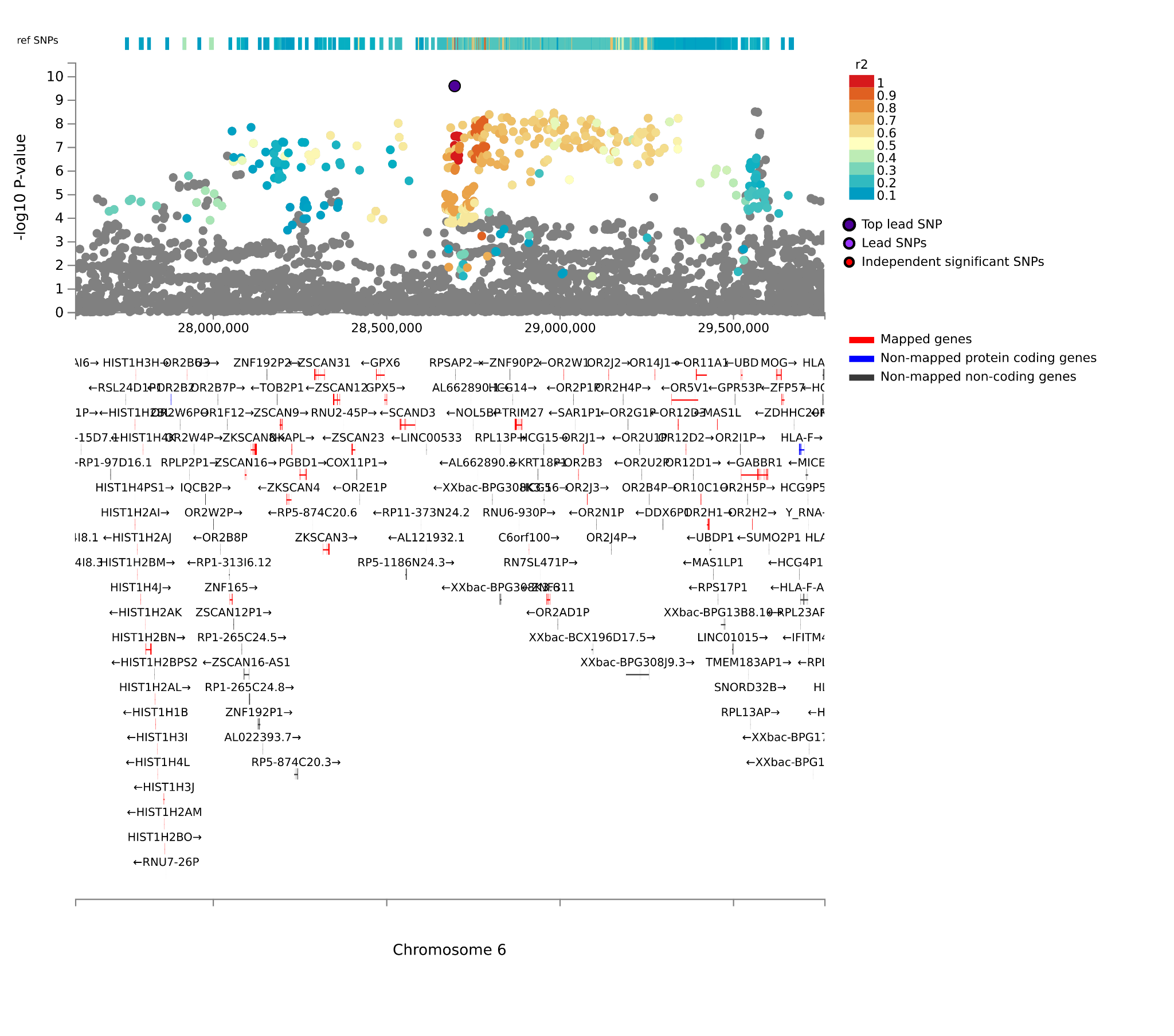

***rs583129***

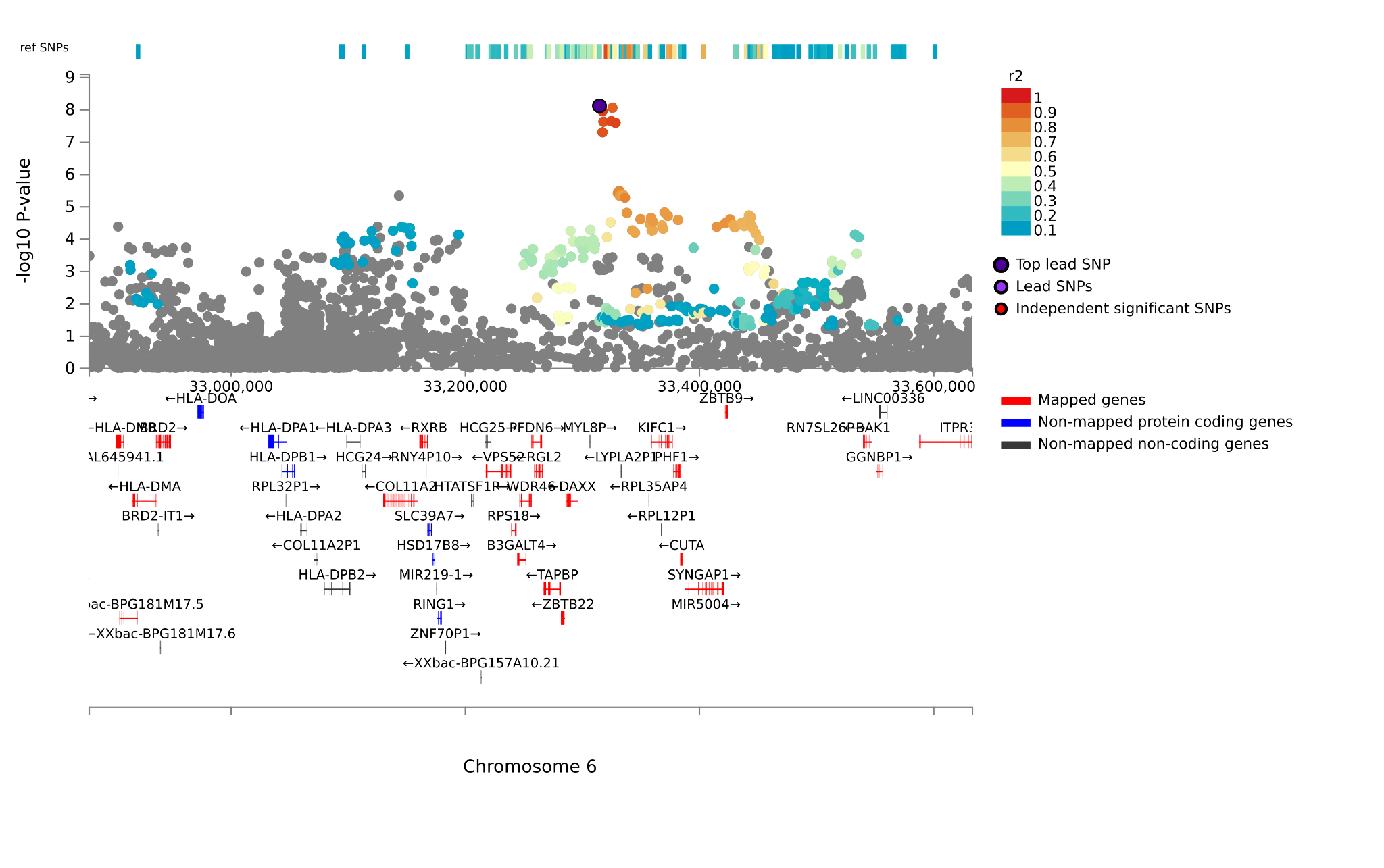

***rs11154014***

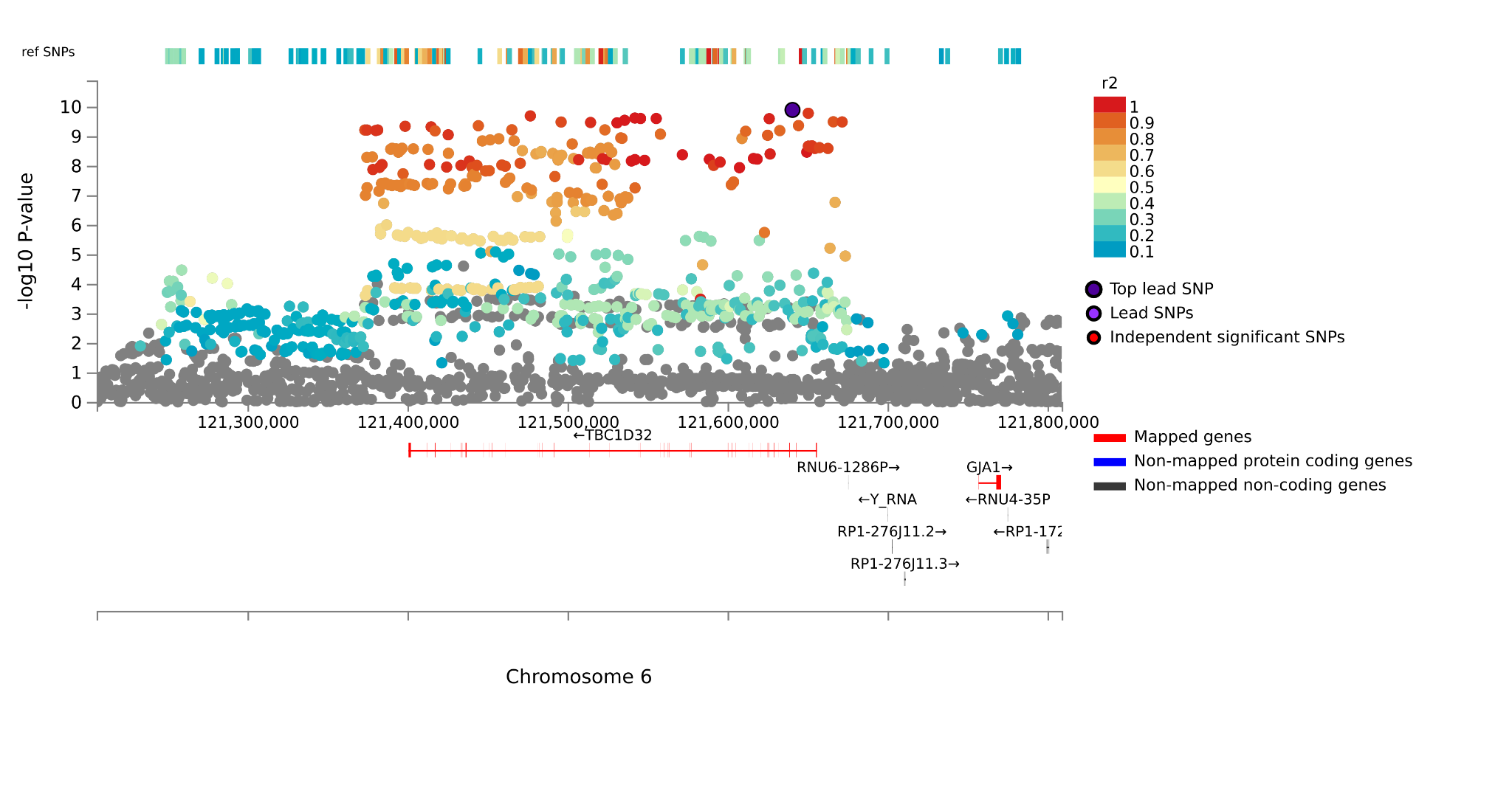

***rs10081338***

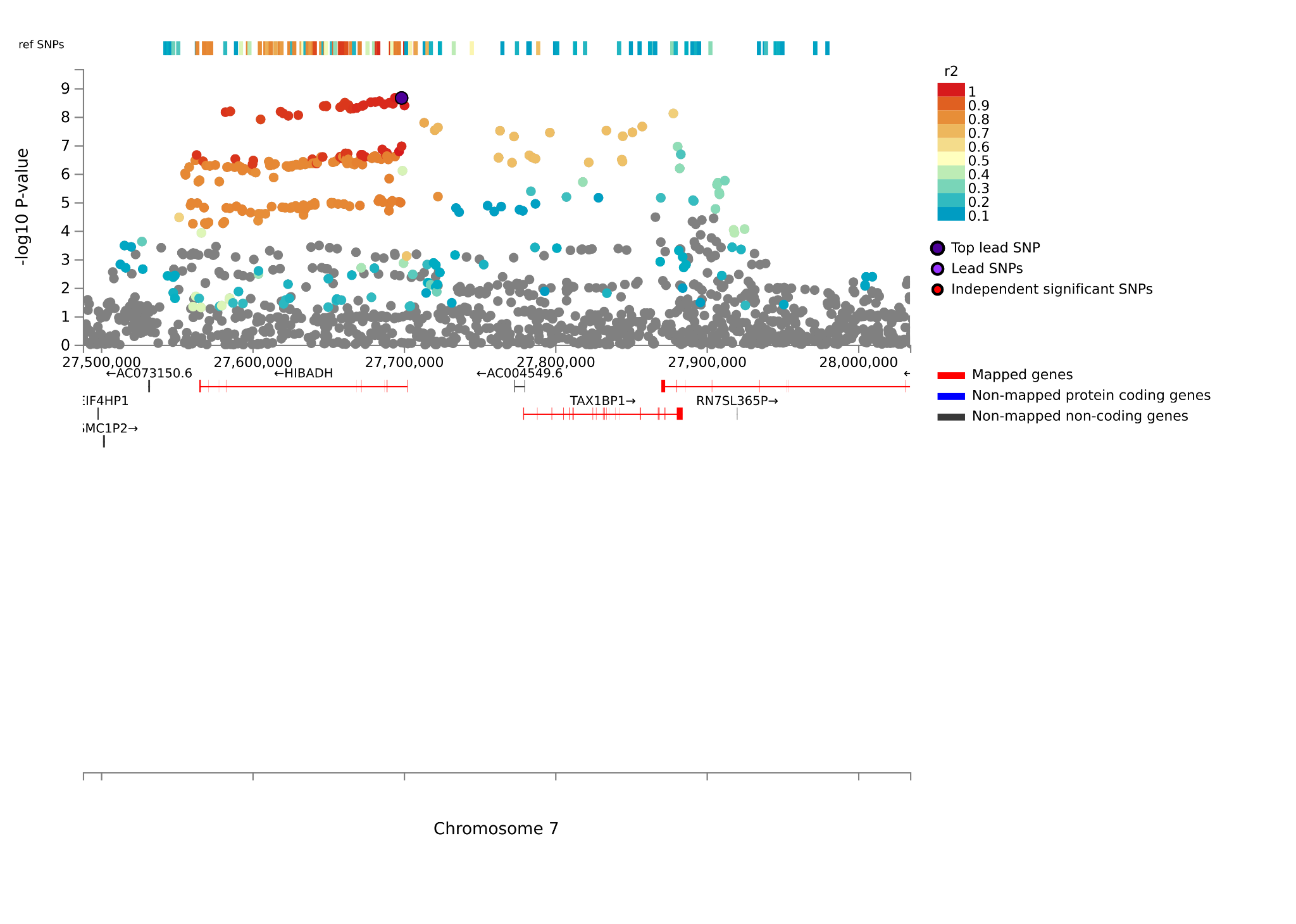

***rs1510302***

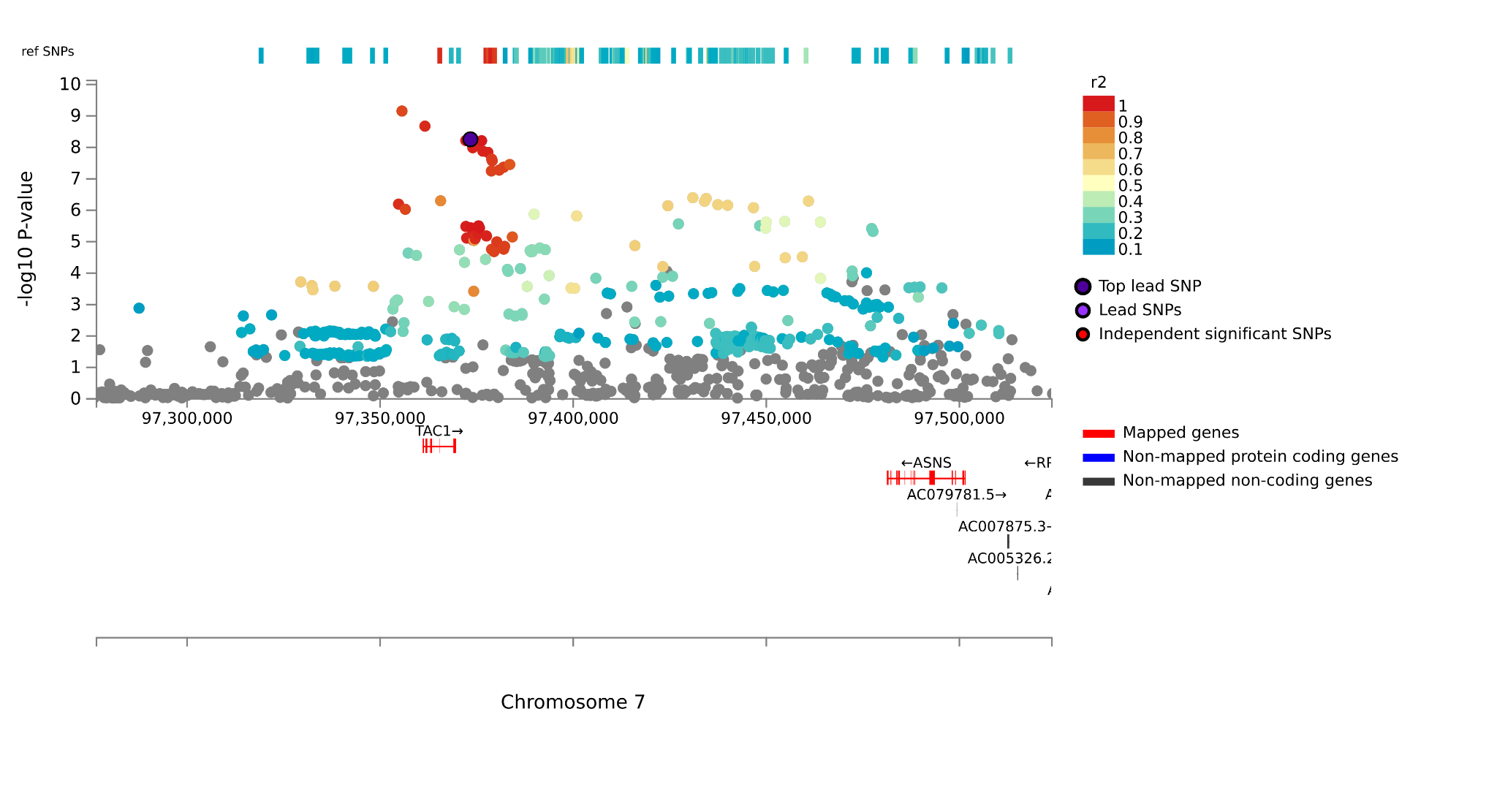

***rs59369558***

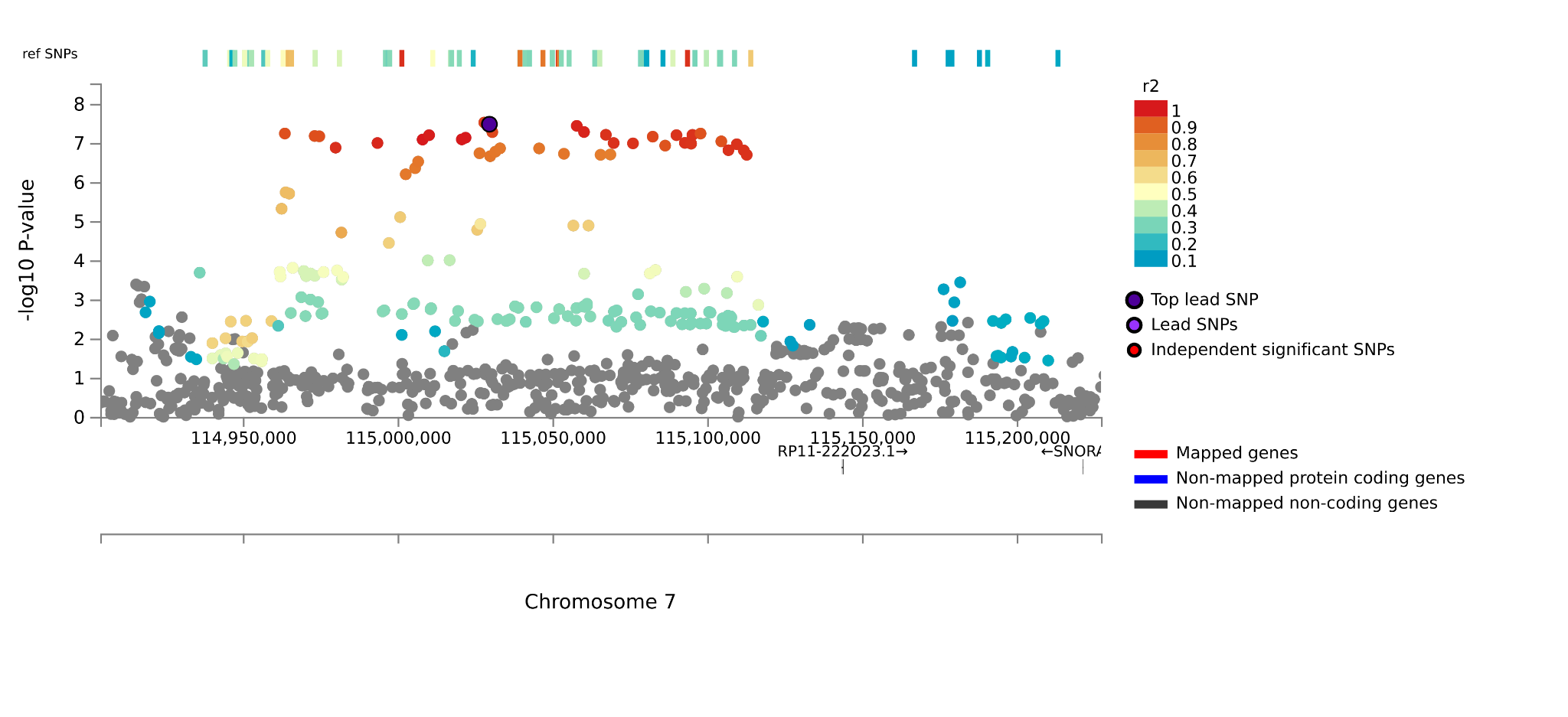

***rs34038233***

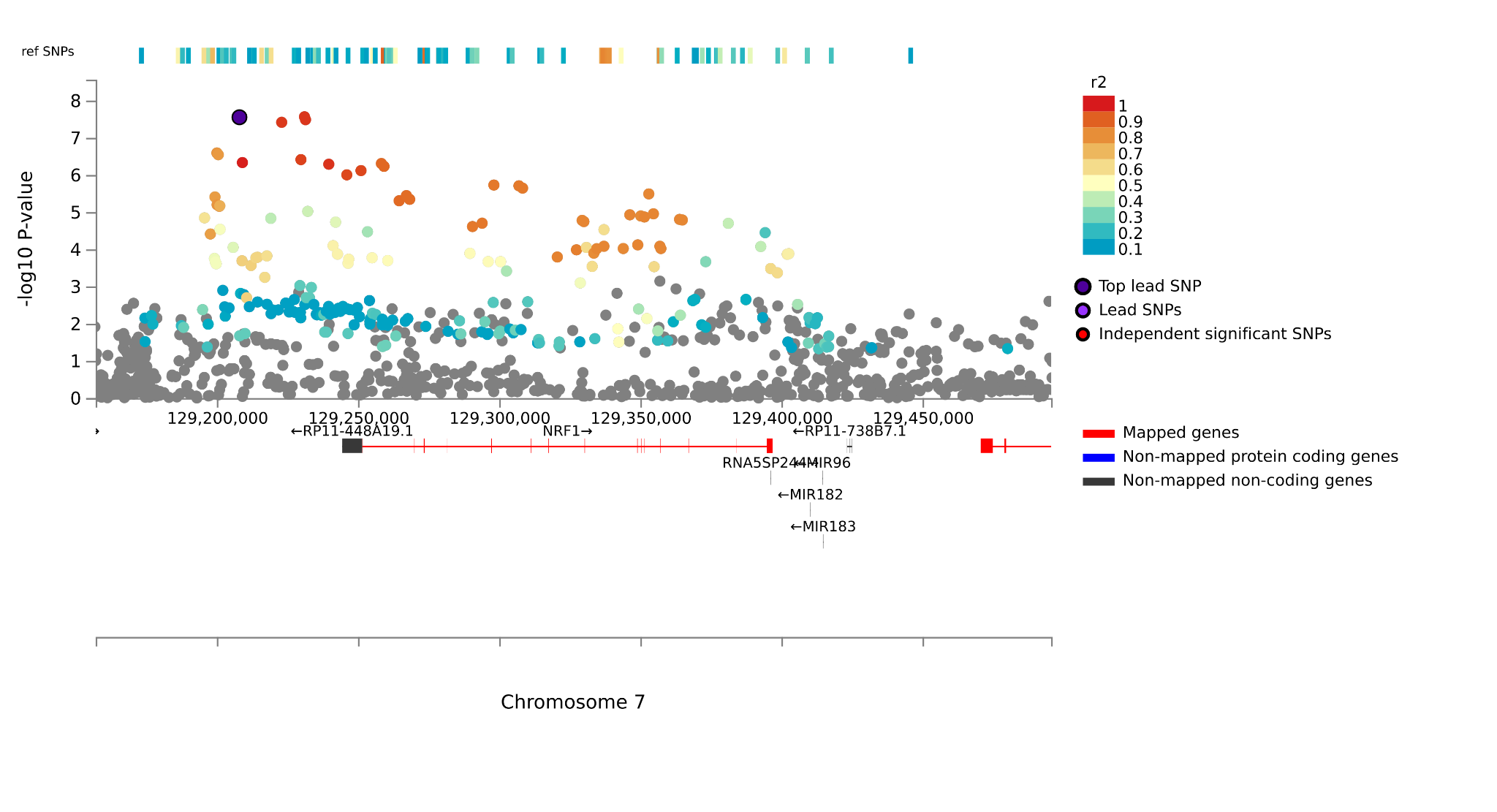

***rs4483247***

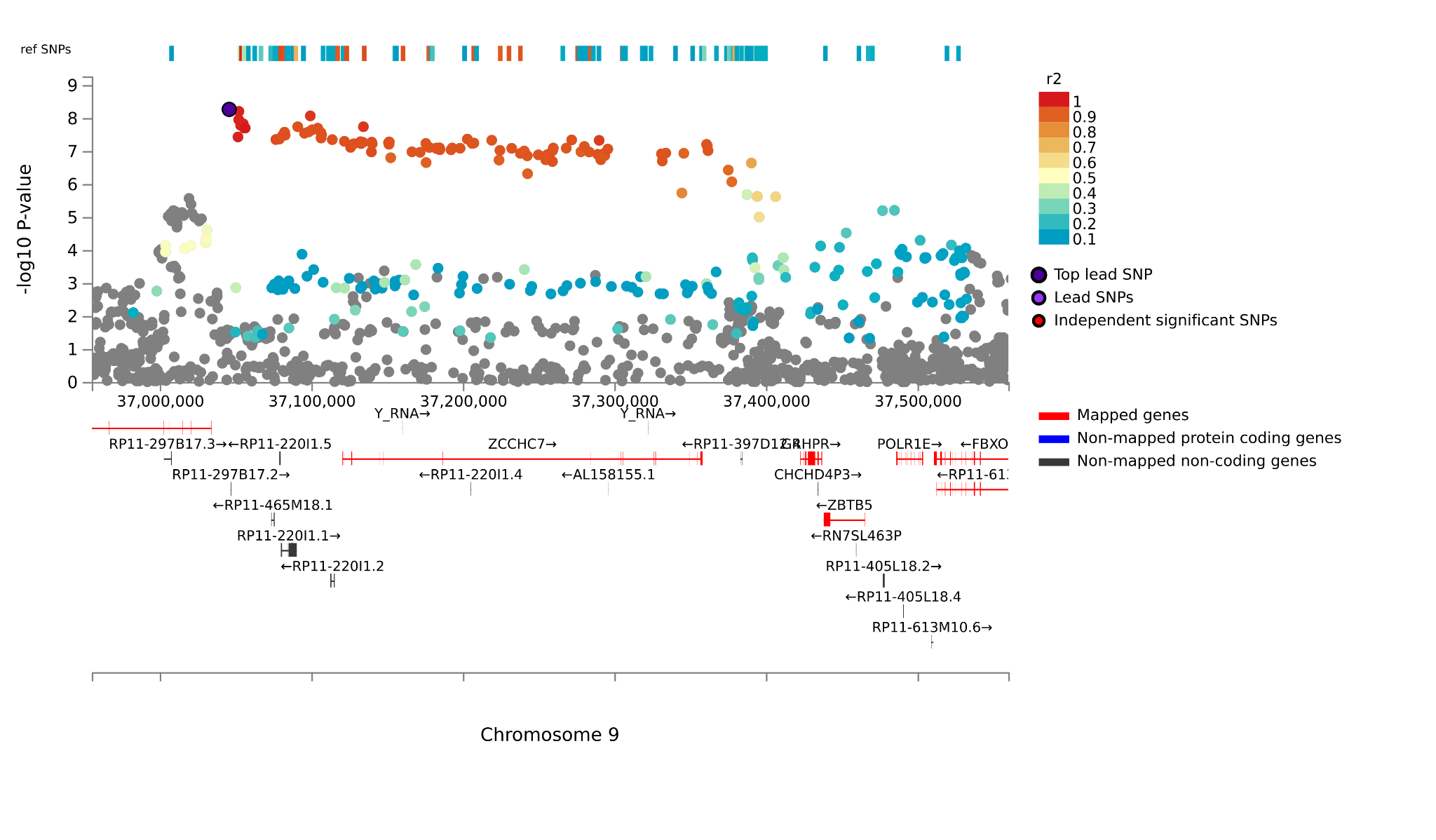

***rs7922812***

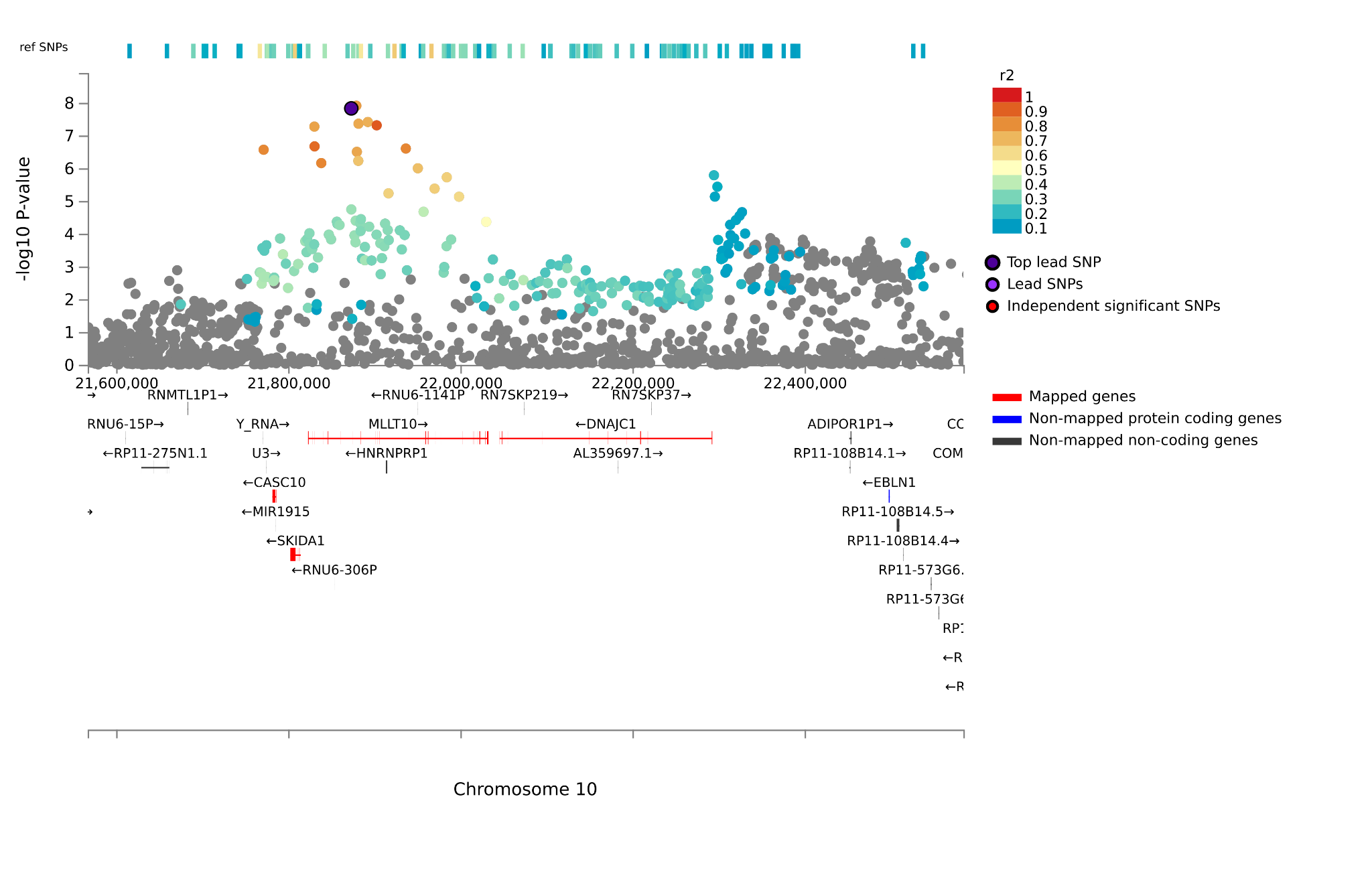

***rs1806153***

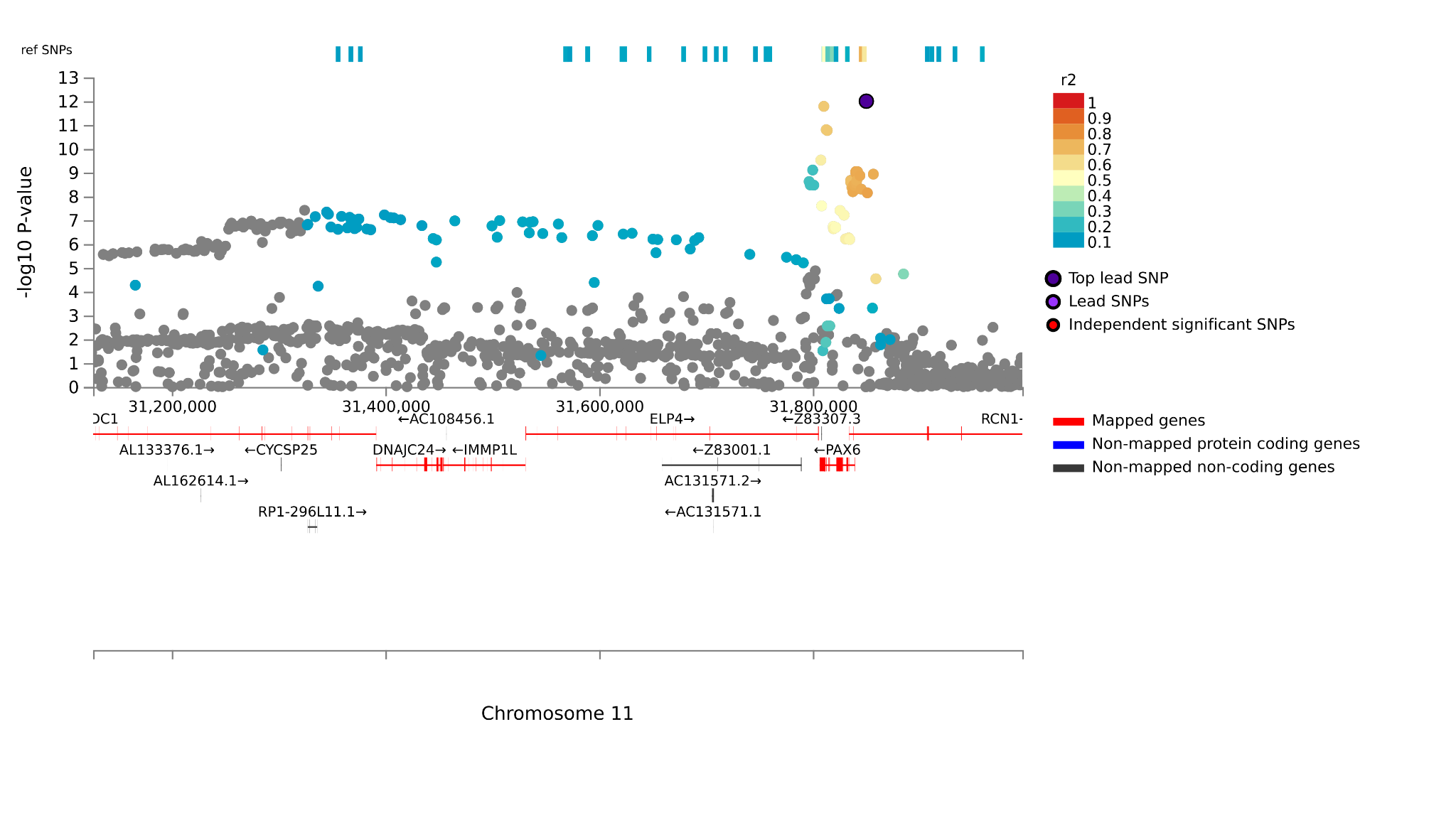

***rs7107356***
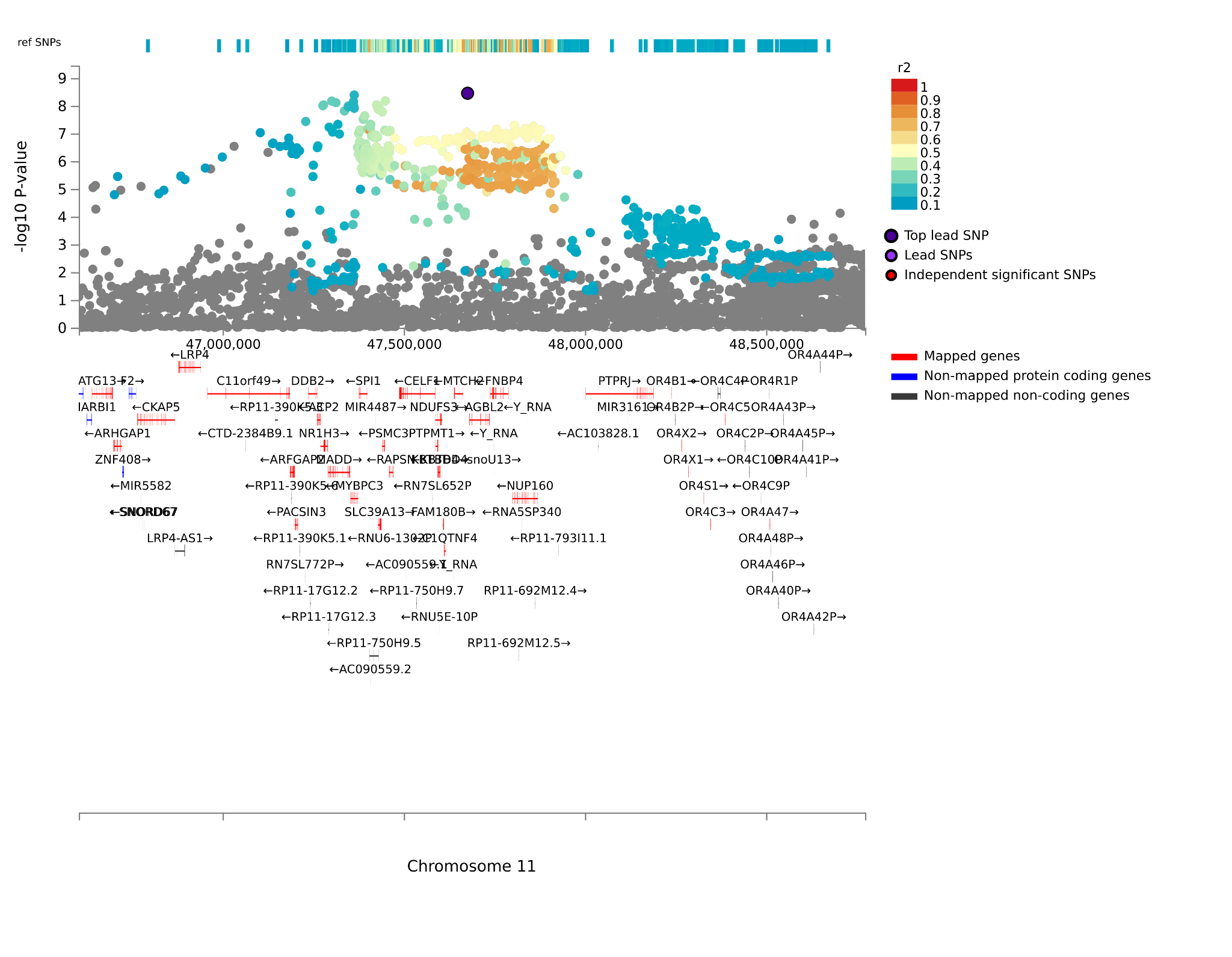

***rs11835982***

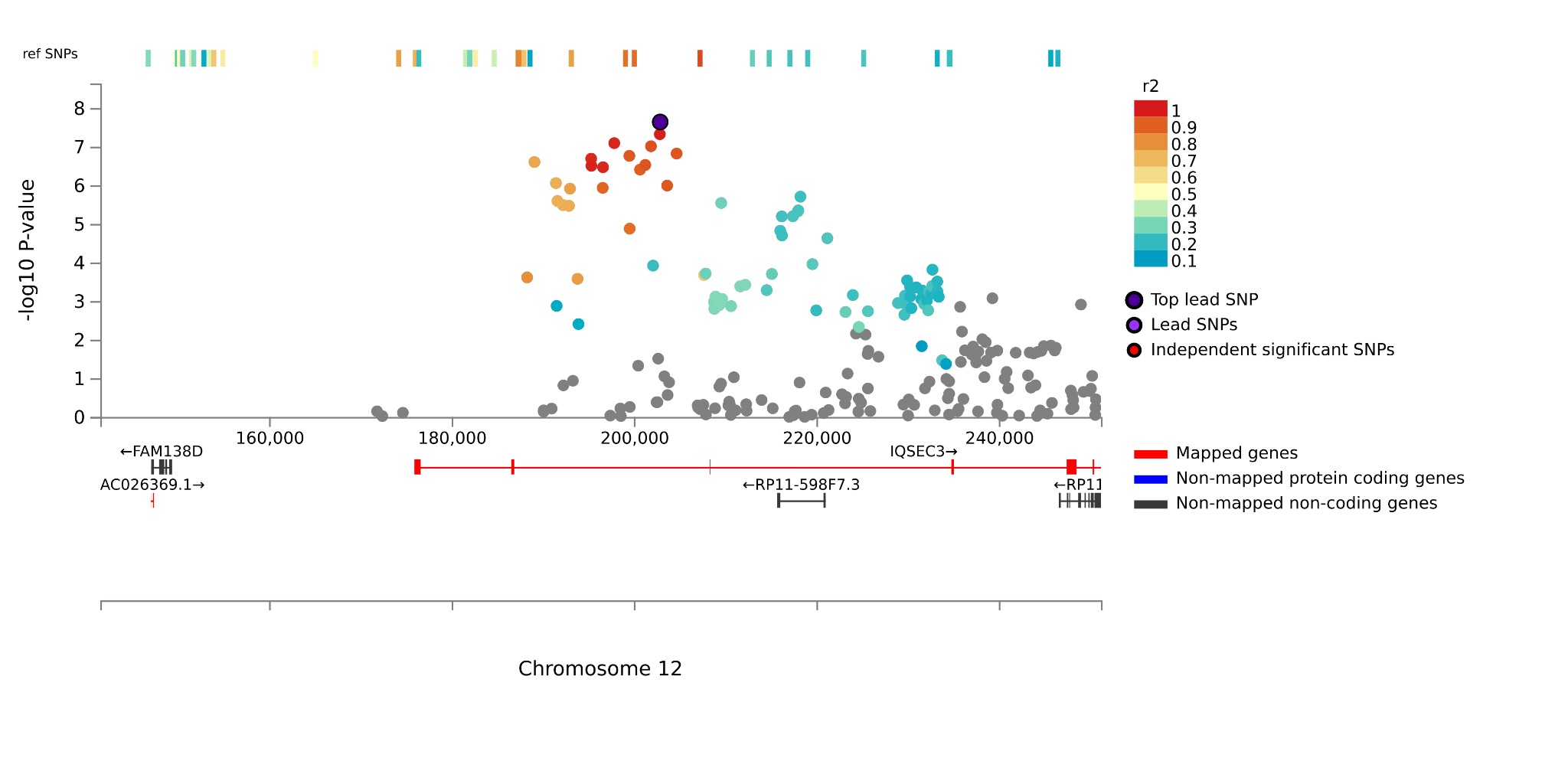

***rs4882465***

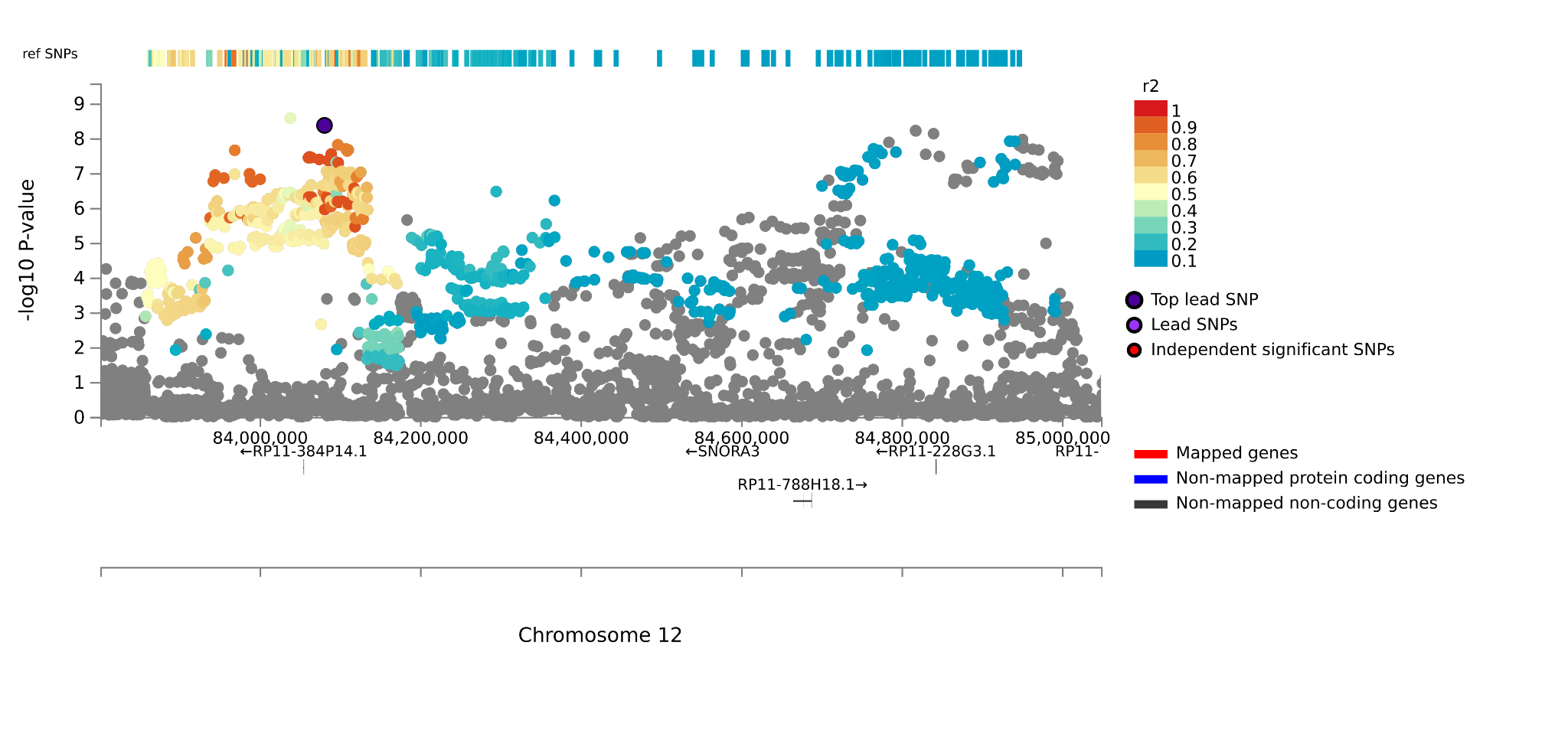

***rs9527069***

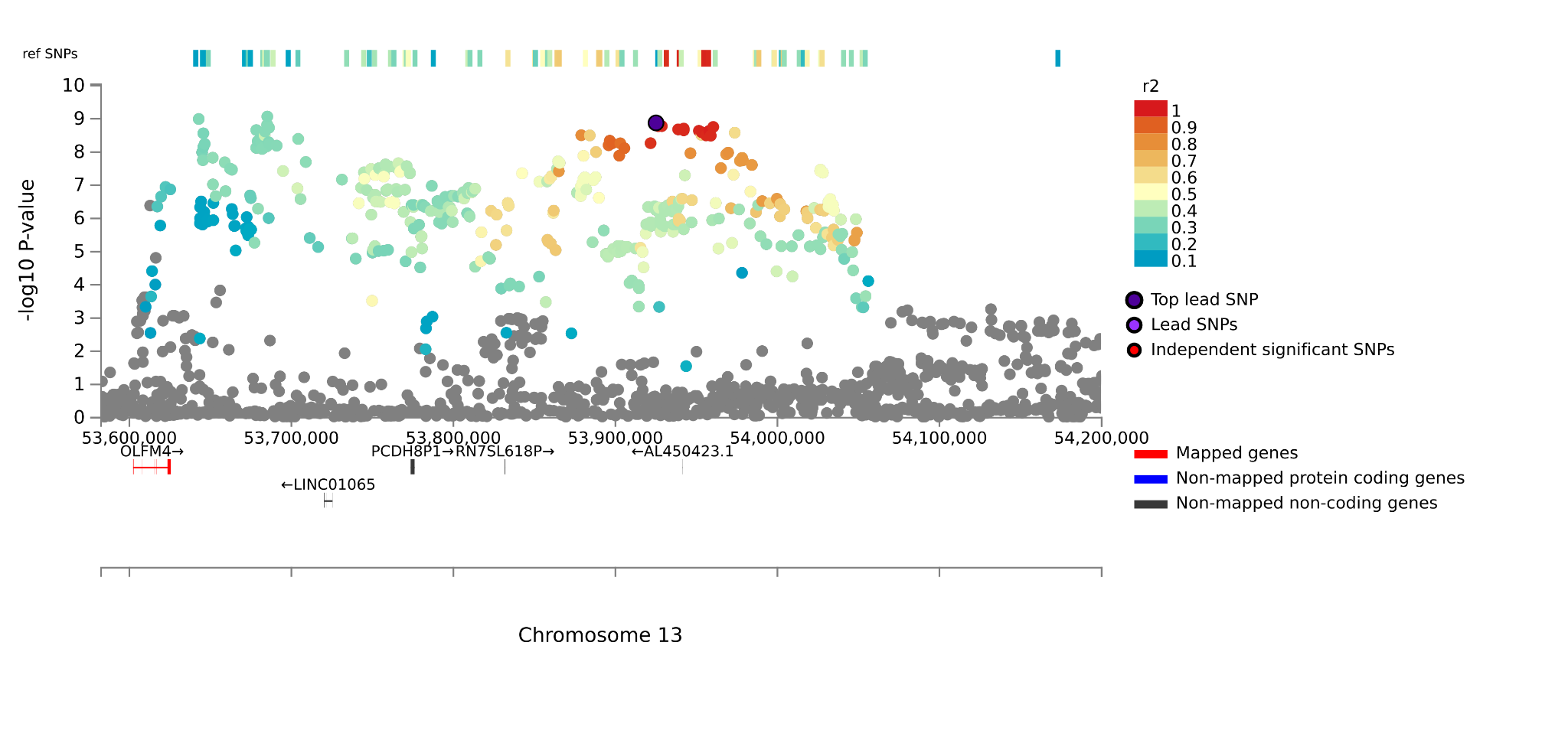

***rs9317586***

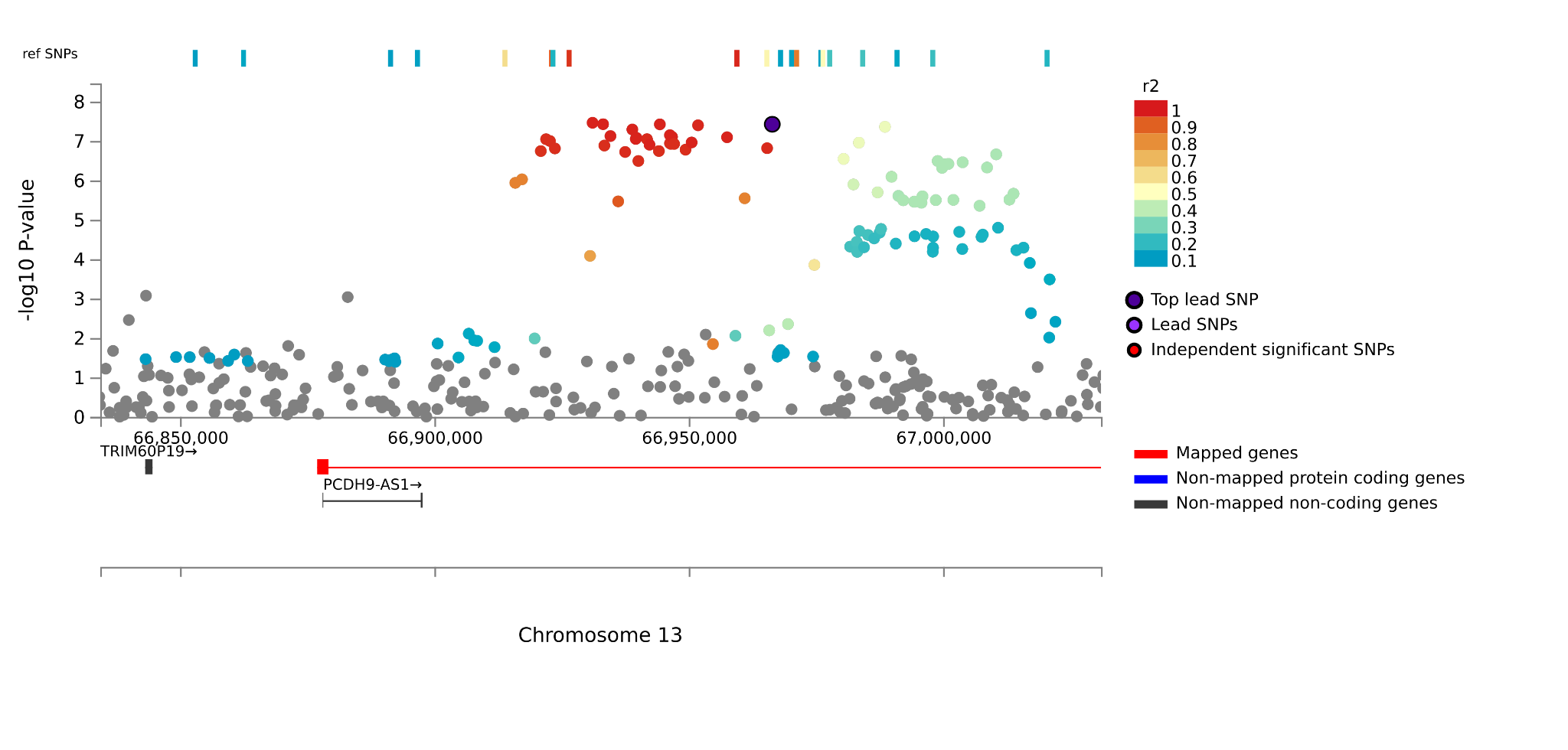

***rs9589493***

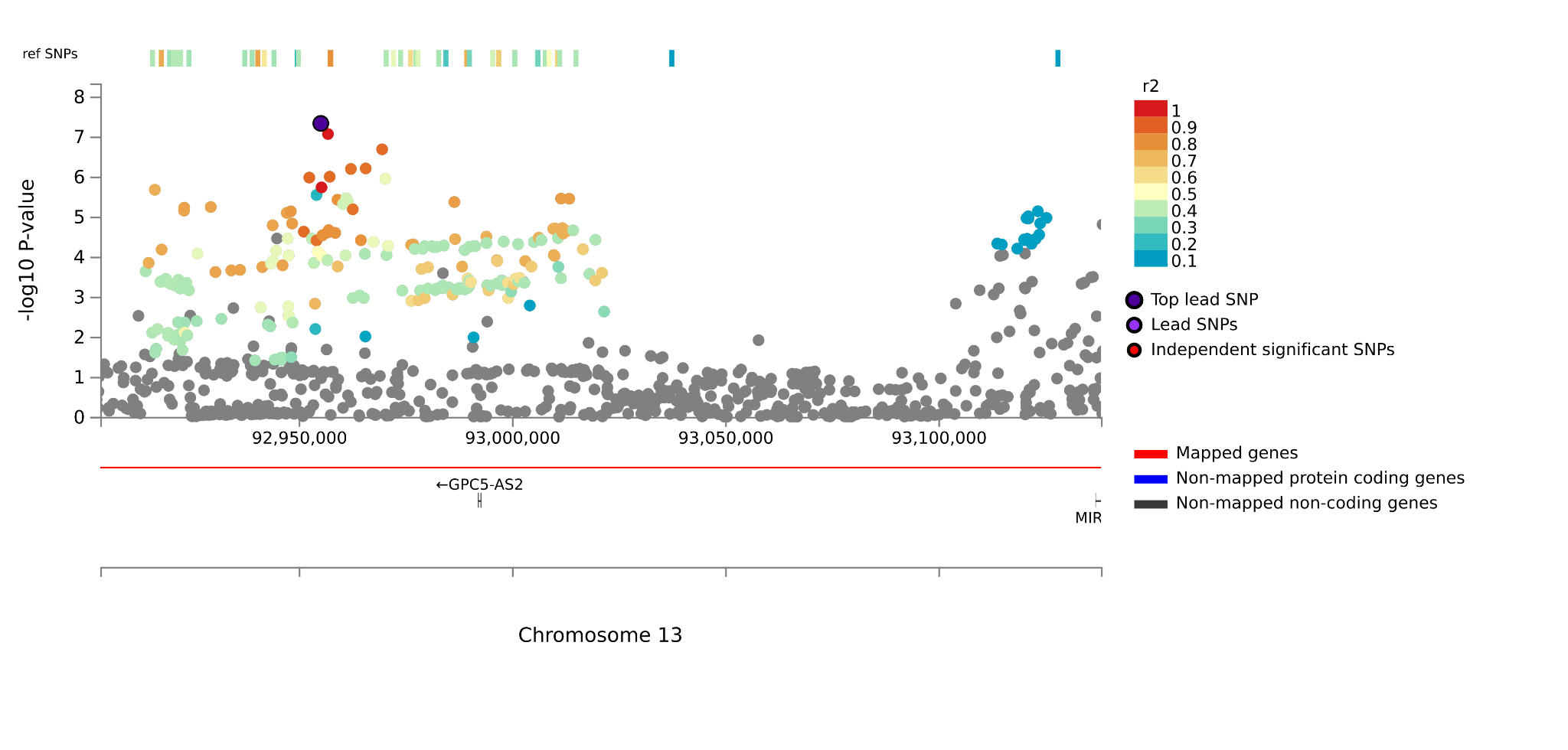

***rs28426374***

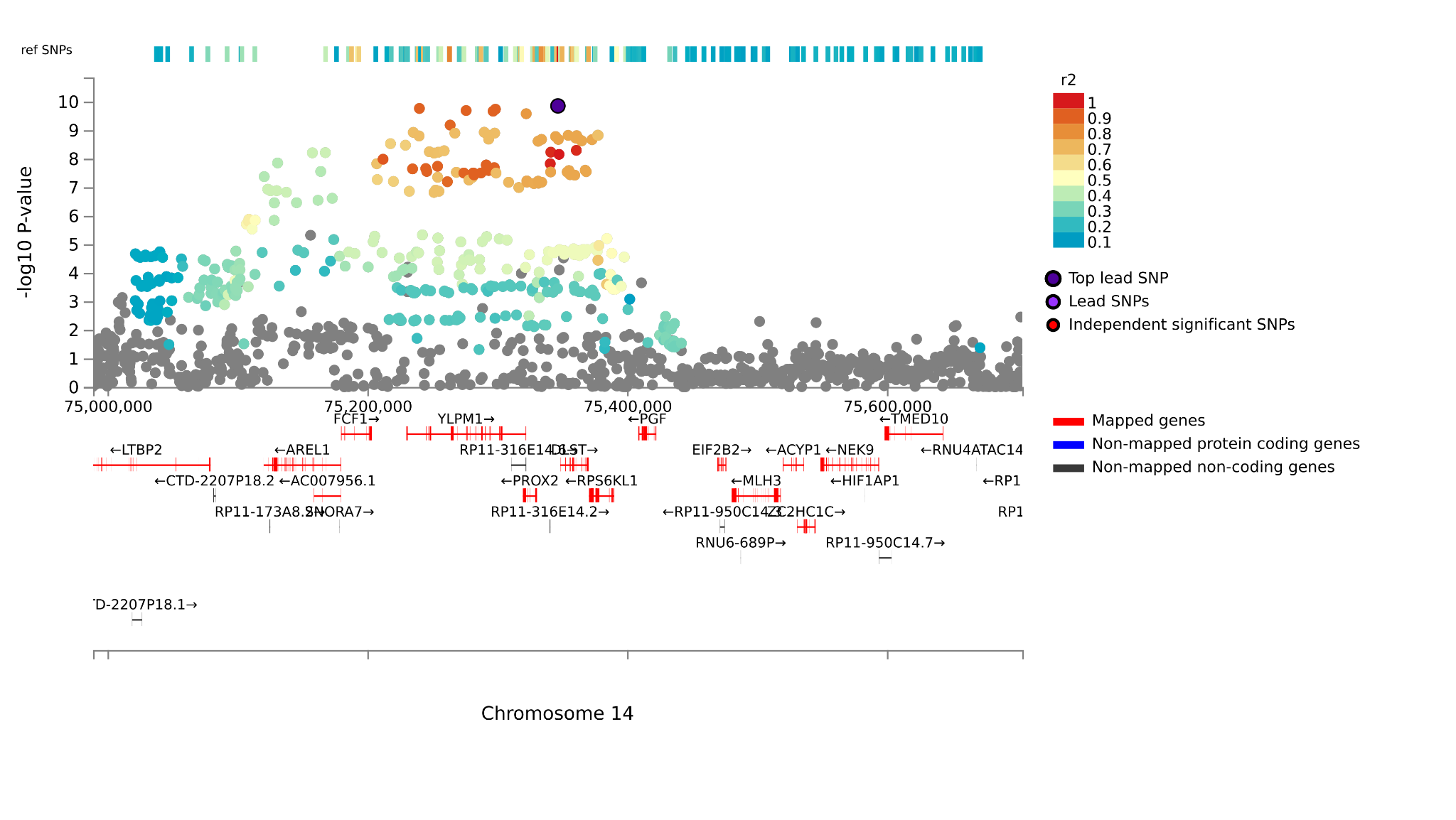

***rs112484789***

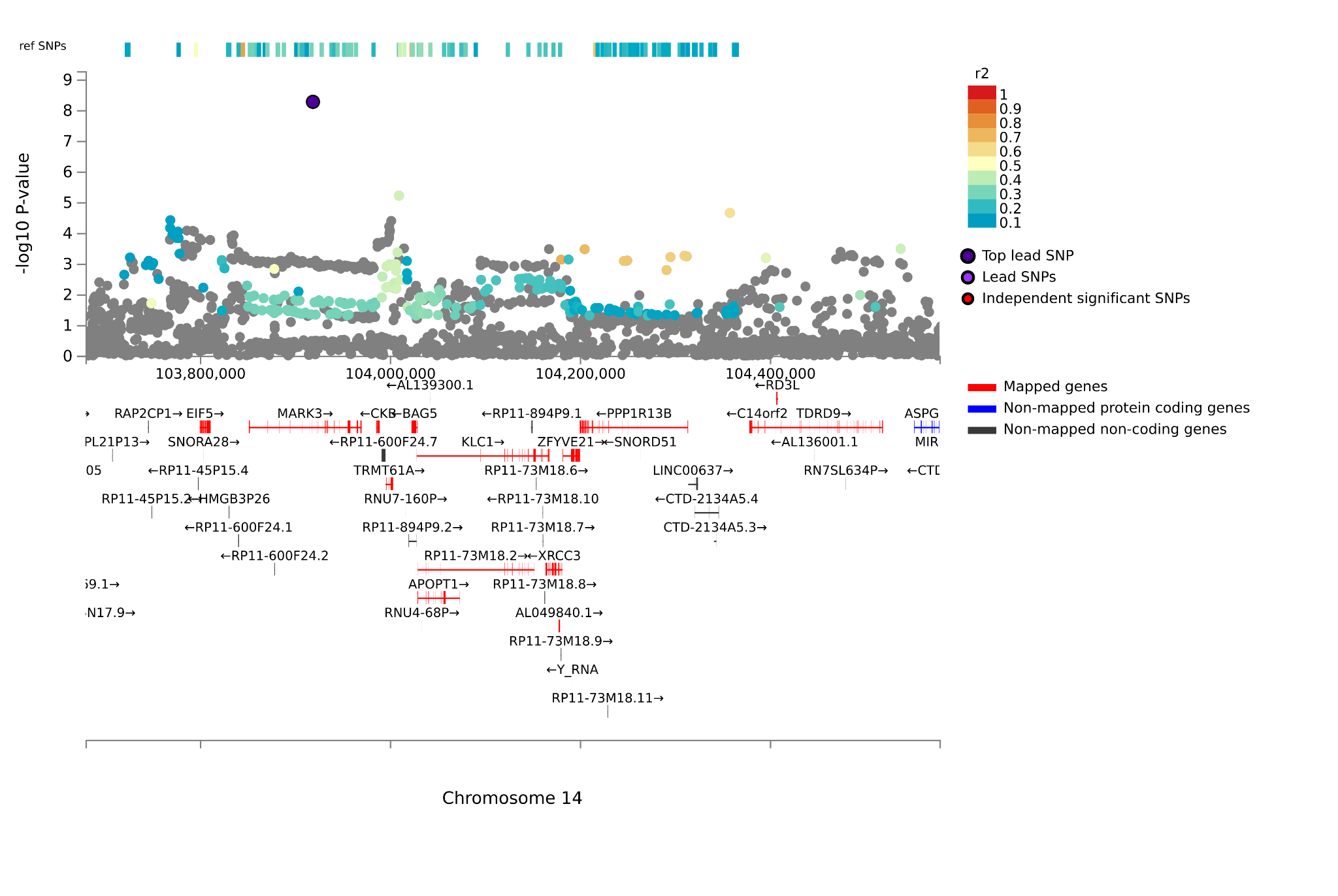

***rs7204169***

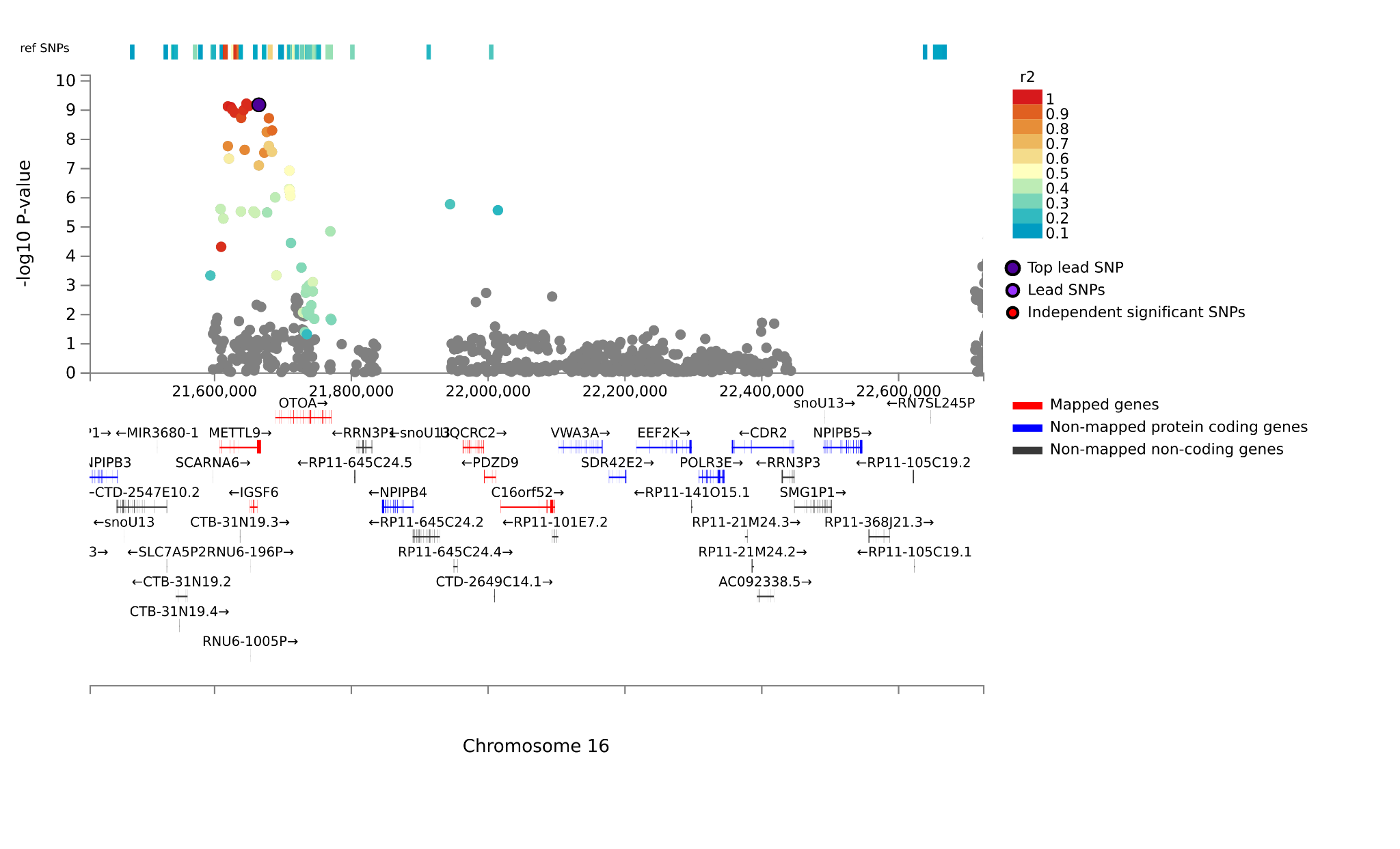

***rs4628973***

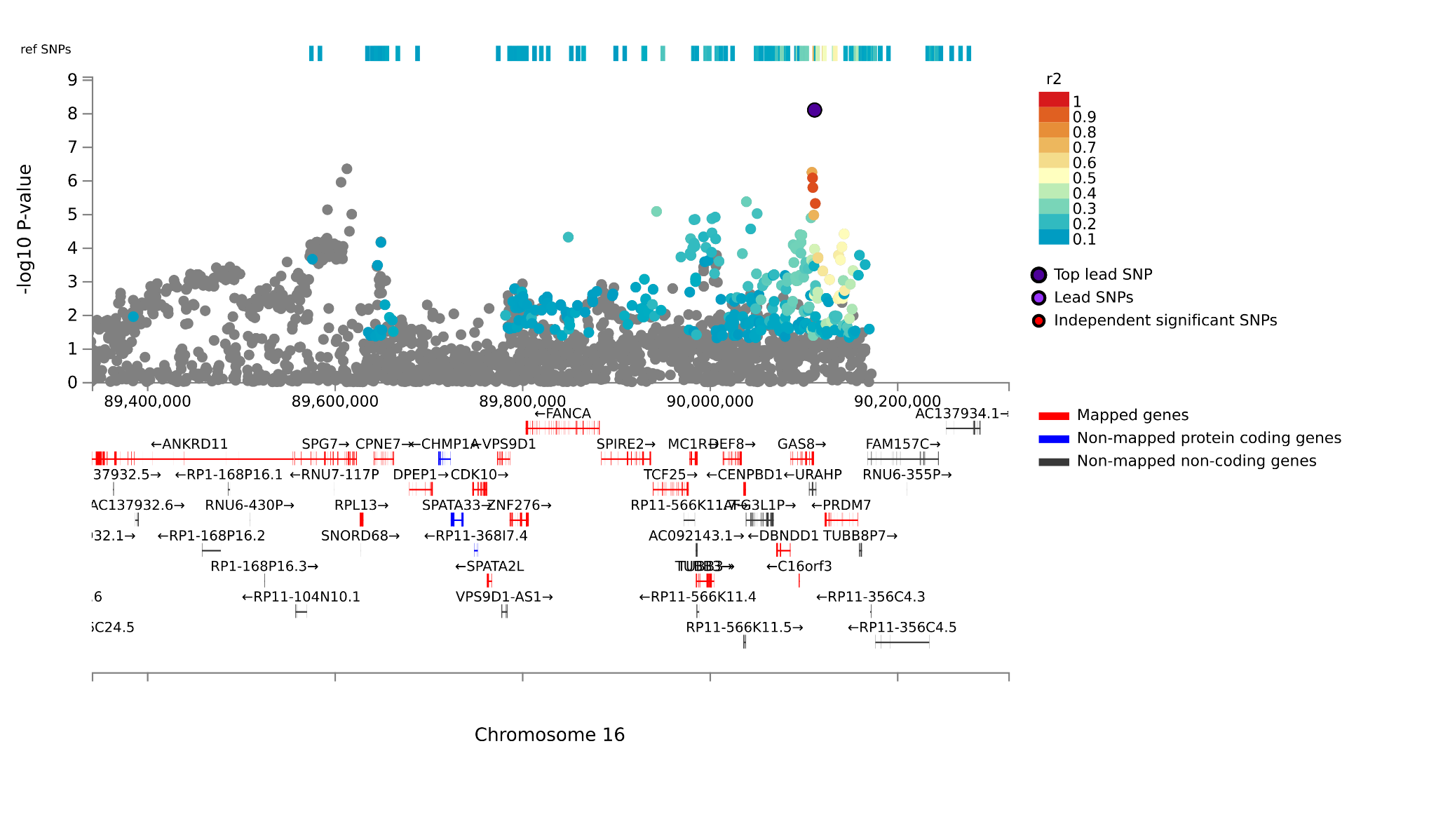

***rs34291370***

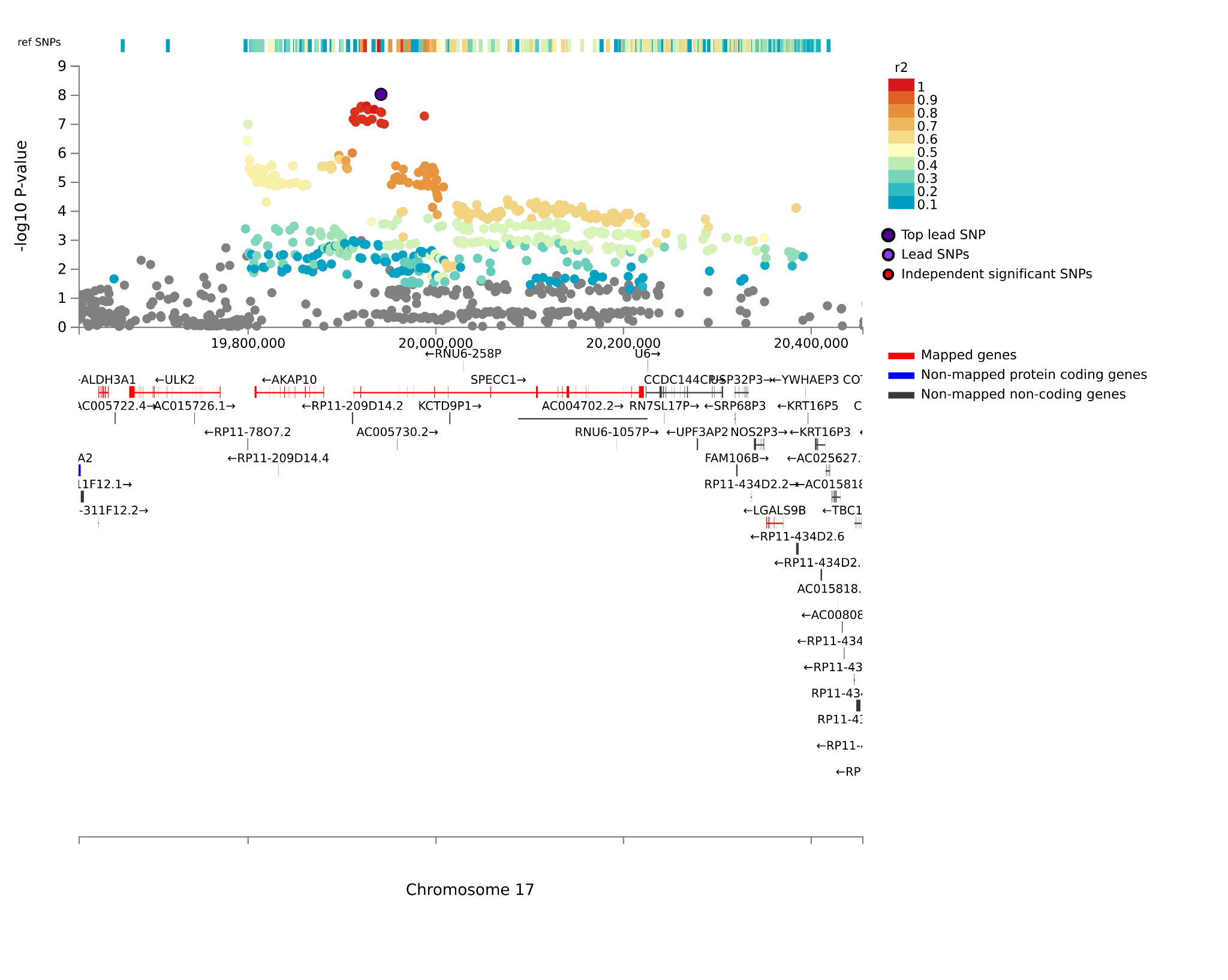

***rs12607755***

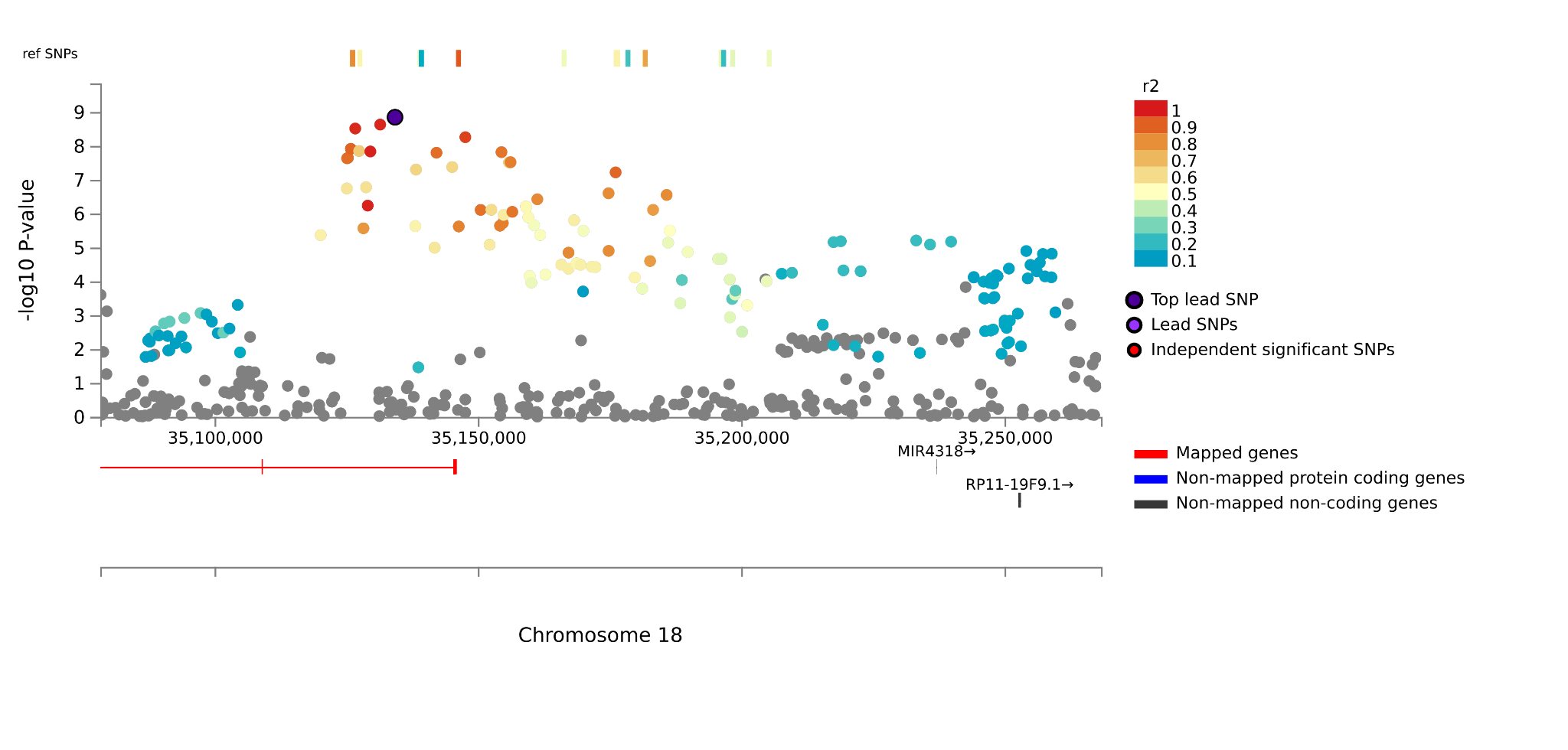

***rs17408393***

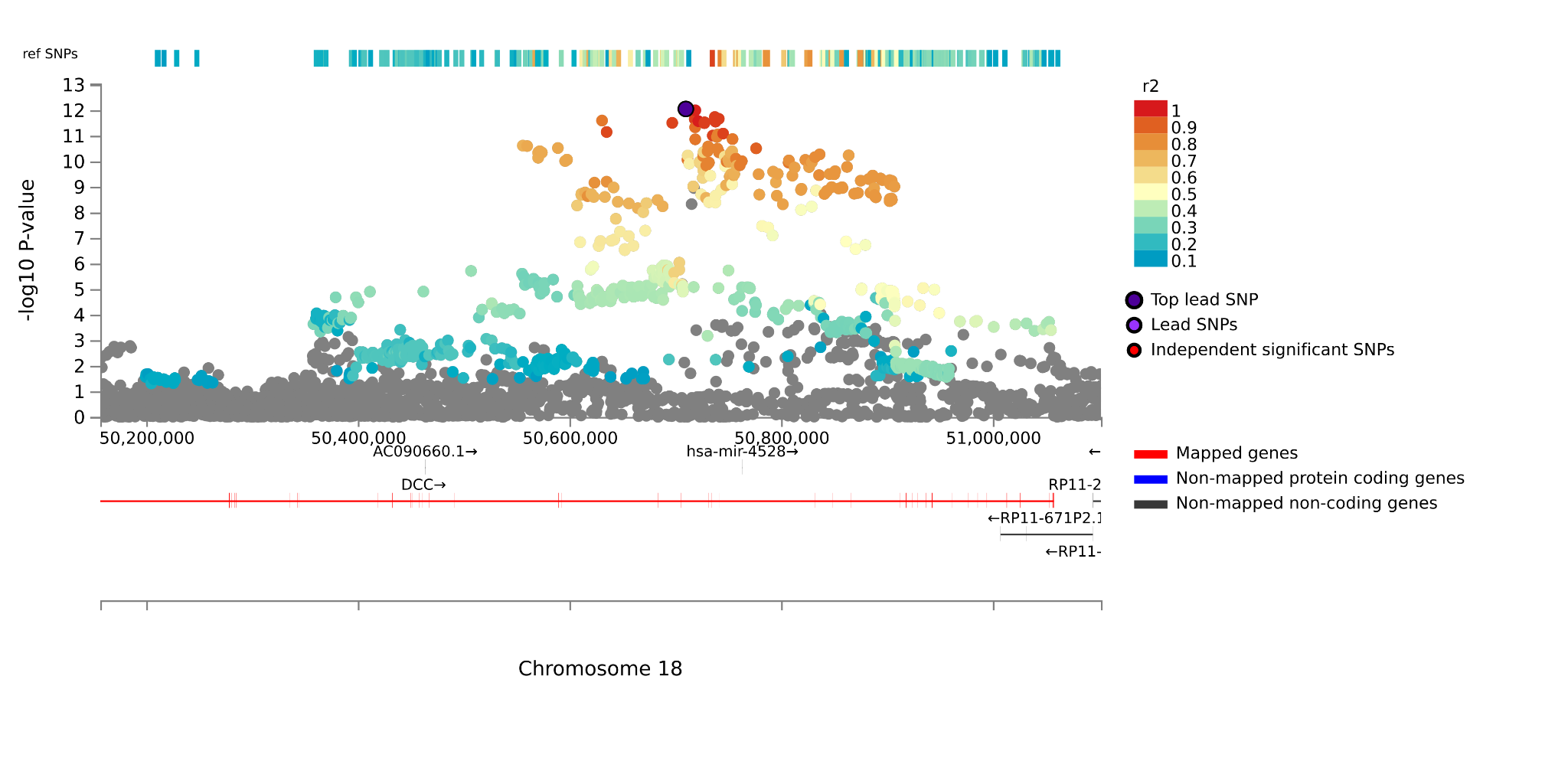

***rs6082358***

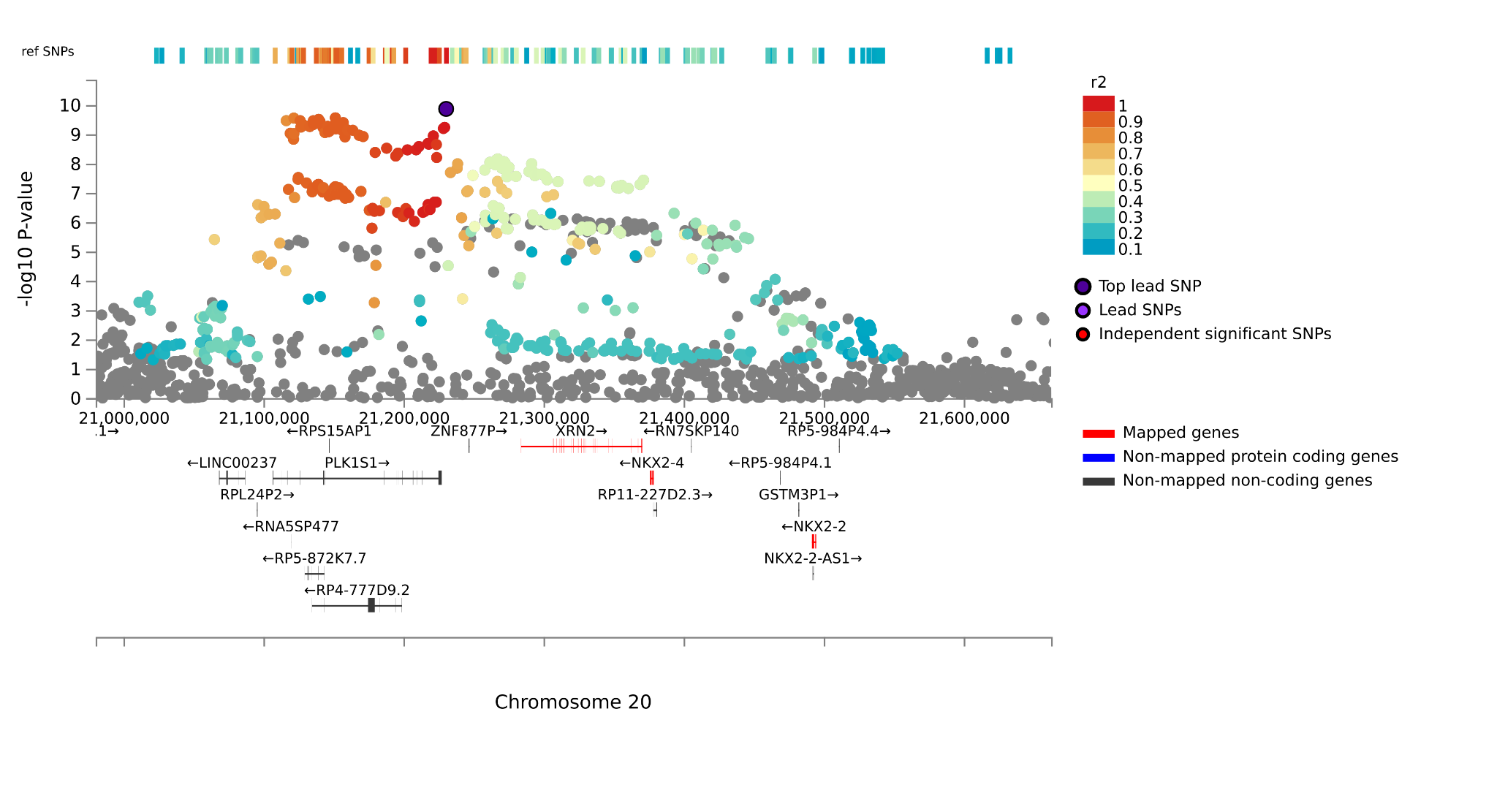

***rs12482317***

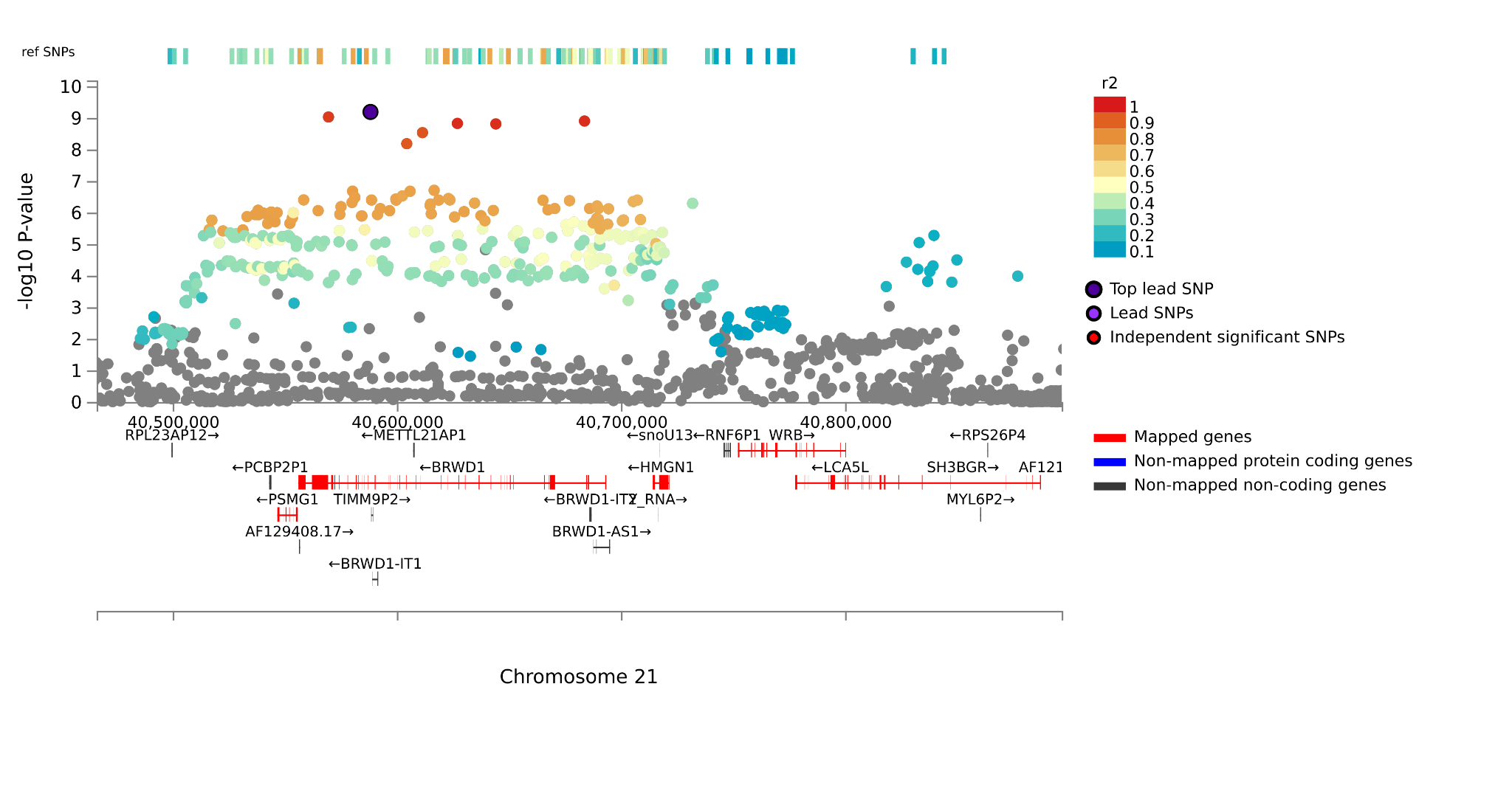

***rs2267158***

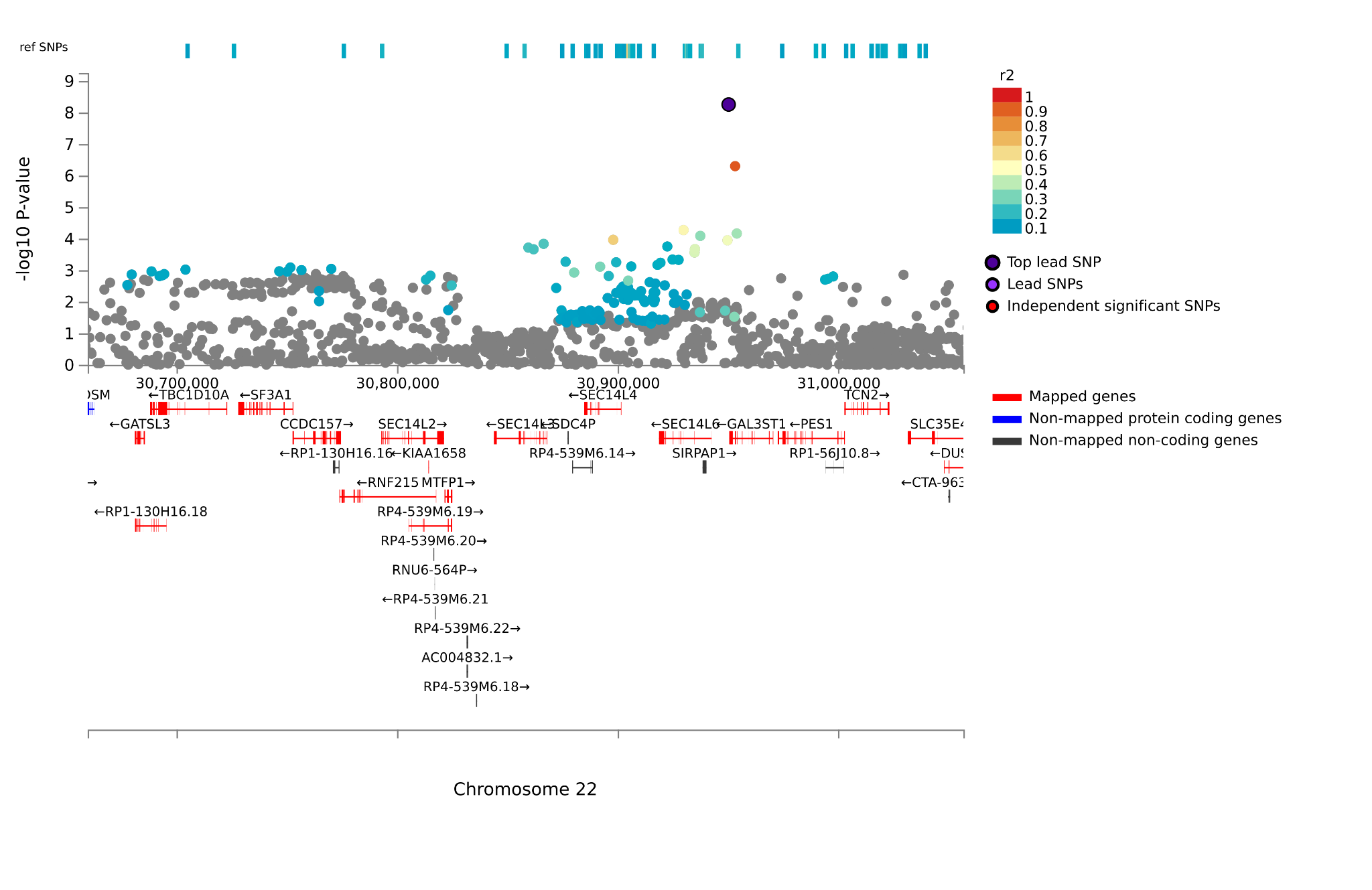

***rs2024567***

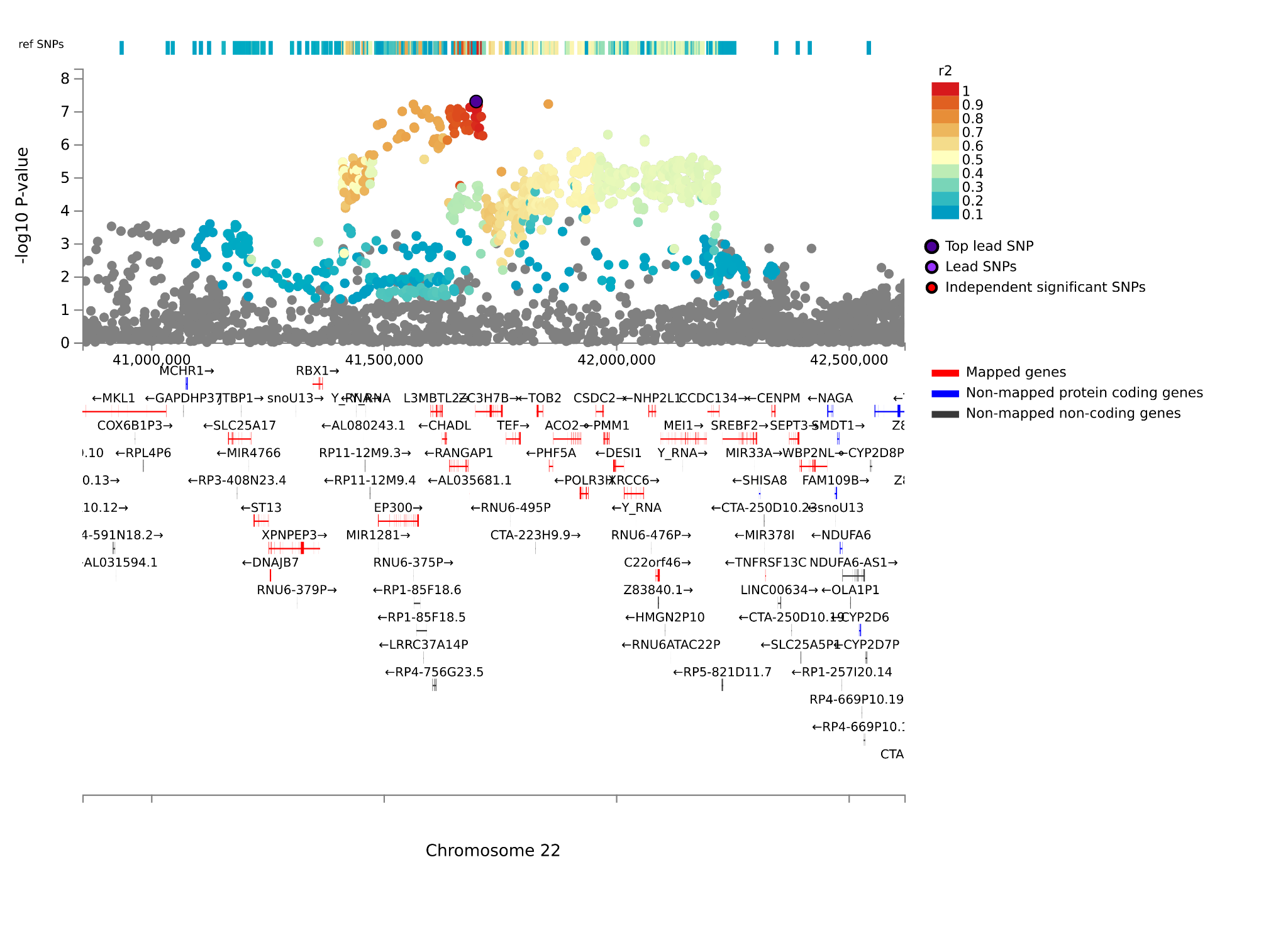

***rs6633421***

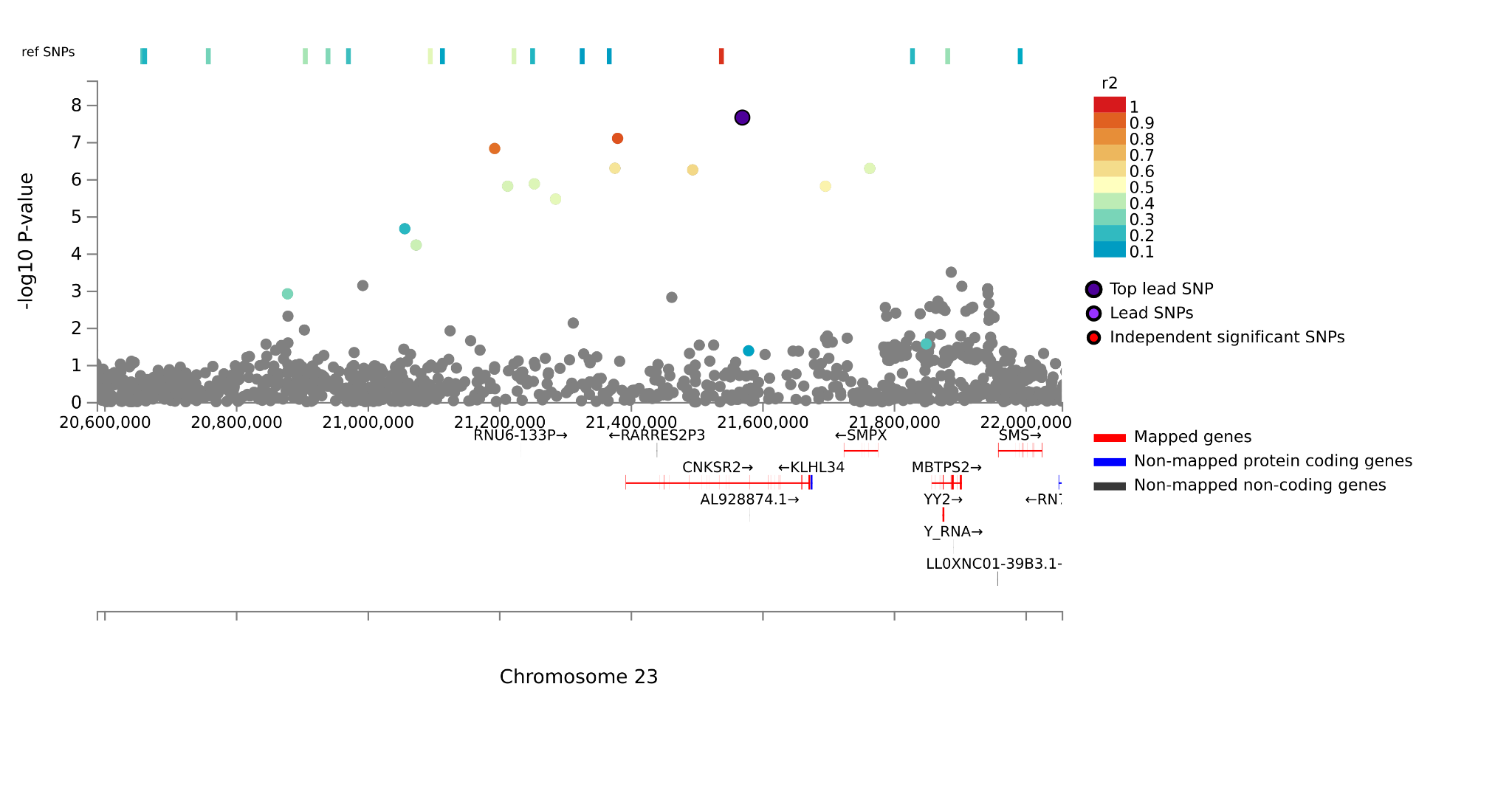

#### Analysis with controls screened only for panic

The meta-analysis was repeated with controls screened only for panic phenotypes (‘simple’ screening), rather than depression and anxiety disorders more broadly (‘super’ screening, resulting in 403,610 controls for broad and disorder, and 394,206 for attacks. The number of cases was unchanged (panic broad N = 90,840, panic disorder N = 41,721, panic attacks N = 49,119). The genetic correlations between the simple and super screened phenotypes were approximately unity at 0.99 (SE = 0.003), 1.00 (SE = 0.002), and 0.99 (SE = 0.006) for panic broad, disorder, and attacks, respectively. In this less well powered simple analysis, we found 20 genome-wide significant loci for broad, 11 loci for disorder, and 2 for attacks (Figure S2; Table S2).

For the panic broad lead variants, 14 replicated across the two screening methods; 10 of the 11 loci significant only for the main anxiety and depression screened phenotype were underpowered in the panic-screened version; the 6 loci significant specifically for panic-screened showed the same direction of effect in both analyses, with significance values ranging 1.5x10^-7^ - 9.9x10^-6^ for the anxiety and depression-screened.

For the 11 panic disorder loci, all were replications from the main screened analysis. The 4 loci significant only in the main screened analysis were all underpowered in the panic-screened version. The 2 loci for panic attacks were replications from the main screened analysis. The 3 loci significant only in the main screened analysis were all underpowered in the panic-screened phenotype. Finally, genetic correlations performed in LDSC showed a similar pattern of associations as the screened analysis (Tables S17-19; Figure S12).

#### Twin-based heritability

Twin heritability estimates were derived using the OpenMX^24^ version 2.22 package in R. Univariate liability threshold ACE twin models were fitted to panic broad (2720 twin pairs, 1121 MZ and 1599 DZ) and disorder (1570 twin pairs, 677 MZ and 893 DZ) data from the Twins Early Development Study (TEDS). The classic twin ACE model partitions the phenotypic variance into additive genetic effects (A), shared/common environmental effects (C) and non-shared environmental effects (E). Higher phenotypic similarity within MZ twin pairs, in comparison with DZ twin pairs, can be attributed to MZ twins’ higher genetic similarity. Non-additive genetic effects, such as dominance (D) are indicated if MZ correlation is twice higher than DZ correlation. Liability threshold twin model estimates the correlation in liabilities separately for MZ and DZ pairs from their count data, and variance decomposition (A, C, E) can be applied to the liability of the trait. Correlations in liability are determined by path model, and to estimate the heritability of the liability. ACE model fit was determined by comparing the -2ll statistics with that of the fully saturated model, with a non-significant p-value indicating good model fit.

Comparison of MZ and DZ correlations (Supplementary Table 28) indicated both panic disorder and panic attacks are heritable, with heritability estimates of 65% (CI: 57%- 72%) panic disorder and 41% (CI: 35% - 46%) for panic attacks. MZ correlations were more than double DZ correlations, suggesting the estimates reflect both additive and non-additive genetic effects. There was no evidence of shared environments on liability to panic disorder nor attack, with non-shared environments explaining 35% (CI: 28% - 43%) and 59% (CI: 53% - 64%) of the variance in liability to panic disorder and attacks, respectively.

#### Fine-mapping

Functionally-informed fine-mapping identified one putatively causal single variant, rs112484789, an intronic SNP in the Microtubule Affinity Regulating Kinase 3 (*MARK3*) gene, which had PIP=0.958 for causality in the main panic attack GWAS (Supplementary Table 6). Among other roles, this variant is a brain sQTL for the kinesin light chain 1 (*KLC1*) gene, the product of which has been implicated in (amongst many processes) neurodegeneration in Alzheimer's disease^25^. However, the variant is the only genome-wide significant variant in the locus. While this is consistent with the surrounding pattern of linkage disequilibrium, this finding should be interpreted cautiously until replicated in future analyses.

In addition to rs112484789, five small credible causal sets (<10 variants) were identified (Supplementary Table 6). This included sets around rs2267158 (chromosome 22, panic broad and panic attack main, in both the primary super and sensitivity simple screened controls analyses), rs1806153 (chromosome 11) and rs4628973 (chromosome 16, panic broad in both the primary super and sensitivity simple screened controls analyses), and rs9971210 (chromosome 10) and rs62223042 (chromosome 21, panic broad in both the primary super and sensitivity simple screened controls analyses). The size of the credible set around rs2267158 varies across analyses, but consistently prioritises rs2267158 and rs2267161, the latter of which is a conservative missense variant in the Galactose-3-O-Sulfotransferase 1 (*GAL3ST1*) gene, the product of which is key to the synthesis of sulfatide, an important component of myelin^26^. The credible set around rs1806153 lies broadly across Paired Box 6 (PAX6), a master regulator involved in early neurogenesis [PMID 41744767], although the PIP is diffuse across the set, making it difficult to determine a clear mechanistic link to PAX6 rather than other nearby genes. A similar statement can be made for the set around rs9971210, which lies over the MLLT10 Histone Lysine Methyltransferase DOT1L Cofactor (MLLT10). This contrasts with the set around rs4628973, where the PIP is concentrated in the lead variant, an intronic SNP in the Urate Hydrolase Pseudogene (URAHP) and an eQTL and sQTL for numerous genes. Again, specific mechanistic insight is difficult to determine. Finally, the set around rs62223042 lies over Bromodomain And WD Repeat Domain Containing 1 (BRWD1), although the lead variant is perhaps more convincing as an eQTL and sQTL of Proteasome Assembly Chaperone 1 (PSMG1), the relevance of which to panic is unclear.

#### MAGMA and Biological pathway analyses

MAGMA^27^ was used to perform gene-based and gene-set analysis, identifying biological pathways and gene ontology terms enriched for associations with panic disorder. This approach provides insight into potential biological mechanisms underlying the genetic associations.

**Gene-based associations** using MAGMA^27^ and a significance threshold corrected for 19,886 genes tested (0.05/19886 = 2.514x10^-6^) revealed 38 genes across 23 loci that were associated with panic broad. For panic disorder, 19 genes were associated across 15 loci. For panic attacks, 5 genes were associated across 4 loci. **Gene-set analyses** identified several significant gene-sets for panic broad and disorder, but none survived Bonferroni correction.

#### Single-cell analyses for panic attacks

To further investigate the function of the visceral neuronal populations from the foetal gene expression atlas, we first clustered them with all other cell types based on their transcriptional signal. This analysis highlighted their similarity to other neuronal types from the PNS/CNS (Supplementary Fig 7), dissuading any concerns of mislabeling in the peripheral organs.

Based on the findings of differing neuronal transcriptional signals (Supplementary Fig 7), we inferred that these visceral neurons are likely present in the peripheral tissue of the lungs and heart projecting inward but also within the organ themselves which can be seen in spatial transcriptomic data using visceral cell type gene markers in a published spatial transcriptomic data from the sinoatrial node^28^ (Supplementary Fig 8).

Based on these findings, we are confident that the observed peripheral neuronal enrichments are genuine and contribute to the PNS’s involvement in panic disorder. However, differential enrichment analyses revealed that while this peripheral neuronal signal remained significant when compared to non-neuronal signatures (FDR = 0.022), PNS neurons did not reach significance when compared to CNS neurons (FDR = 0.95). This suggests that the enrichment signal may be driven by shared neuronal features rather than being specific to neuronal location (Supplementary Table 13).

Finally, we tested the cell-type-specific enrichment of other psychiatric disorders using the same foetal developmental single-cell atlas to determine whether these enrichments were unique to panic or common across psychiatric traits. MAGMA celltyping analysis revealed similar enrichments of CNS and PNS neurons across all tested psychiatric disorders, including the visceral neurons of the heart and lungs and the amacrine and ganglion cells of the retina (Supplementary Fig 11). To assess whether the magnitude of enrichment of each neuronal population was equivalent across disorders, we measured the proportion of heritability explained using an orthogonal approach, s-LDSC^29^, whose enrichment estimates are not directly affected by differing GWAS power (Figure 4). This revealed that, while eye neurons were most enriched in panic broad, significant associations were also observed in major depressive disorder, dimensional anxiety, schizophrenia and bipolar disorder, with the strongest evidence in schizophrenia and bipolar disorder.

We note that, of the eye cell types implicated in the primary whole-body analysis, only eye bipolar cell enrichment could be tested in an independent single-cell retinal dataset, and it did not replicate. Given the limited independent validation available for these retinal cell populations, the retinal findings should be interpreted with caution.

### Supplementary Figures

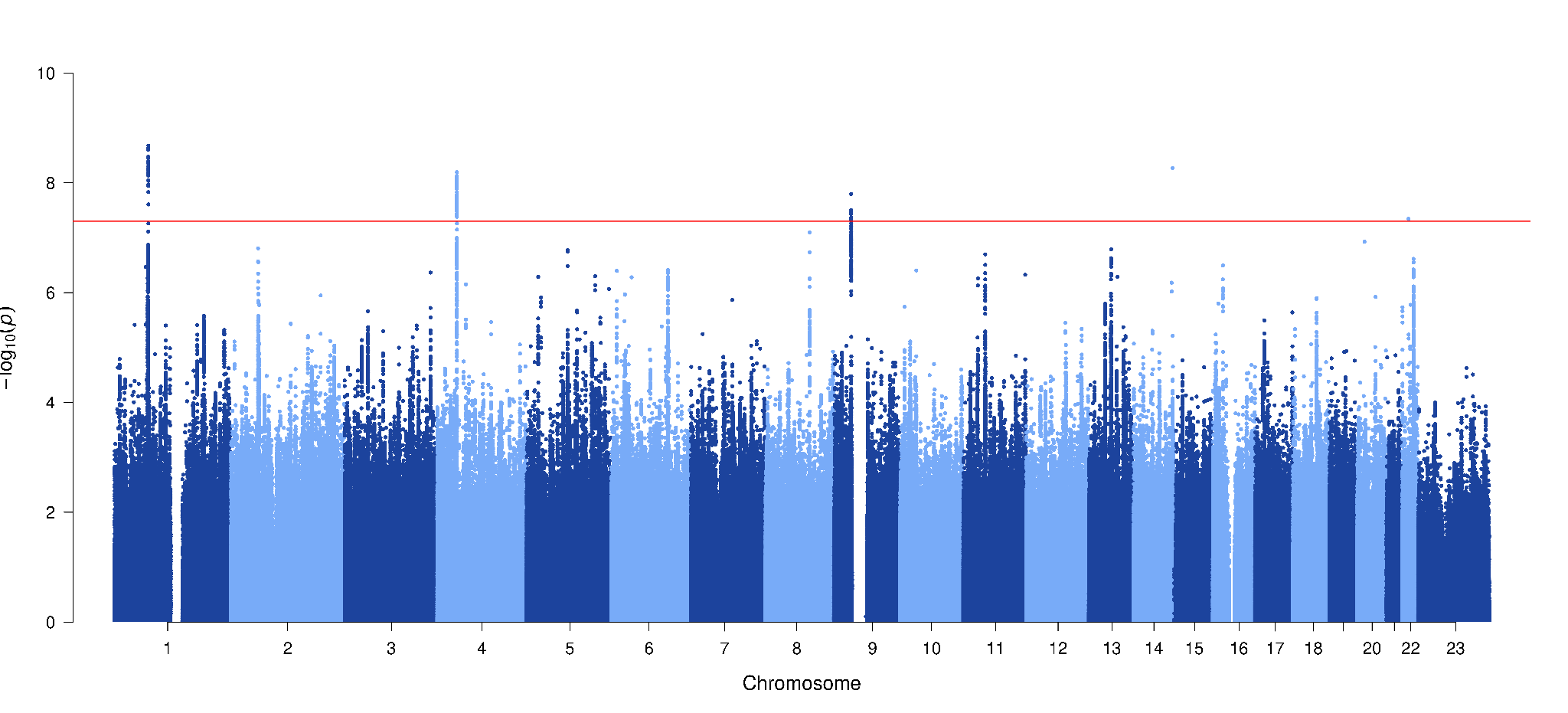

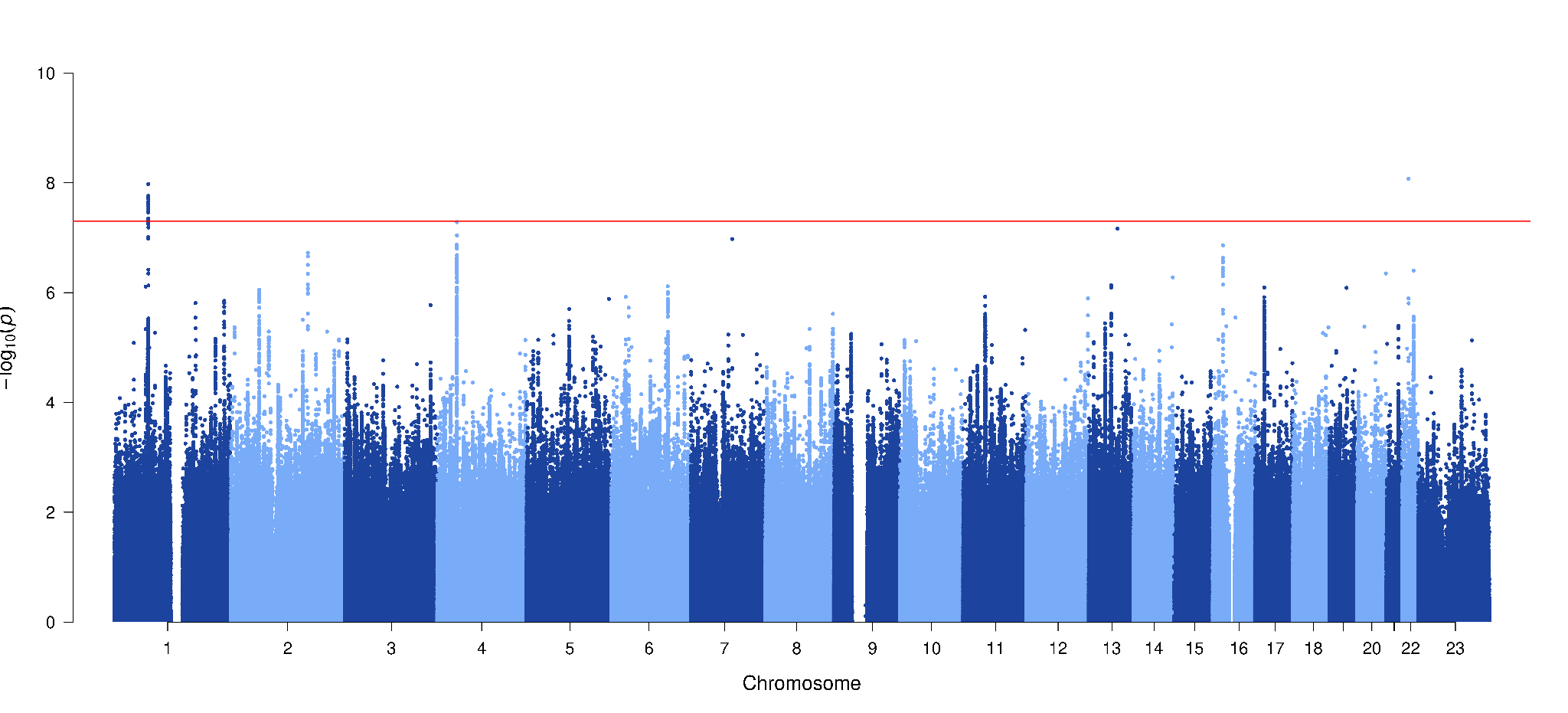

#### Fig S1. Manhattan plots of panic attacks using in our primary analysis with super-screened controls (top) and panic attacks with simple-screened controls (screened only for panic; bottom)

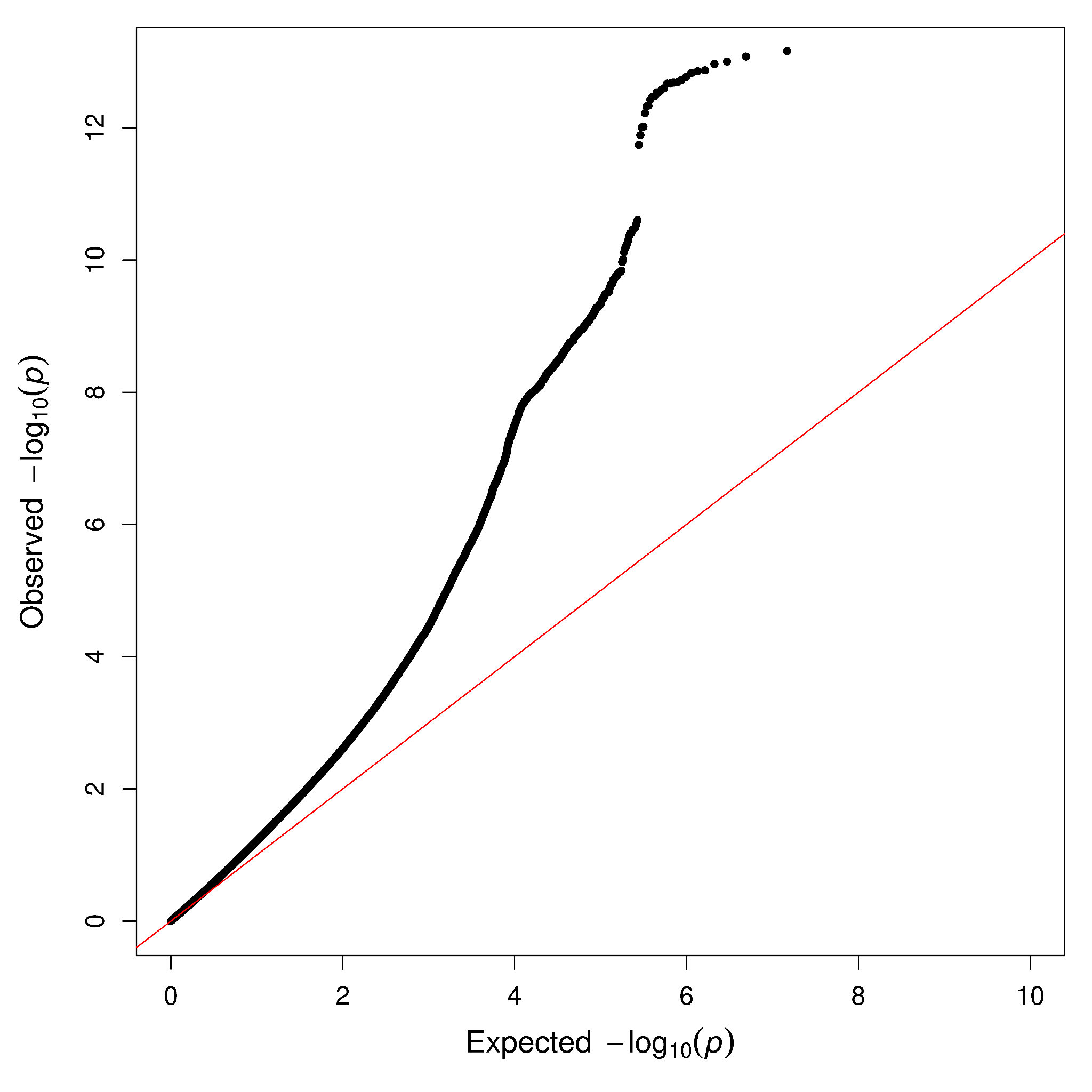

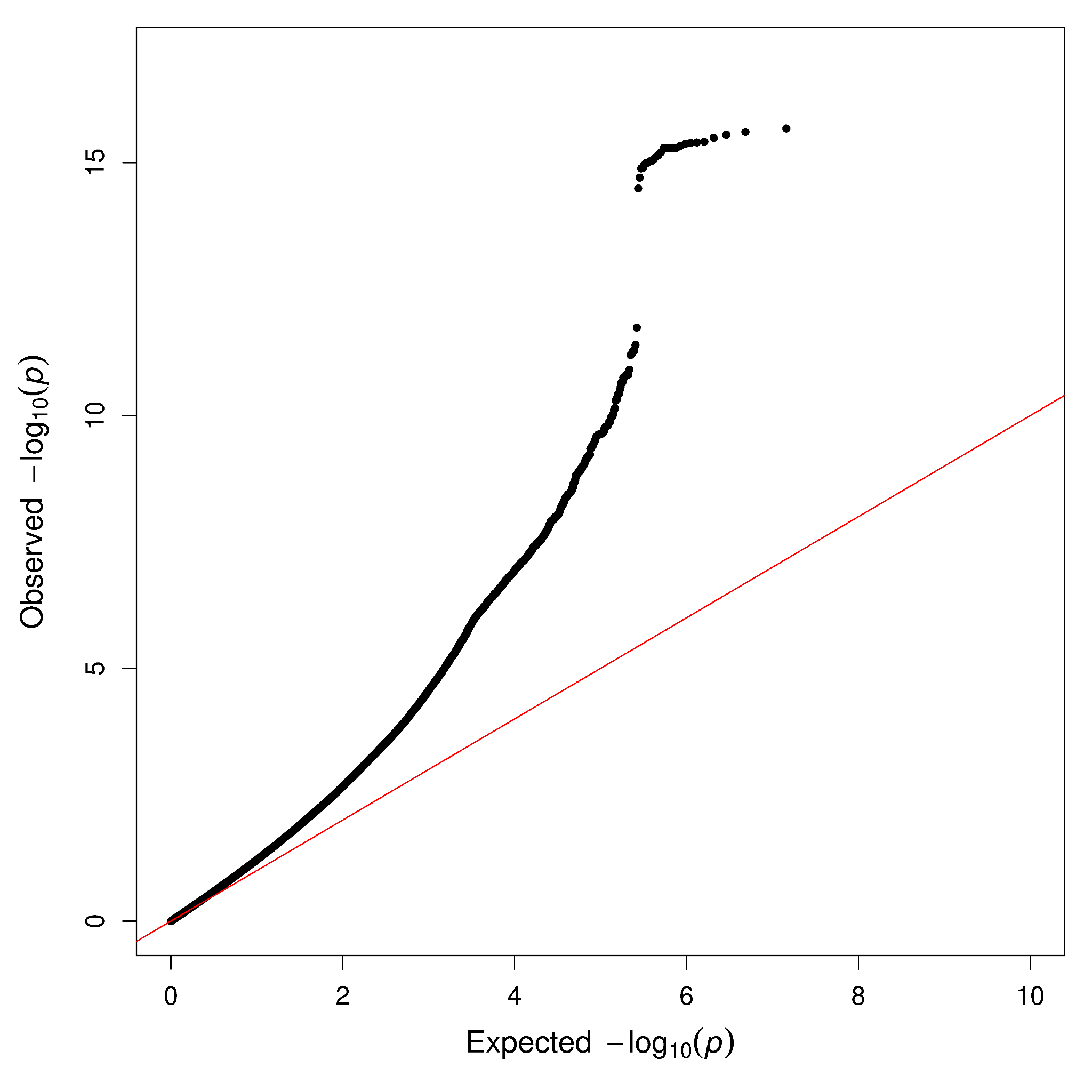

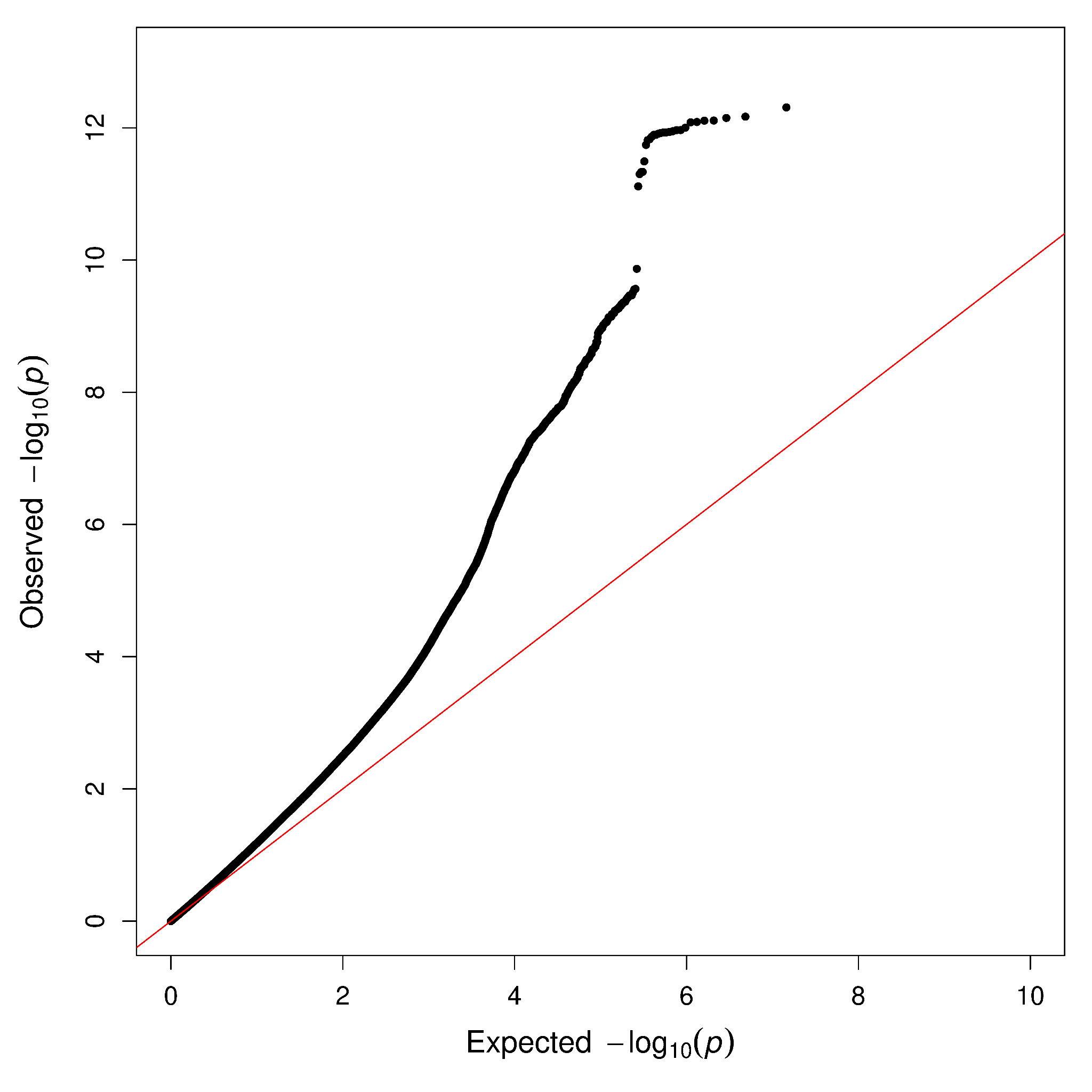

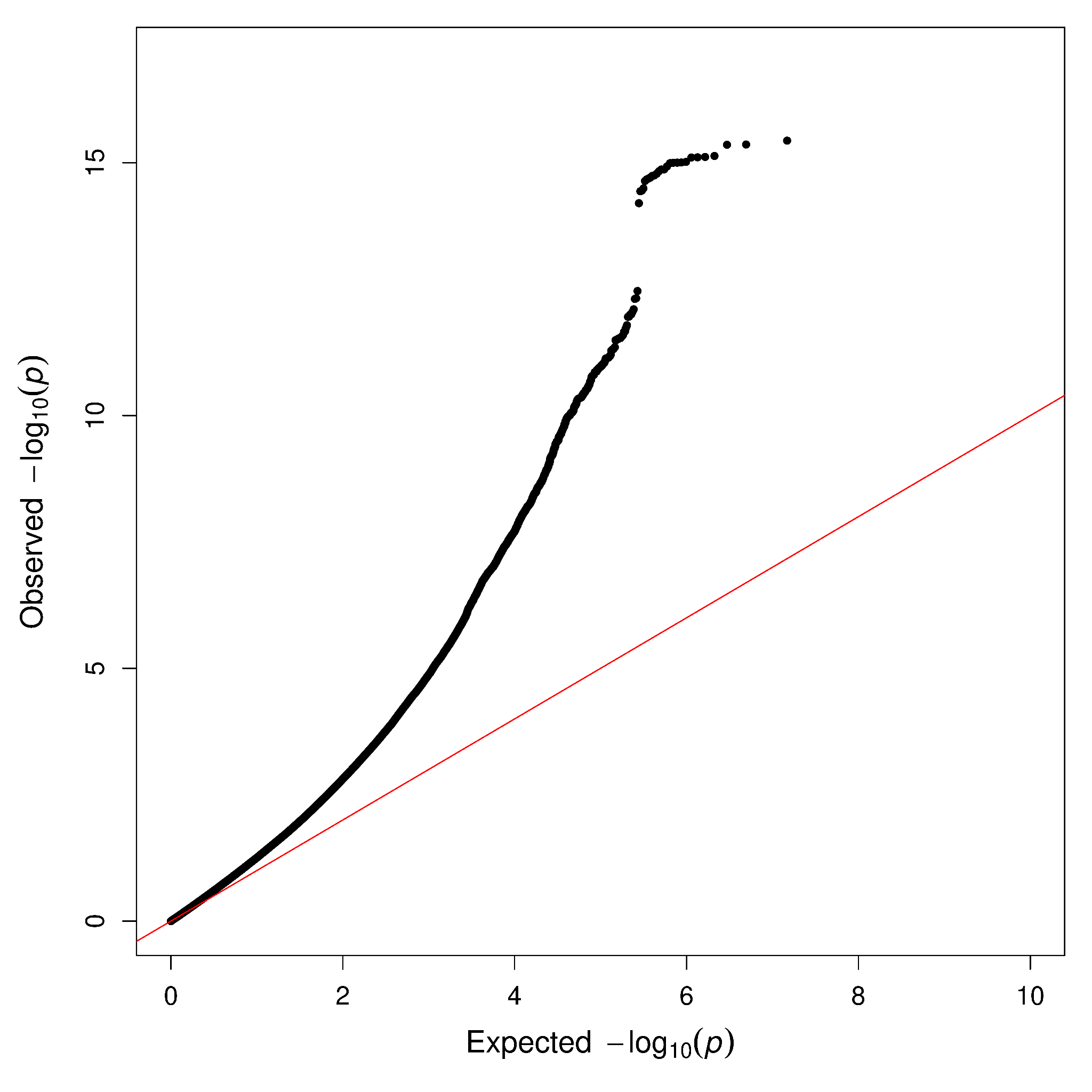

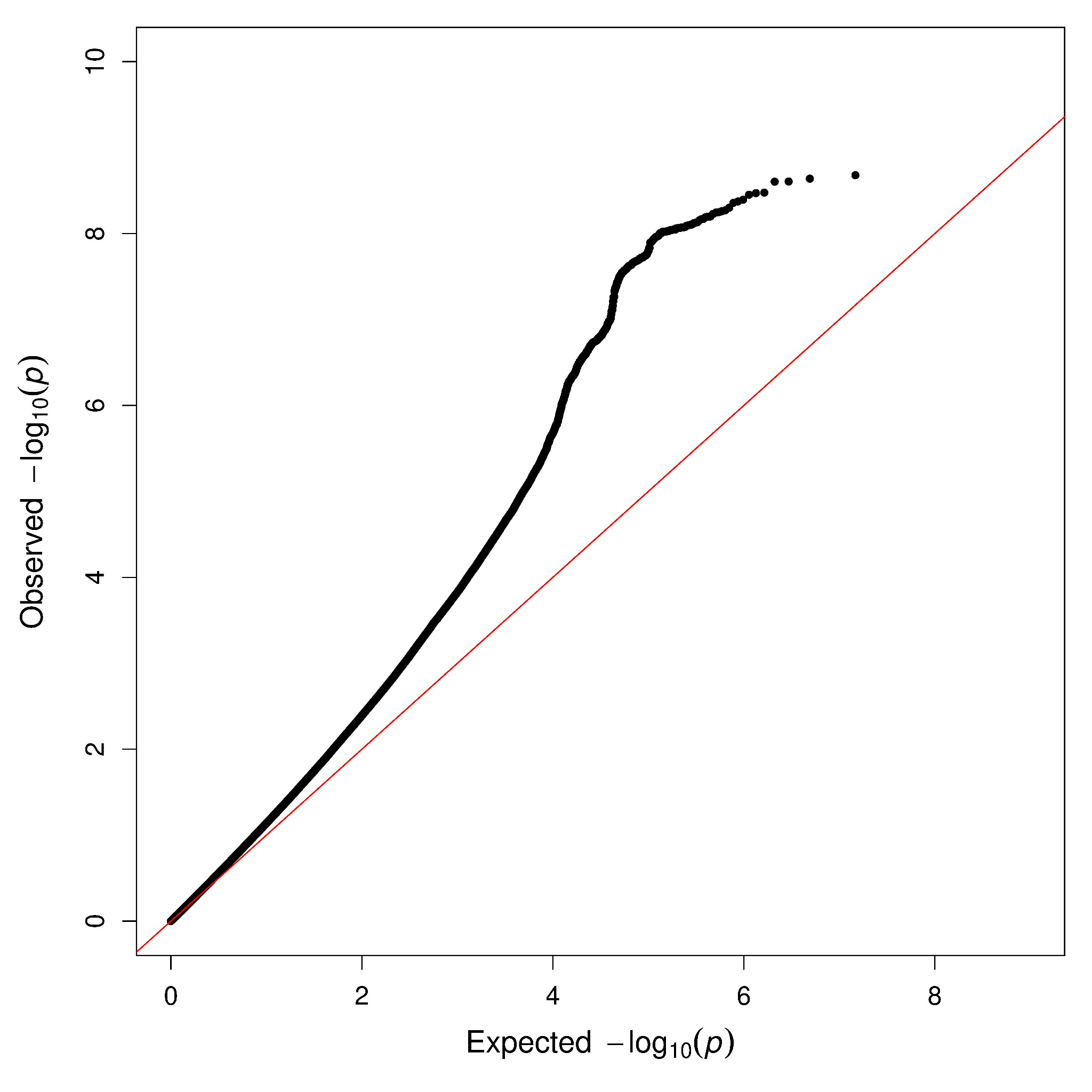

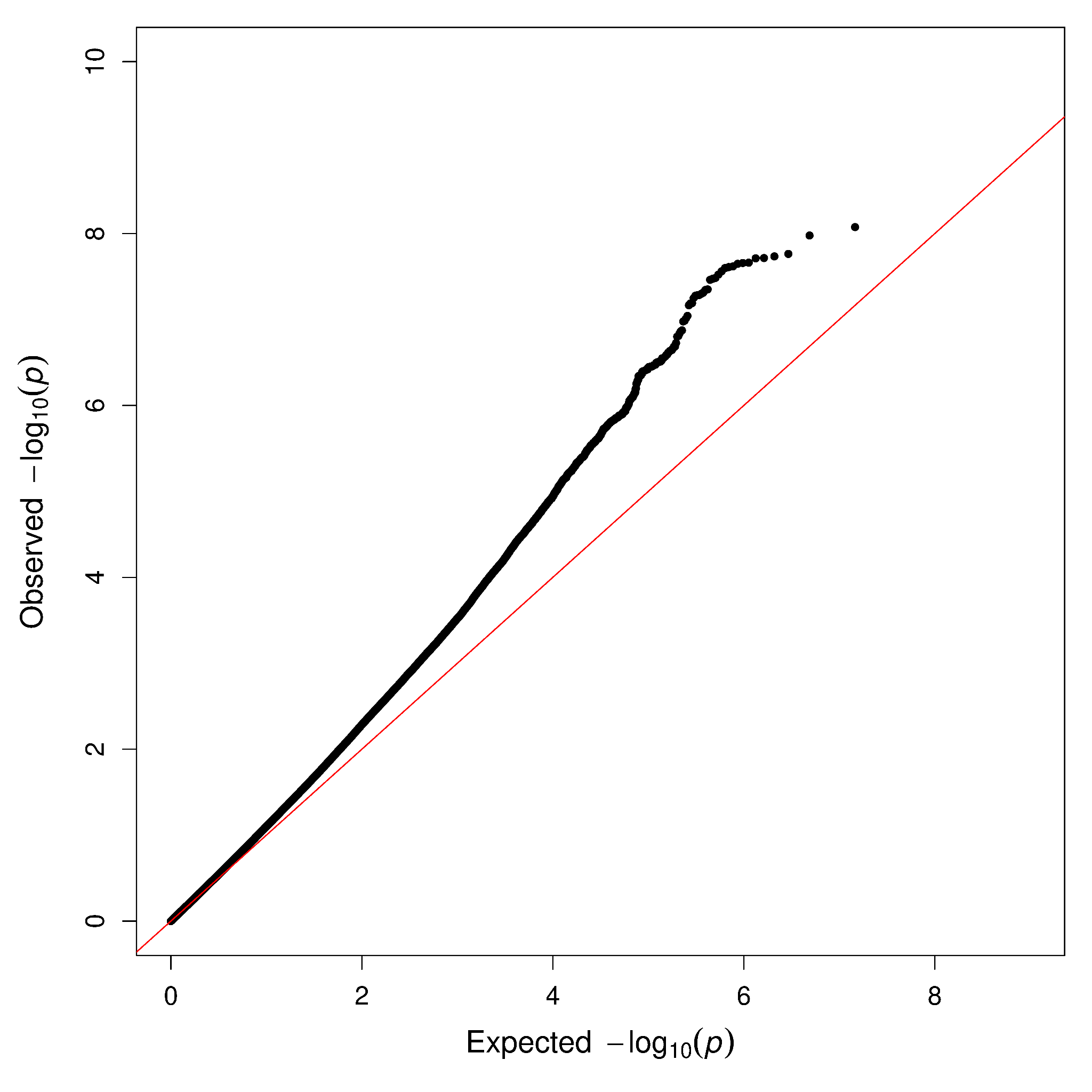

**Fig S2.** Quantile-quantile plots for panic broad (top), panic disorder (middle) and panic attacks (bottom) with controls screened for anxiety and depression (‘super screened’; left) or for only panic (‘simple screened’; right).

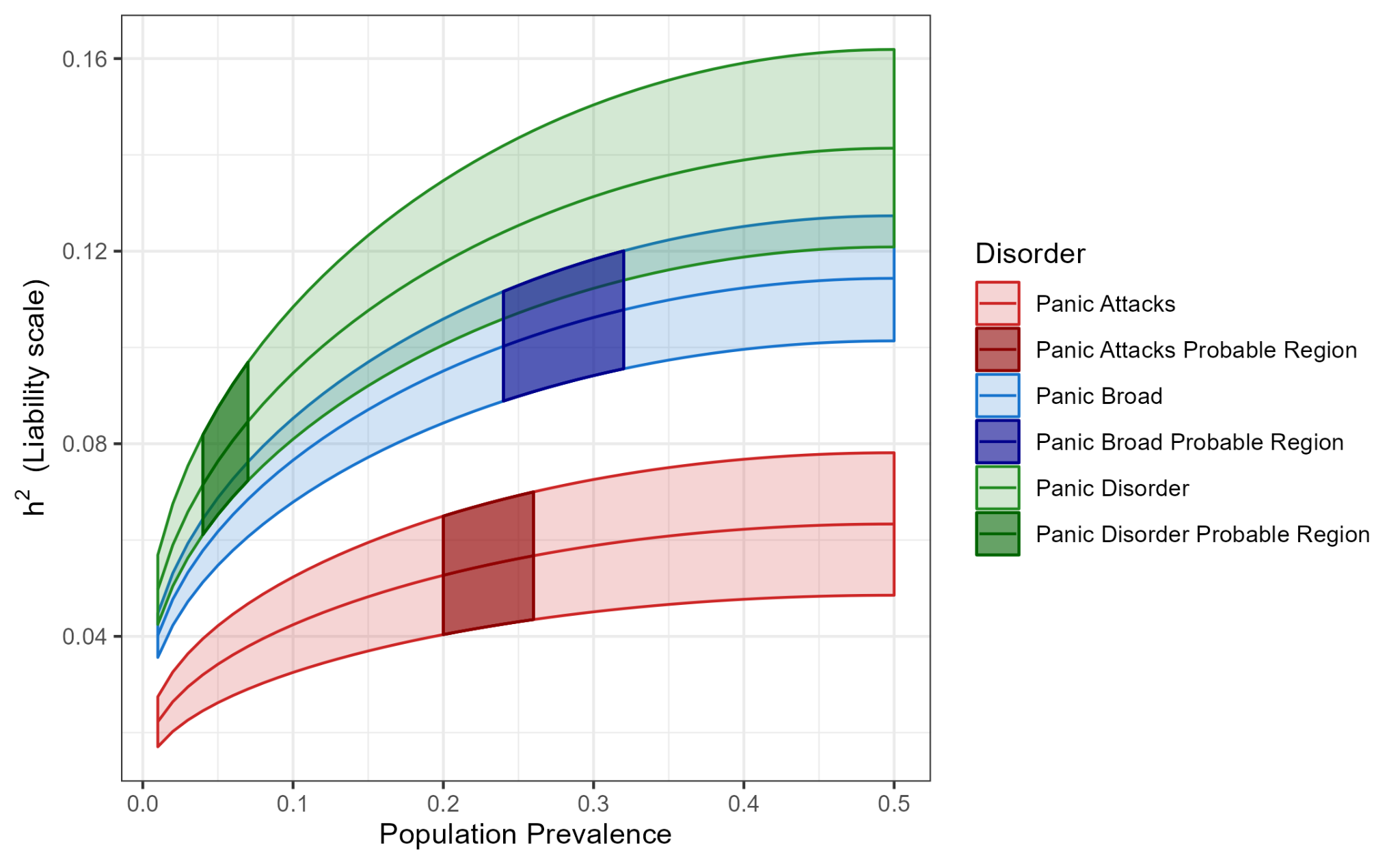

#### **Fig S**3: Rainbow plot of SNP-based heritability estimates for panic attacks, panic disorder and panic broad across a range of population prevalence estimates.

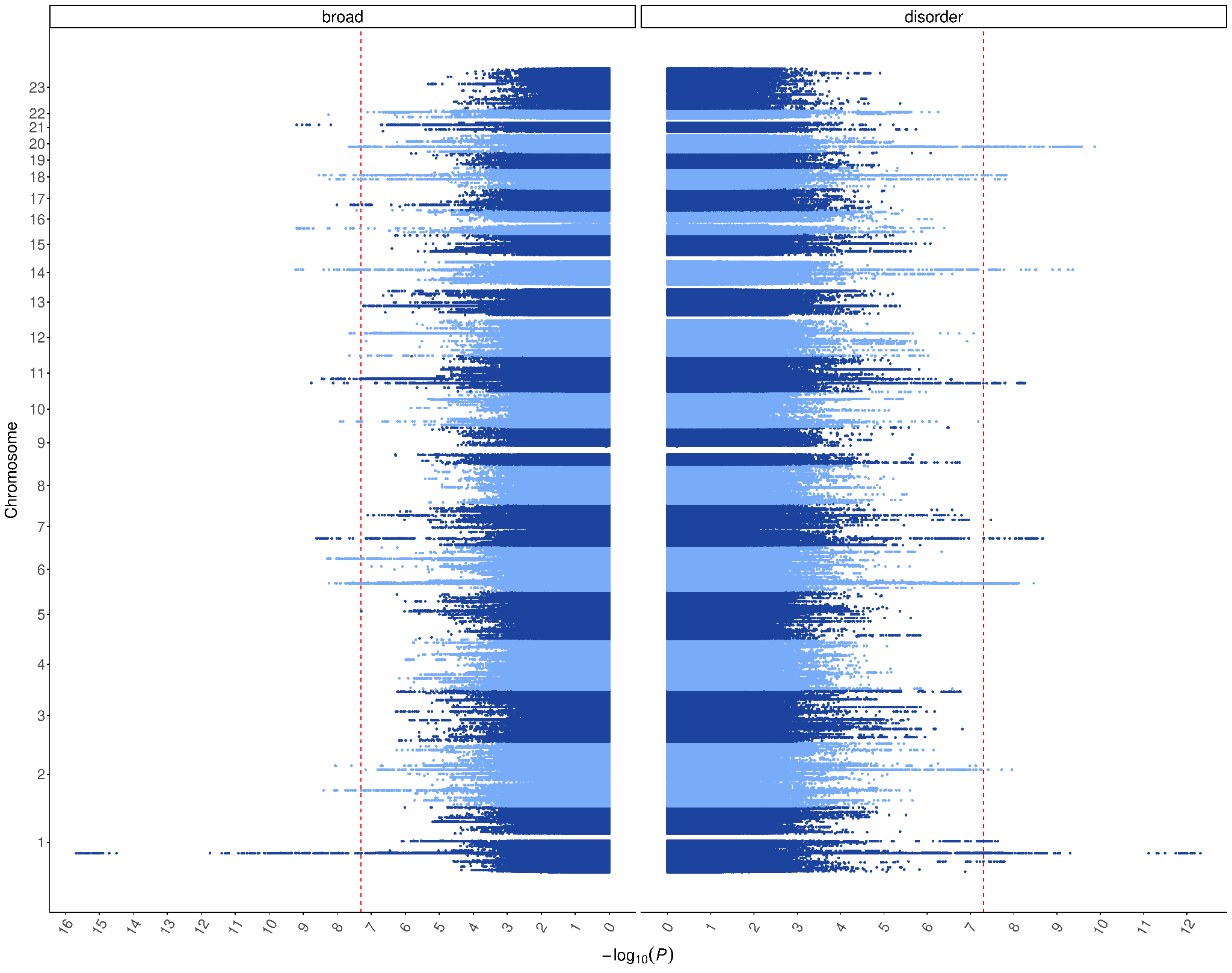

#### Fig S4. Mirror plot of the statistical significance of genome-wide variant associations for panic broad (left; case N = 90,840) and panic disorder (right; case N = 41,721) screened only for panic (common controls N = 403,610). The red vertical lines indicate the genome-wide significance threshold (p < 5x10^-8^).

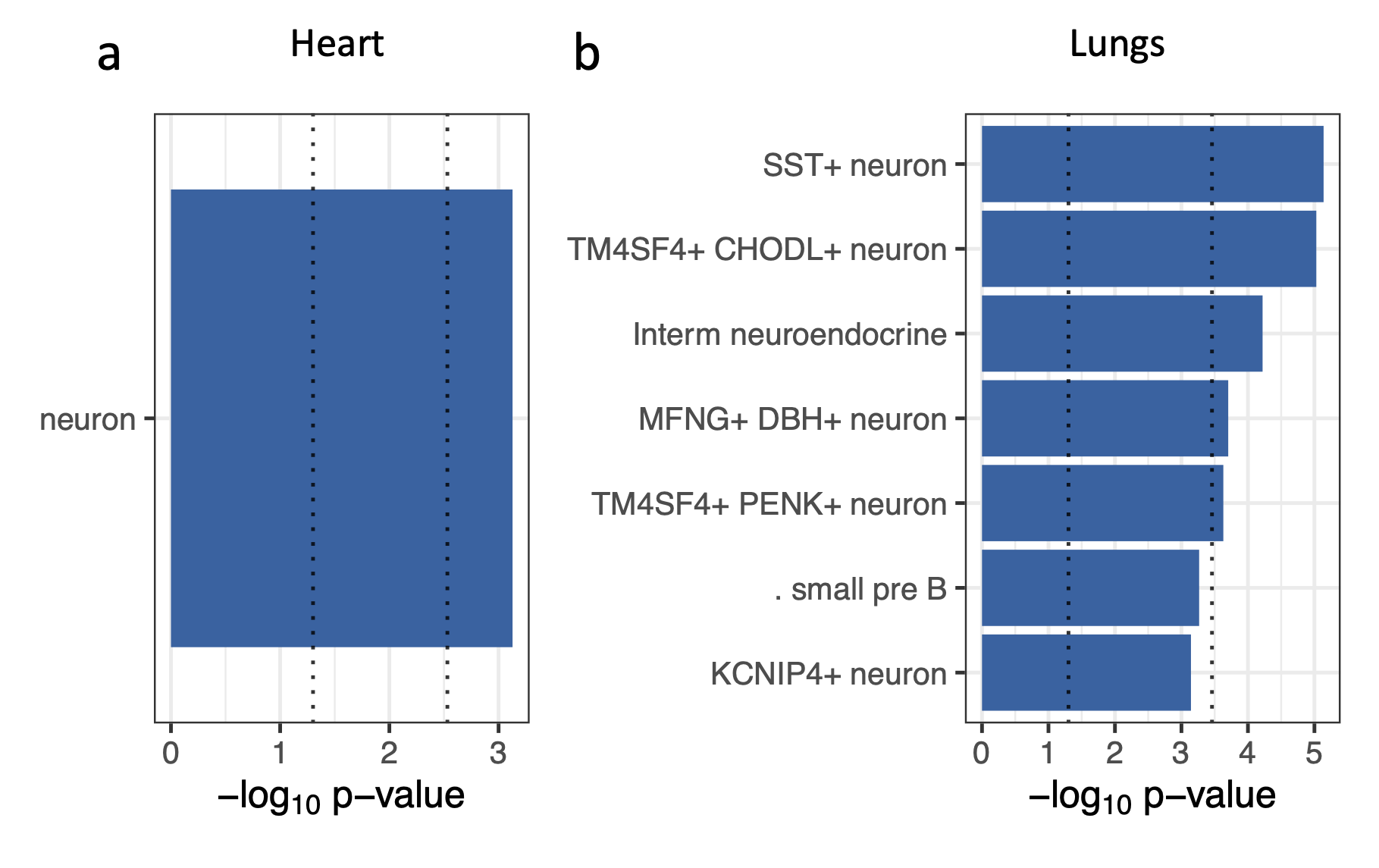

#### **Fig S5:** All cell type-specific, significant genetic enrichments (FDR < 0.05) for panic broad in the heart (a) and lung (b) single-cell transcriptional dataset. The MAGMA p-value enrichments are shown for linear regression. The black dashed vertical lines indicating p-value thresholds of 0.05 and Bonferroni adjusted p-value threshold of 0.0001. The y-axis shows the -log10 p-value where larger values indicate a greater level of significance.

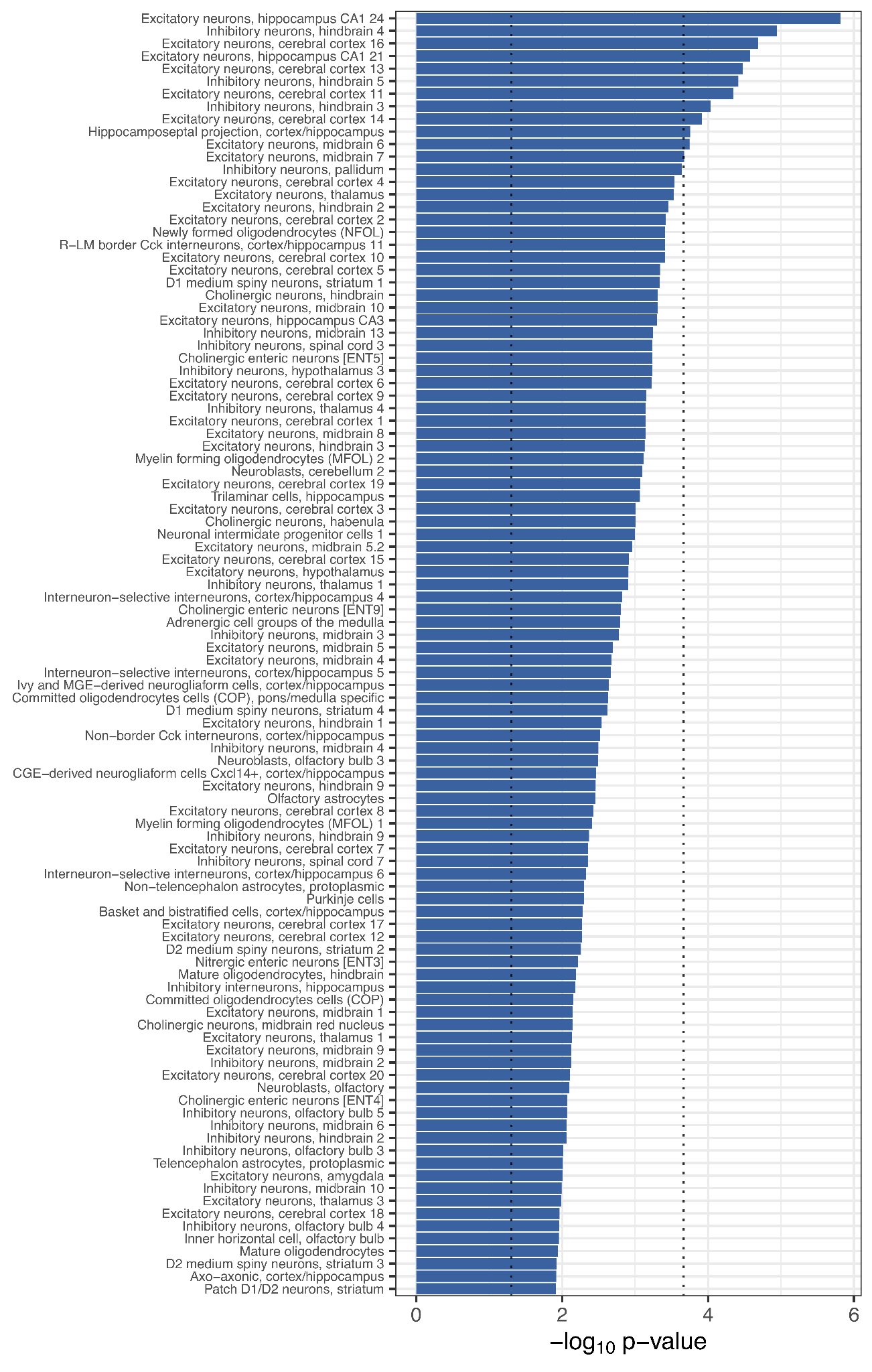

#### **Fig S6**: All cell type-specific, significant genetic enrichments (FDR < 0.05) for panic broad based on a mouse central nervous system single-cell transcriptional dataset. MAGMA gene set enrichment test p-value enrichments are shown for linear regression. Black dashed vertical lines indicate p-value thresholds of 0.05 and Bonferroni-adjusted p-value threshold of 0.0001.

The significant associations observed for lung and cardiac visceral neurons, which have highly related transcriptomes to CNS neurons (Fig S7) and appear to show adrenergic, acetylcholine and cholinergic signals (Fig S8). This enrichment of adrenergic and cholinergic-like neurons was also verified in the organ-specific heart and lung single-cell datasets (Fig S5).

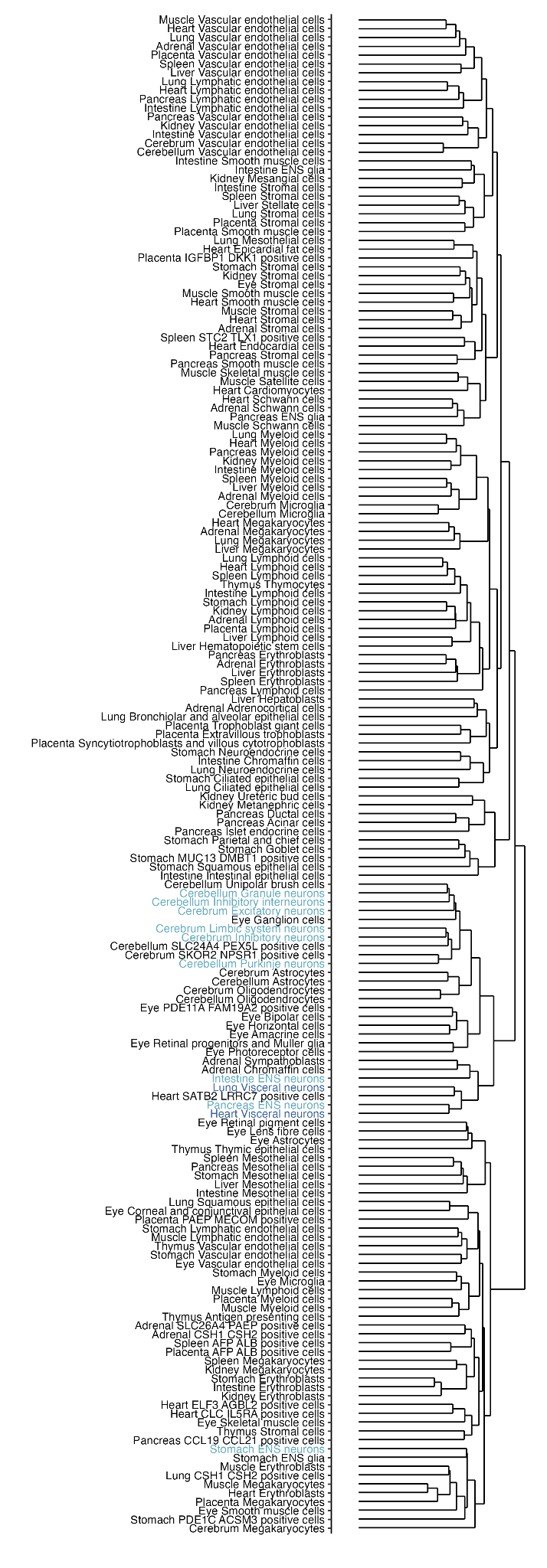

#### **Fig S7:** Dendrogram with hierarchical clustering (euclidean distance) of the cell types from the whole body, foetal developmental single-cell transcriptional dataset. Central and peripheral nervous system (CNS/PNS) neurons are highlighted in light blue and the visceral PNS neurons are in navy.

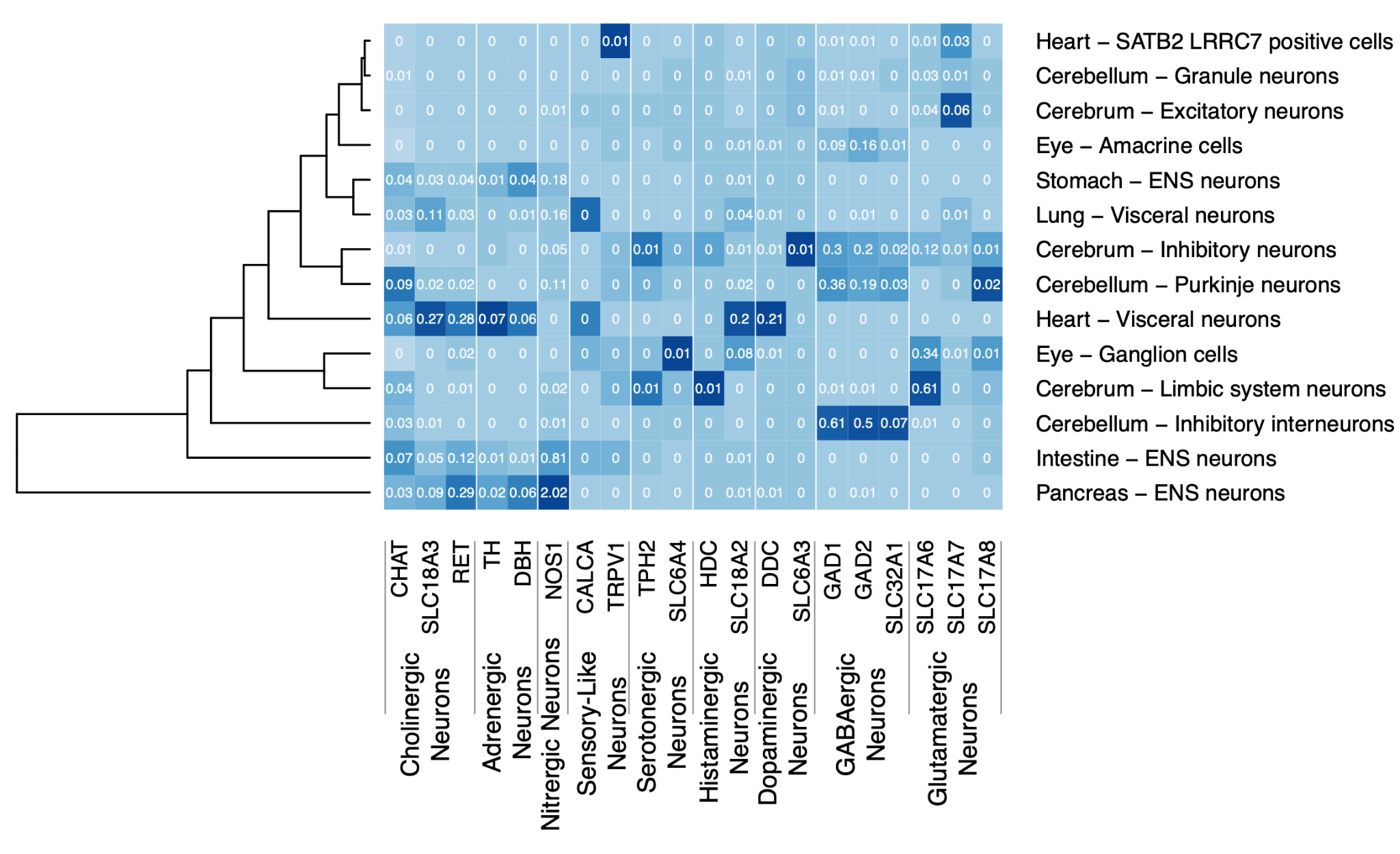

#### **Fig S8:** Heatmap of differing neuronal transcriptional signal from the foetal developmental whole body atlas (x-axis), across known neuronal marker genes (y-axis); cholinergic (*CHAT*, *CLC18A3* and *RET*)^30–32^, adrenergic (*TH, DBH*)^33^, nitrergic (*NOS1*)^34^, sensory-like (*TPH2*, *SLC6A4*)^35,36^), histaminergic (*HDC*, *SLC6A3*)^37,38^, dopaminergic (*DDC*, *SLC6A3*)^39^, GABAergic (*GAD1*, *GAD2*, *SLC32A1*)^40^ and glutamatergic (*SLC17A6*, *SLC17A7*, *SLC17A8*)^41^ neurons. Values represent the average expression and heatmap shade is scaled across all neurons (columns).

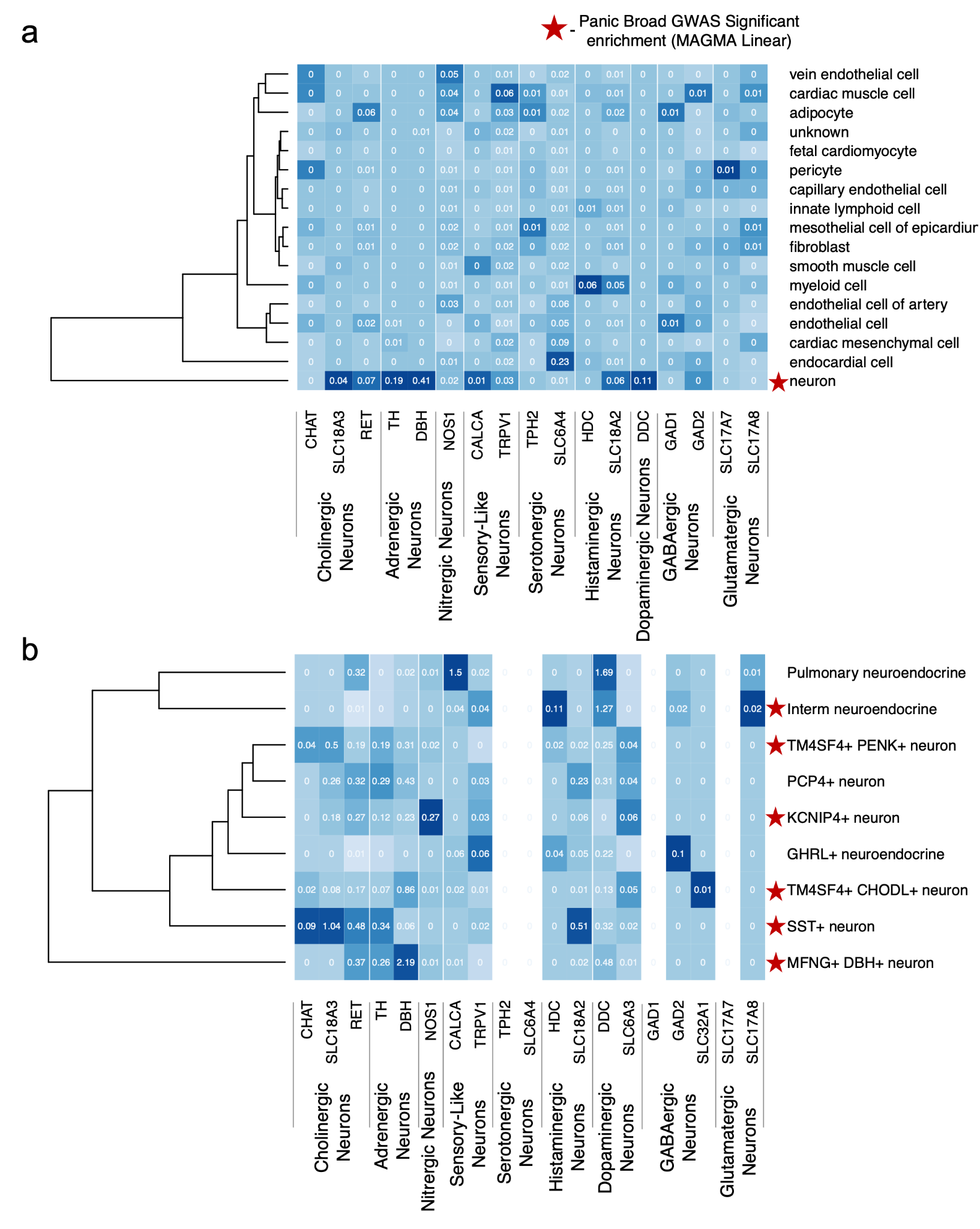

#### **Fig S9:** Heatmap of differing neuronal transcriptional signal from the (a) epicardium (heart) and (b) lung single-cell transcriptomic atlases (x-axis), across known neuronal marker genes (y-axis); cholinergic (*CHAT*, *CLC18A3* and *RET*)^30–32^, adrenergic (*TH, DBH*)^33^, nitrergic (*NOS1*)^34^, sensory-like (*TPH2*, *SLC6A4*)^35^, histaminergic (*HDC*, *SLC6A3*)^37,38^, dopaminergic (*DDC*, *SLC6A3*)^39^, GABAergic (*GAD1*, *GAD2*, *SLC32A1*)^40^ and glutamatergic (*SLC17A6*, *SLC17A7*, *SLC17A8*)^41^ neurons. Values represent the average expression and heatmap shade is scaled across all neurons (columns). The red star indicates significant enrichment for broad panic disorder.

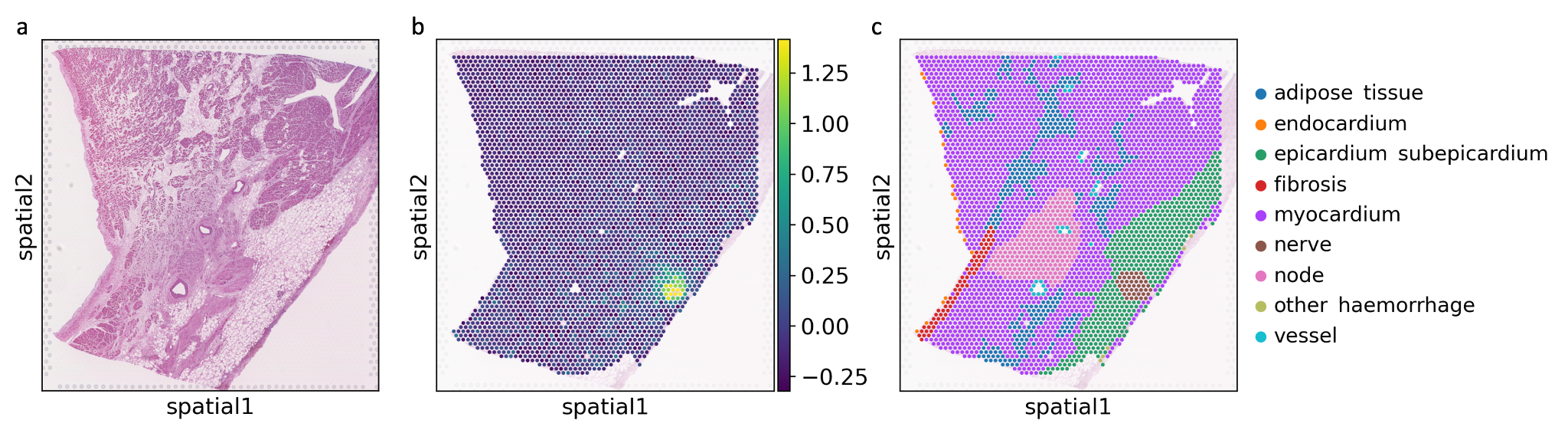

#### **Fig S10:** Spatial transcriptomic map of the sinoatrial node in the heart showing (a) a hematoxylin and eosin staining of the tissue, (b) a spatial projection of the visceral neuron marker gene score based on the heart visceral neurons from the foetal developmental transcriptomic dataset and (c) the structural tissue annotation. The signal resides in nerve ganglia within epicardial adipose tissue near the sinoatrial node.

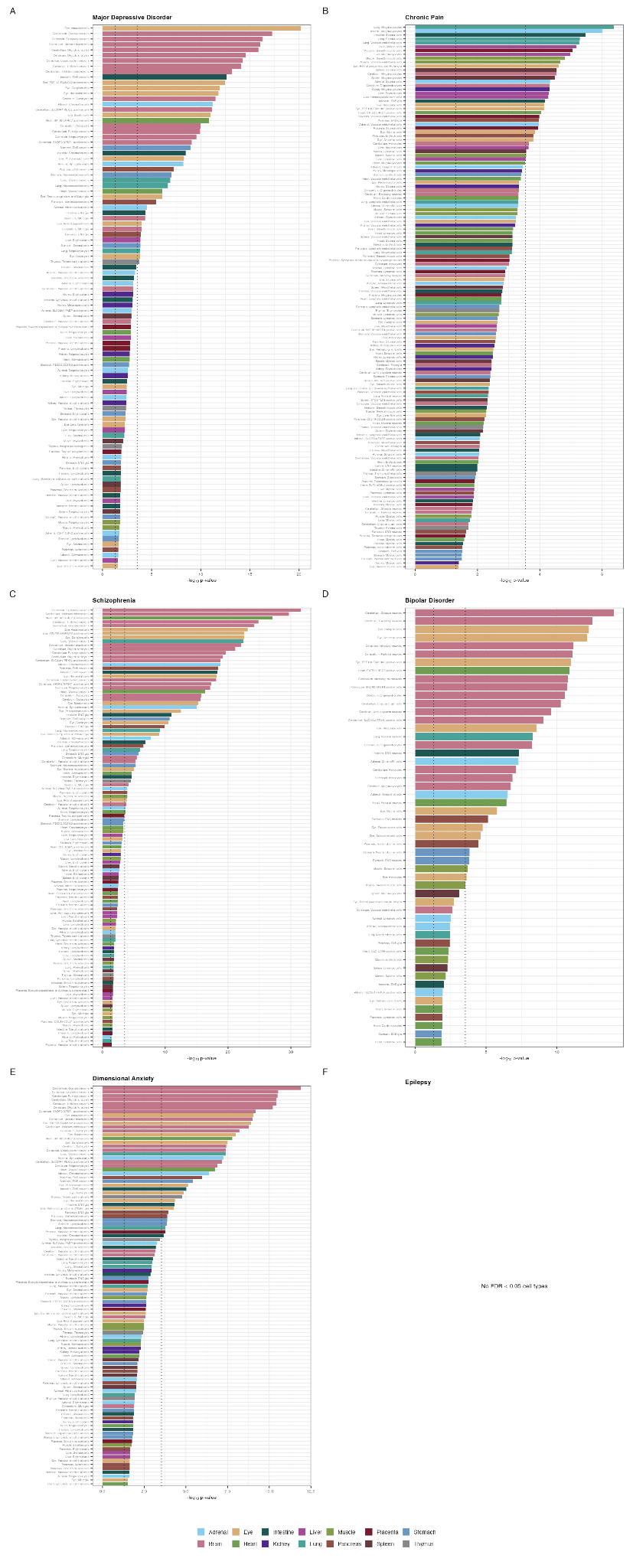

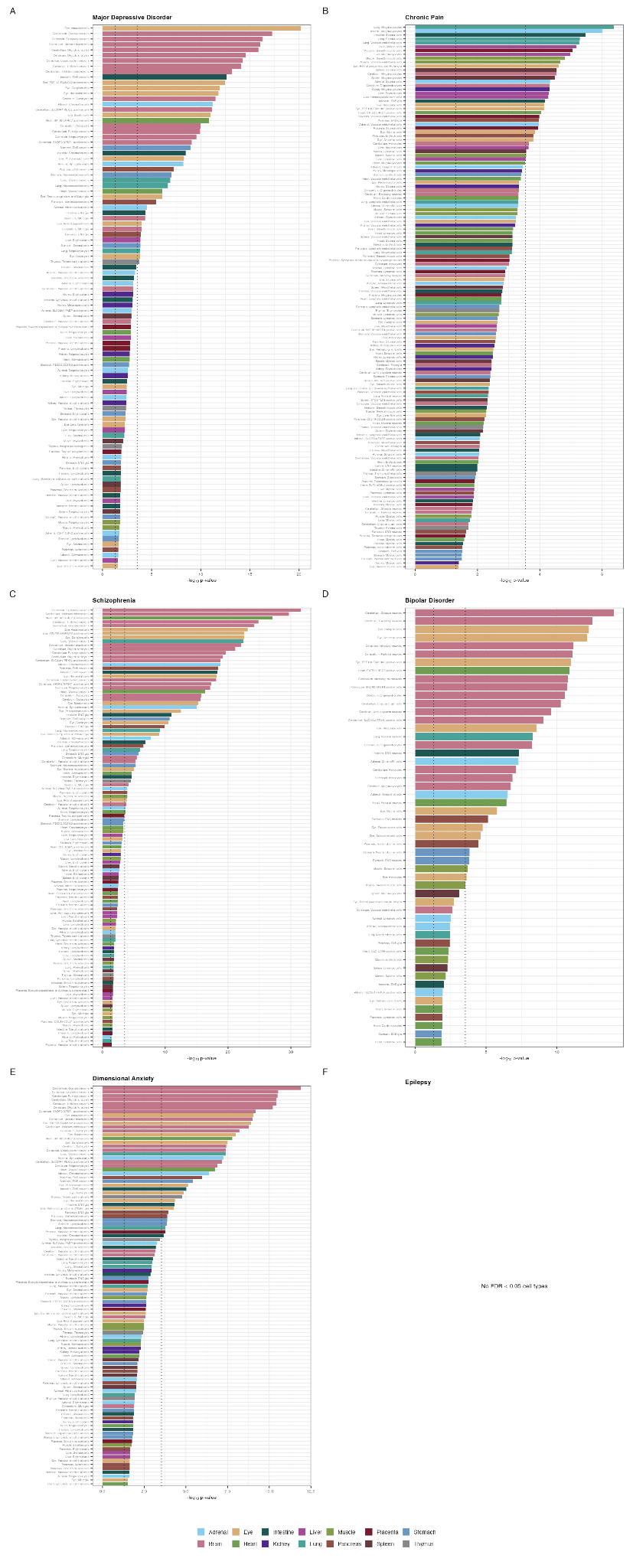

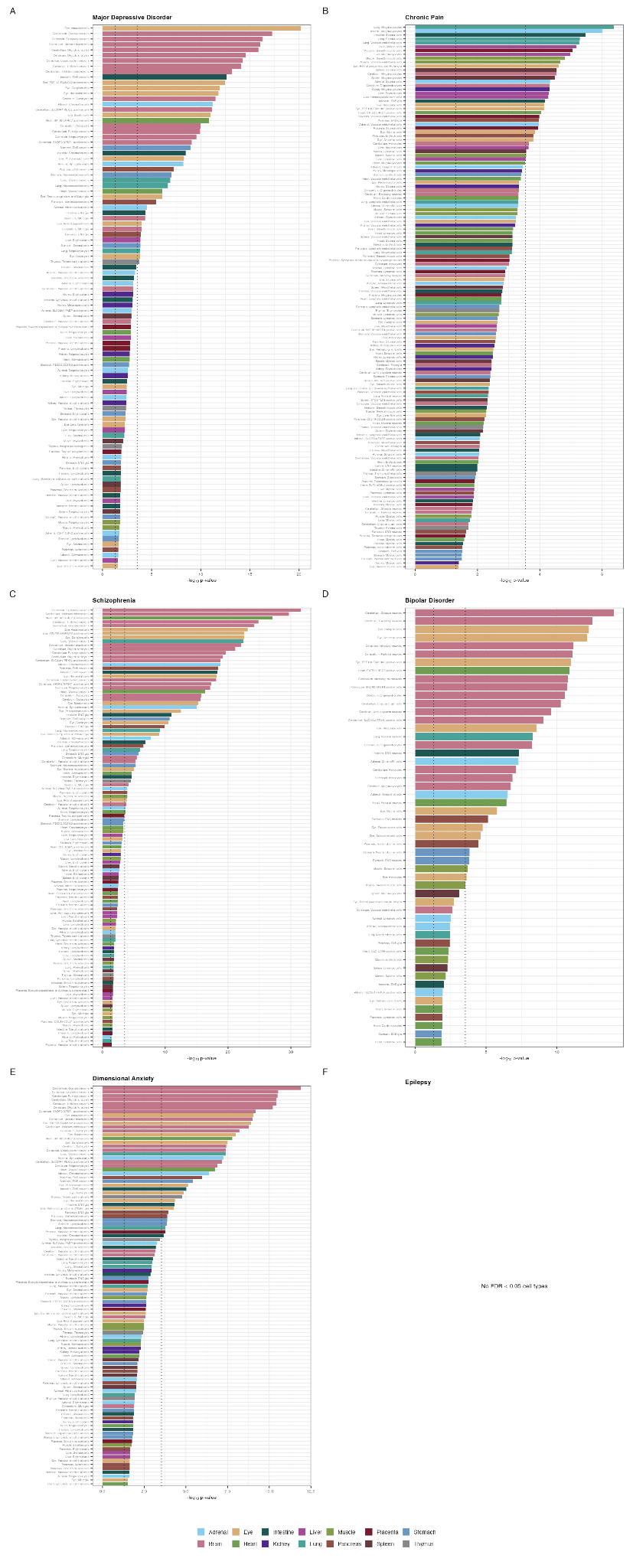

#### **Fig S11:** All cell type-specific, significant genetic enrichments (FDR < 0.05) based on the whole body, foetal developmental single-cell transcriptional dataset for GWAS of (a) major depression^42^ (b) chronic pain^43^ (c) schizophrenia^44^ (d) bipolar disorder^45^ (e) dimensional anxiety^46^ (f) epilepsy^47^. The MAGMA p-value enrichments are shown for linear regression. The black dashed vertical lines indicating p-value thresholds of 0.05 and Bonferroni adjusted p-value threshold of 0.0001. The y-axis shows the -log10 p-value where larger values indicate a greater level of significance.

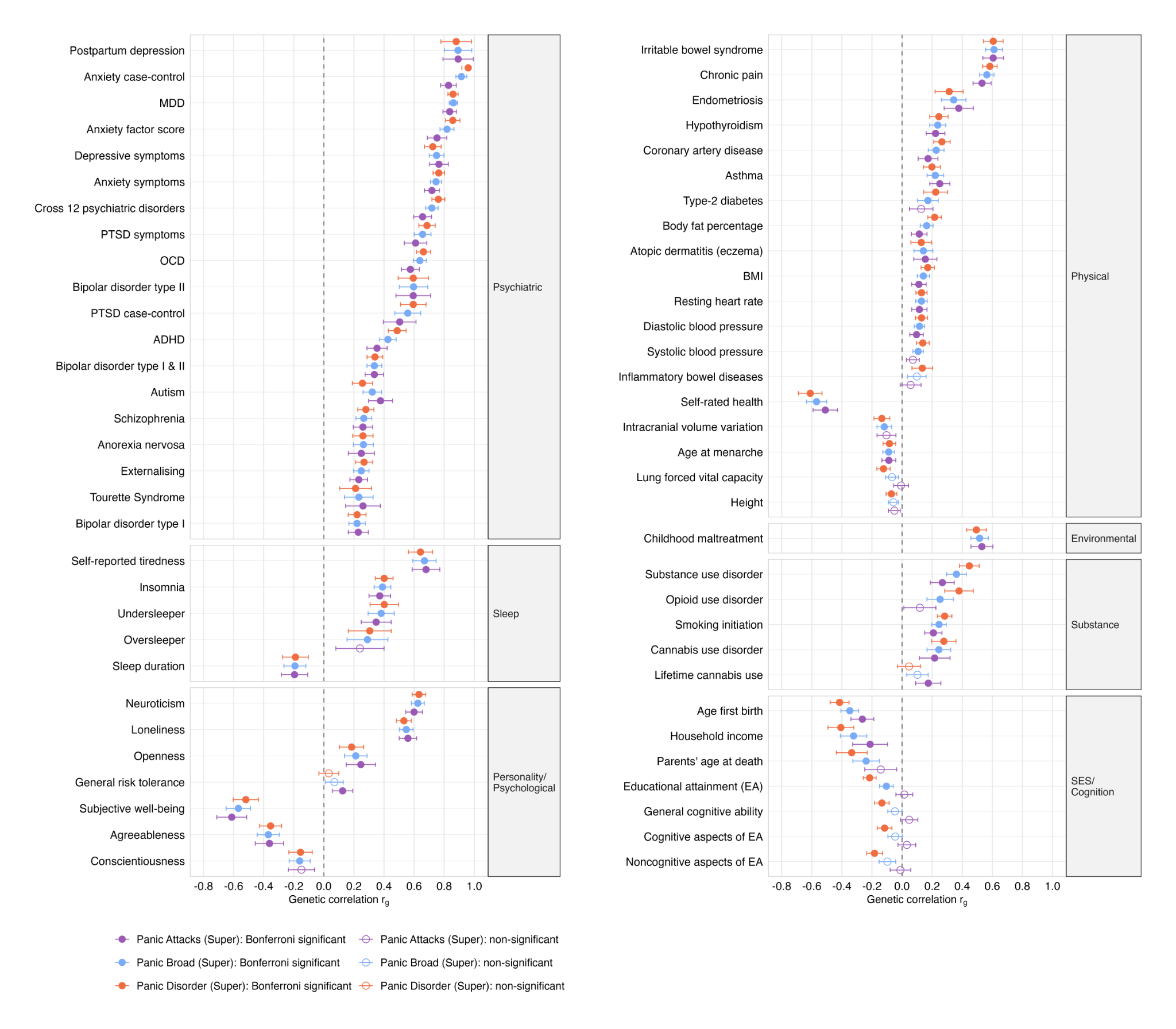

#### Figure S12: Genetic correlations with external traits using super screened controls
